# Origin and structural evolution of the complex genomic regions of human Y chromosome

**DOI:** 10.64898/2026.07.27.26359059

**Authors:** Jing Liu, Qingyang Ni, Yu Zhang, Quanyu Chen, Dan Yu, Chentao Yang, Dongya Wu, Guojie Zhang, Qi Zhou

**Author notes:** These authors contributed equally (Jing Liu, Qingyang Ni).

## Abstract

The human Y chromosome comprises mosaic classes of complex male-specific sequences that remain unclear for their evolution origins, structural variations and pathogenic roles. To address these, we present population-scale analyses of high-quality Y chromosomal sequences of 206 human individuals vs. six ape species, including 160 near complete sequences from the East Asian population, 85 of which are gapless. We uncovered an extensive cryptic diversity of the largest heterochromatin of the human genome on the Y chromosome Yq12, and traced its dual-origin from ancestral centromeric satellites respectively on the Y chromosome and acrocentric autosomes shared with other apes. We showed that such diversity is attributed to the variable and layered expansion of DYZ satellites in the internal array of Yq12, while the Yq12 boundary is marked by conserved inverted arrays. Similarly, the largest Y-linked gene family *TSPY* can be divided into a highly variable tandem array whose copy number was found to be associated with prostate cancer risks, and an invariable *TSPY2* under stronger functional constraints. Within the largest amplicon region AMPL7, we annotated the orientation and copy number per individual of the highly similar amplicons in palindromes, and characterized their recurrent inversion togglings and subsequent microdeletions impacting the male fertility. We further identified population-stratified structural variants in disease-associated genes, including some fixed mutations in *TSPY2* and the promoter of *DDX3Y* in the prevalent Asian haplogroups O1a that are probably associated with elevated prostate cancer risks. Together, our study revealed the mosaic internal structures and evolutionary history of respective complex regions of the human Y chromosome, and provided a foundation for understanding its functional role beyond male determination.

## INTRODUCTION

The human Y and X chromosomes originated from a pair of autosomes over 170 million years ago^1^, and homologous recombination was restricted within the two short pseudo-autosomal regions (PAR) at chromosome ends, probably after episodes of chromosomal inversions^2,3^. Except for some euchromatic regions that share the autosomal origin with, or were recently transposed from, the X chromosome (X-degenerate and X-transposed regions, XDR and XTR), most of the non-recombining male-specific region of the extant human Y chromosome (MSY) is composed of highly repetitive sequences that historically posed major challenges for sequencing and assembly^4,5^. This difficulty arises because majorities of MSY regions consist of long and composite repeated sequence units organised in extensive tandem or inverted arrays. The most prominent example is the largest heterochromatic region in the human genome that constitutes nearly half the MSY length in the q arm of Y chromosome (Yq12). Recent advances in long-read sequencing technologies have enabled the first complete telomere to telomere (T2T) assembly of a human Y chromosome (HG002-Y)^6^ and subsequent assembly of 43 globally diverse human individuals (‘43Ys’ hereafter)^7^, which achieved gapless resolution of the Yq12 heterochromatic region in seven individuals and revealed extensive structural and copy number variations (CNVs) in DYZ1/DYZ2 satellite array of Yq12. With the recently available gapless Y chromosomal sequences of six non-human ape species^8^, it is clear that such alternating DYZ structure^9,10^ is human-specific. However, the evolutionary trajectories of human-specific complex Y-linked structures remain unresolved, as incomplete assembly of ampliconic regions across diverse haplogroups are still lacking. This has left their copy numbers, orientations, differences between the units, therefore potential disease-associated functions unclear among the human populations.

By contrast, Y-linked euchromatic regions have been better assembled and annotated with most functional genes, including the male-determination (e.g. *SRY*, Sex-determining Region Y)^11^, dosage-sensitive housekeeping genes^12–14^ and multicopy gene families related to male-fertility, e.g., *TSPY* (Testis-specific Protein, Y-encoded)^15^ and *DAZ* (Deleted in Azoospermia)^16^. Many genes are embedded in ampliconic regions prone for pathogenic structural rearrangements including inversions, microdeletions and duplications. For example, there is a 7-fold higher rate of recurrent inversions (‘inversion toggling’) on the Y chromosome than autosomes^17,18^. This is because of the much more frequent non-allelic homologous recombination (NAHR) between the long inverted and nearly identical sequence segments, i.e., palindromes, and more distantly spaced inverted repeats (IRs) in ampliconic regions. And the *TSPY* of ampliconic region 2 (AMPL2), the largest Y-linked multicopy gene family, has undergone human-specific expansion and is implicated in both spermatogenesis regulation and as an oncogene for testicular and prostate cancer^19,20^. Similarly, microdeletions in the azoospermia factor regions (AZF), particularly in AMPL7 spanning different amplicon classes are associated with increased risks of oligozoospermia or azoospermia (low or no sperm production)^21–24^. However, extensive assembly gaps still exist in most published Y chromosomes: 33 of the 43 Ys remain incomplete in at least one ampliconic region^7^, leaving the population-level diversity and functional impacts of these disease-associated structural variations (SVs) insufficiently explored across human populations.

Finally, an underrepresentation of East Asian (EAS) samples in the recently published Y chromosomal sequences (14% of 43Ys)^7^ motivated us to produce here a high-quality dataset of 160 EAS Y chromosomes, half of which have reached a gapless state, representing the largest collection of gapless Y chromosomes from a single population to date. Combined with the published near-complete Y chromosomal sequences of human^6,7,25^ and apes^8^, our analyses covered all the major Y haplogroups across population, enabling unprecedented characterization for the diversity of the complex regions of MSY, i.e., the Yq12 heterochromatin and the AMPL regions of euchromatin. These results revealed recurrent structural variants (SVs), including copy number expansions, inversion togglings and microdeletions, that are associated with male infertility and male-specific cancer risk, and provided insights into the mosaic evolutionary history and functional consequences of Y chromosome structural diversity.

## RESULTS

### Genome sequences and size variation of the 160 East Asian Y chromosomes

We generated high-quality Y chromosome assemblies from 160 EAS males (160Ys) representing 13 major (sub)haplogroups^26–28^ (**Figure 1A**; **Table S1**), and achieved 85 gapless assemblies representing the largest collection of complete Y chromosomes from a single population to date (**Figure 1A**). This dataset substantially expands representation of haplogroups previously missed (e.g., the ancient D1 haplogroup dominant among Tibetans^29,30^) or underrepresented (e.g., the most prevalent O1/2 haplogroups among the East and Southeast Asia^30,31^) in the 43Ys^7^ (**Figures S1A-B**). For the remaining 75 samples, we generated high-quality pseudo-chromosome sequences by scaffolding contigs (median: 4 per sample) using the HG002-Y reference, (**Table S2**), followed by comprehensive annotation of 24 subregions, 36 single-copy protein genes and 9 multicopy gene families across all 160 Y chromosomes (**Figures 1B** and **S2**).

**Figure 1.**
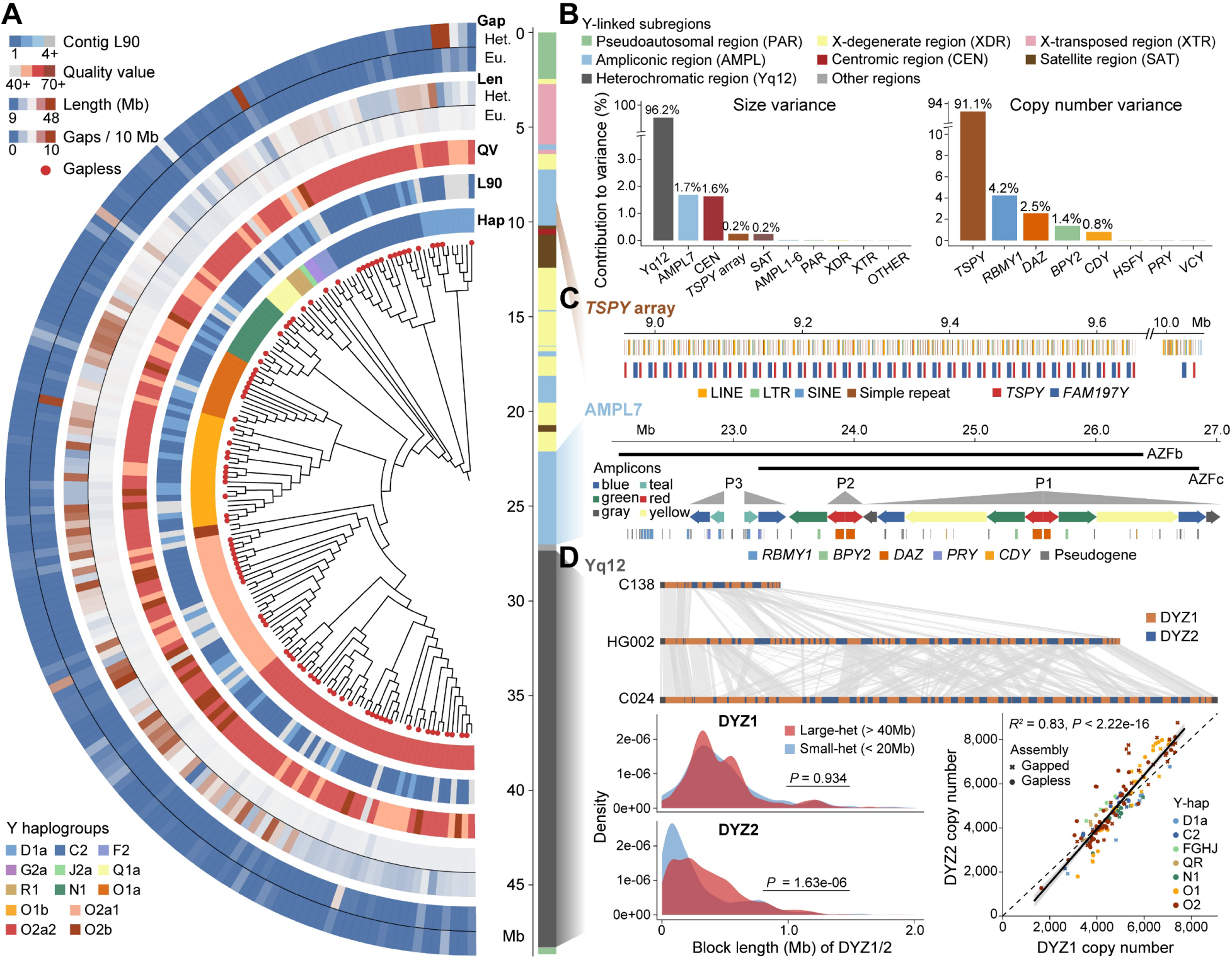
Y chromosomal sequences of 160 East Asian individuals and their length variation. **A)** From the inner to outer circle, we show the haplogroup phylogeny, the 85 gapless Y chromosomal sequences marked by red dots, the haplogroup information, the contig L90 values (a smaller value indicates greater assembly continuity), the assembly quality value (QV), the assembly length (Mb) and gap number (per 10Mb) separately for the euchromatin (Eu.) and heterochromatin (Het.) regions. **B)** 24 subregions of a gapless Y chromosome (sample C002-CHA-E02) followed the previous definition^4^. The left bar plot shows the relative contribution of these subregions to the overall Y chromosomal length variation among individuals. The right bar plot shows the relative contribution of different multigene families to the overall copy number variations of Y-linked genes. Both are calculated using gapless Y chromosomal sequences. *TSPY*: Testis-specific protein, Y-encoded. *RBMY1*: RNA-binding motif gene 1 on Y chromosome. *CDY*: Chromodomain Y-Linked 1. *DAZ*: Deleted in Azoospermia. *BPY2*: Basic Charge Y-Linked 2. *HSFY*: Heat Shock Factor Y-Linked. *PRY*: PTPN13 Like Y-Linked. *VCY*: Variable Charge Y-Linked. **C)** The genomic structure of the *TSPY* array and the ampliconic region AMPL7. The *TSPY* array is located at AMPL2 and consists of between 19-45 repeated units each containing the *TSPY* gene, the non-coding gene *FAM197Y* and composite repeats among the studied individuals in this work. AMPL7 is the largest Y-linked ampliconic region that overlaps with the azoospermia factor (AZFb/c) regions, several palindromic regions (P1-P3) and multiple amplicons shown by colored arrows in inverted pairs. **D)** The Yq12 is the largest heterochromatin region in the human genome, and primarily consists of alternating arrays of DYZ1 and DYZ2 satellite repeats. This region is the predominant source of Y chromosome length variation, exemplified by the syntenic plot between three individuals with gapless Y chromosomal sequences (C138-CMH05, HG002 and C024-CHA-S04, bar length scaled to the Y chromosome length). The DYZ1/2 alternating repeats form continuous blocks, whose sizes have significant differences between individuals, and the copy number is almost the same between DYZ1 and DYZ2 repeats of the same individual.

Our assemblies demonstrate superior quality metrics compared to previous datasets, with average contig N50 of 46.2 vs. 19.1Mb in the 43Ys^7^, and reduced contig L50 numbers (1.2 vs. 2.5) (**Figure S1C**), reflecting the advance of higher sequencing coverages and longer read lengths^32^. Besides the 85 gapless Y sequences, nearly all samples achieved complete assembly across most subregions (99.7% gapless on average), with remaining gaps concentrated in the PAR1, centromeric, AMPL6 and 7, and Yq12 subregions (**Figure S1D**; **Table S4**). These regions also showed substantially improved contiguity over published assemblies (mean percentage of gapless sequence: 86.5% of the 160Ys vs. 37.7% of the 43Ys), though they are prone for different types of assembly challenges^6,7^. PAR1 exhibits frequent phase-switching errors (64.1%) (**Figures S3-4**) due to its exceptionally high recombination rate^33,34^, while Yq12, centromeric and ampliconic regions tend to have collapsed tandem repeats detectable through coverage analysis (**Figure S3**). After systematic error detection and manual curation (**Table S3**; **METHODS**), flagged errors comprised only 0.0052% to 1.32% (median proportions) of total length among the 160Ys (**Table S5**). The resulting assemblies achieved high accuracy at both nucleotide levels (median quality value (QV): 63.4) (**Figures 1A** and **S1E**) and structural level (median Genome Continuity Inspector (GCI score^35^: 55.4), with five samples showing an equal or higher QV value^36^ compared with the published HG002-Y (QV = 73)^6^, and 66 having a GCI value of 100, i.e., without any detected structural errors (**Table S2**).

These high-quality Y chromosomal assemblies revealed extensive structural diversity across the East Asian population, with the total length ranging from 37.3Mb to 76.0Mb (median: 56.4Mb). This is predominantly attributed to the CNVs of the DYZ1 and DYZ2 repeat units in the Yq12 heterochromatin, which spans 9.0 to 45.8Mb (median: 28.3Mb) and constitutes 26.2 to 65.2% of the 85 gapless Y chromosomes, comprising the largest and most variable MSY subregion. Centromeric (CEN) and ampliconic regions AMPL2 and 7 also exhibit great length variations to a lower extent, while euchromatic regions (PARs, XDR and XTR) remain nearly invariant in length (**Figure 1B**). We resolved the precise structure, orientation and copy numbers of the genes and repeat units within these complex regions across most individuals. In particular, AMPL2 contains the *TSPY* gene family, which varies from 19 to 45 copies (median: 33) across the studied individuals; while AMPL7 contains multigene families including *RBMY1* (RNA-binding motif gene 1 on Y chromosome, median copy: 8) and *DAZ* within large palindromes (P1-P3) that overlap AZFb/c (**Figures 1C** and **S5**; **Table S6**). These three gene families account for major sources of euchromatin length variation and are more prone for CNVs than those elsewhere on the Y chromosome. Interestingly, the copy numbers of *TSPY* and *RBMY1* exhibit a significant negative correlation (Spearman’s correlation, *R* = −0.24, *P* = 0.000645; **Figure S5**) among the examined 160 individuals, implicating their possible complementary functions in male fertility. And both families display haplogroup-specific expansion (e.g., *RBMY1* in the C2 haplogroup) or contraction (e.g., *TSPY* in the N1 haplogroup) that warrant functional investigation (**Figure S6**). Within the Yq12 heterochromatin, CNVs of the alternating DYZ repeat block (continuous DYZ1 or DYZ2 units) produced up to five-fold length difference between individuals (e.g., individual C138-CMH05 vs. C024-CHA-S04; **Figures 1D** and **S7**). Although the copy numbers of DYZ1 and DYZ2 units are highly correlated within individuals (Pearson’s correlation, *R* = 0.91, *P* < 2.22e-16), indicating coordinated expansion or contraction (**Figure 1D**), DYZ2 contributes disproportionately (64.0% vs. 36.0% of DYZ1) higher to the overall length variation of Yq12, due to its significantly (W = 100119, *P* = 1.63e-06, Wilcoxon rank sum test) larger difference of block sizes between individuals with long Yq12 (> 40Mb, *n* = 10) vs. those with short Yq12 (< 20Mb, *n* = 7) (**Figure 1D**).

We further refined the pseudoautosomal boundary, a long-debated feature of Y chromosome architecture^7,37,38^. PAR1 expectedly shows a sharp reduction of genetic diversity (measured by single nucleotide polymorphism (SNPs) and SVs per 10Kb, **Figure S8**) at its boundary with the non-recombining MSY. Through expanded variant calling across our dataset (**METHODS**), we narrowed the PAR1 boundary to a 116 bp interval toward the distal end of previously defined coordinates^38^, providing the most precise localization to date. Given the exceptional structural diversity and functional importance of AMPL2/7 and Yq12, which encompass fertility-associated gene families and disease-linked regions, we focused subsequent evolutionary analyses on the most variable euchromatic and heterochromatic MSY regions to reconstruct their origin and diversification across human populations.

### Dual-origin, evolution and transcription of the human Yq12 heterochromatin

Complete Yq12 sequences from 116 humans (107 from this work (**Table S4**), 7 from the 43Ys^7^, HG002-Y^6^ and CN1-Y^25^) and six ape species^8^ enable us to reconstruct the evolutionary origin of this massive human-specific heterochromatin and trace its subsequent expansion or contraction across populations. The DYZ1 satellites are human Y-specific members of Human Satellite 3 (HSat3) A6 subfamily^39^, and DYZ2 satellites belong to the HSat1B clade comprising an AT-rich simple repeat, a 5′ truncated *Alu*Y element and a GC-rich HSATI region^39–41^. The alternating structure of DYZ1/DYZ2 blocks constituting the Yq12 is absent on all other human chromosomes^42^ and all non-human ape chromosomes ^8,43^, yet related satellite families provide clues to its origin. Tandem arrays of other HSat3 subfamilies (A3 and A4), but almost no DYZ2 satellites can be found on the human Y chromosome outside the Yq12 region, nor on the Y chromosomes of other apes (**Figures 2A** and **S9A**). This suggests the presence of Y-linked DYZ1-like HSat3 satellites in the ancestor of apes followed by human-specific insertions of autosomal DYZ2 onto the Y chromosome.

**Figure 2.**
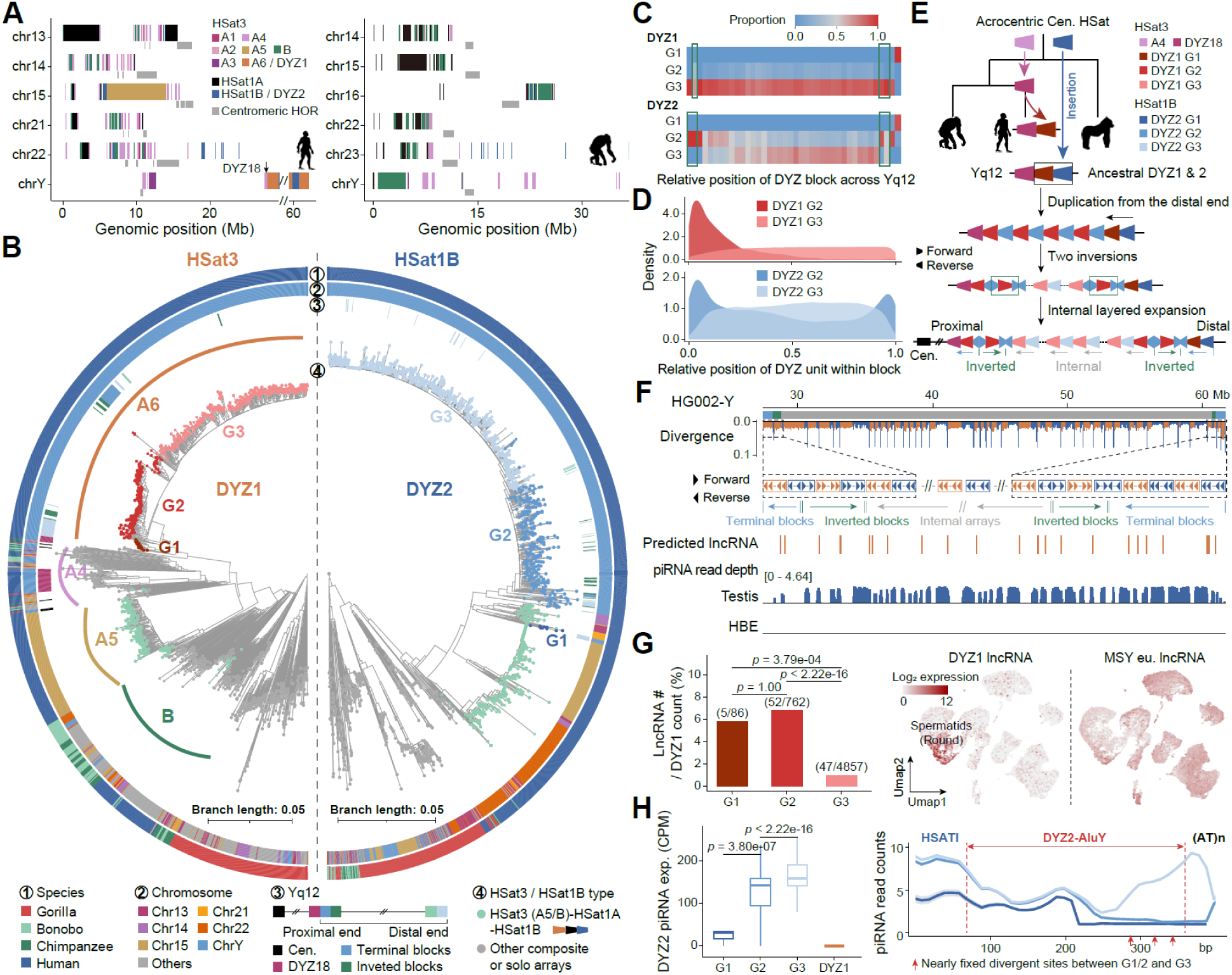
Dual-origin, evolution and transcription of the Yq12 heterochromatin. **A)** The DYZ1 and DYZ2 are respectively a subfamily of human satellite 3 (HSat3) and HSat1B satellites. We show here for human (CHM13v2.0 for autosomes, HG002-Y for chrY) and chimpanzee (mPanTro3) their Y chromosomes and other five autosomes with the highest abundance of HSat3, HSat1A, and HSat1B. The horizontal gray bars of each chromosome indicate the active HORs (higher-order repeats) of centromeric regions. DYZ18 (HSat3-A4) is pointed out with arrows in the panel. **B)** Phylogenetic trees of HSat3 and HSat1B repeats from genomes of great apes. The HSat3 tree was inferred from a k-mer based Mash-distance matrix of randomly downsampled repeat units using the neighbor-joining method (**METHODS**). For HSat1B, alignments of concatenated *AluY* and HSATI satellite sequences were used to reconstruct a maximum-likelihood (ML) tree. The circos plot from the outer to inner rings show (1) species, (2) chromosomal locations, and (3) different types of Yq12 blocks defined in Figure 2f and (4) different combinations of satellites on autosomes. The dots with light green represent the HSat3 or HSat1B repeats from the composite array, HSat3-HSat1A-HSat1B, in human autosomal (chr13/14/15/21/22) (peri-)centromeric regions, whereas the gray ones represent the repeats with other composited structures (e.g., HSat3-HSat1A) or solo HSat array. DYZ1 repeats belong to the HSat3-A6 subfamily, whose closest lineage is HSat3-A4. Both DYZ1 and DYZ2 repeats on Y chromosomes can be divided into three subgroups (G1-G3). **C-D)** The positional distribution of three subgroups of DYZ1 or 2 across the Yq12 region. **E)** The dual-origin model for the DYZ repeats in Yq12. DYZ1-G1 likely originated from the ancestral Y-linked HSat3-A4 (in human, it is termed ‘DYZ18’ adjacent to Yq12) sequences and later formed an alternating DYZ1-DYZ2 unit with an autosomal HSat1B copy (DYZ2-G1) that inserted into the most distal end of human Y chromosome. Subsequent amplifications, likely seeded from this composite unit, gave rise to the primary alternating DYZ1/DYZ2 blocks. Two inversions toward the end of the Yq12 then set the boundary, after which units in internal regions underwent further amplification, mainly involving the younger DYZ1-G2/3 and DYZ2-G2/G3 units. **F)** The two peripheral inversions partitioned the human Yq12 region into terminal, inverted, and internal arrays. The DYZ1 repeats encode non-coding RNAs (lncRNAs), whereas the DYZ2 repeats encode piRNAs specifically in testis. HBE: human bronchial epithelial. **G)** Predicted lncRNA gene models among DYZ1 subgroups, normalized by DYZ1 subgroup copy number (left panel). These transcripts are predominantly detected in round spermatids, in contrast to lncRNAs derived from euchromatic regions of the MSY (right panel). **H)** Testis piRNA expression across the DYZ2 subgroups. The DYZ2 G3 subgroup has the highest piRNA levels. The right panel illustrates differences in piRNA depth among subgroups. Red arrows indicate nearly fixed sequence differences between G2 and G3 units. CPM: Counts Per Million.

Our phylogenetic analyses of all annotated DYZ1 and DYZ2 copies of the HG002-Y, and randomly selected copies of all subfamilies of HSat3 and HSat1B throughout the human (T2T-CHM13v2.0)^44^ and other great ape genomes, uncovered that all DYZ1 copies are clustered with the HSat3 A4 subfamily mainly located in centromeres of the human and ape Y chromosomes (**Figure 2B**). Specifically, some HSat3-A4 copies (termed ‘DYZ18’, **Figure 2A**) are juxtaposed with the Yq12, which possibly represents the ancestral state of Yq12 before the DYZ2 insertion, although we cannot completely exclude that autosomal A4 copies also contributed to the origin of DYZ1 satellites. While all Y-linked DYZ2 copies cluster with the HSat1B copies mainly located in (peri-)centromeric regions of human chr13/14/21 (**Figure 2B**). We have found almost no solo HSat1B copies in human and other ape genomes. Most HSat1B copies formed a composite structure with HSat1A and HSat3 subfamilies (mostly A5) on acrocentric chromosomes chr13/14/15/21/22 in humans, or another structure only with HSat3 in non-human apes (**Figures 2A-B**). These HSat1B-containing composite repeats do not contain the HSat3 copies that are most closely related to, or have the same orientation with, the extant DYZ1 copies of Yq12. They likely share the ancestral HSat1B copies that give rise to the Y-linked DYZ2 through independent formation of composite repeats (**Figure 2E**). In summary, these results suggest that the Yq12 DYZ1/DYZ2 units have a dual-origin through combining an ancestral Y-linked mutated variant of HSat3-A4 satellite (namely, A6) with an autosomal HSat1B satellite that was specifically recruited to the human Y chromosome (**Figures S9B** and **2E**). This was followed by human-specific coordinated expansion that generated the observed copy number correlation between DYZ1 and DYZ2 within individuals (**Figure 1D**).

To understand how this alternating DYZ unit propagated to massive copy numbers specifically on the human Y chromosome, we utilized phylogenetic clustering (**Figure 2B**) by incorporating ape homologous repeats to classify DYZ1 and DYZ2 copies of all individuals into three sub-groups (G1-G3; **Table S7**). These sub-groups were further supported with principal component analysis (PCA) based on k-mer frequencies (**Figures S10**) and show distinct divergence from their consensus sequences (**Figures S11**). The spatial distribution of these hierarchically defined sequence clusters reveals a non-uniform chromosomal organization that is informative for understanding DYZ array expansion dynamics. The oldest copies of both DYZ1 and DYZ2 (G1) are exclusively found at the most distal block of the Yq12 (**Figures 2C** and **S12A**) and have the lowest CNV across individuals (**Figure S12C**), suggesting this region likely represents an ancestral insertion site of DYZ2. The middle-aged copies (G2) are enriched within a pair of inverted blocks near both Yq12 boundaries^7^, with the proximal inverted block containing a higher proportion of G2 copies than the distal one, implicating sequential inversion events (**Figure S12B**). The youngest copies (G3) which show the lowest divergence from the subgroup-specific consensus sequences, are enriched within the ‘internal arrays’ between the inverted boundary blocks that comprise the largest and most variable copy numbers of DYZ1/2 (**Figures 2C** and **S12**). This pattern reflects local homogenization states within arrays, likely driven by ongoing gene conversion coupled with more recent expansion events.

Within each array containing multiple copies of DYZ1 or DYZ2, the older G2 copies are consistently localized to one or both ends of each block (**Figure 2D**), while younger copies occupy central positions. This layered architecture parallels the expansion mechanism of human centromeric repeats^42^, wherein new satellite copies arose both within and between blocks (**Figures 2C-D**) while the older flanking copies diverged in sequences and were eventually replaced or deleted (**Figure 2E**). The expansion of DYZ1/2 satellites was likely initiated from the most distal Yq12 end following the emergence of composite repeats containing the ancestral HSat3-A4 (DYZ18) and acrocentric HSat1B (**Figure 2E**). However, Yq12 differs fundamentally from centromeres in lacking diverse satellite families or large segmental duplications at its border (the non-satellite ‘centric transition’ region^42^). Instead, each Yq12 boundary features a ‘terminal block’ with the same orientation with the internal array, and an adjacent inverted block (**Figure 2F**). This boundary organization is associated with a marked reduction in copy number variability in terminal and inverted blocks compared with internal arrays (1.65-3.20-fold lower; **Figure S12C**). One possible explanation is that inverted and terminal blocks may participate in a higher-order chromatin configuration that constrains local recombination or replication-based expansion.

What is the potential function of such a large Y-linked heterochromatin? We found that the DYZ1 and DYZ2 satellite units respectively encode long non-coding RNA (lncRNA) and PIWI-interacting RNA (piRNA) (**Figure 2F**) with testis-specific expression patterns that correlate with the satellite age. Using the published testis Iso-Seq data, we observed that the older DYZ1 satellites (G1 & G2) harbor a higher number of annotated lncRNAs (**Table S8**) per copy than the younger satellites (**Figures 2G** and **S13**), and the expression of these DYZ1-encoded lncRNAs are restricted to round spermatids (**Figures 2G** and **S14**). This is in contrast to the euchromatic MSY lncRNAs, which are transcribed across all testicular cell types. Conversely, DYZ2 satellites encode adult testis-specific piRNAs (**Figure 2F**), with younger DYZ2 copies producing significantly a higher transcription level of piRNAs than older copies (*W* = 1905816, *P* < 2.22e-16, Wilcoxon rank sum test; **Figures 2H** and **S15**), which can be discriminated by their nearly fixed sequence differences within the *AluY* regions (**Figures 2H** and **S16**). Together, these results uncovered that Yq12 heterochromatin likely plays a regulatory role in spermatogenesis through copy number-dependent expression of non-coding RNAs, with substantial variation in transcriptional outcome across individuals reflecting the extensive structural diversity of this region.

### Functional stratification of human *TSPY*/*FAM197Y* array since the ancestor of apes

The *TSPY* gene originated after an inversion of a proto-Y chromosomal region dated to the ancestor of eutherians^45^, and underwent subsequent independent expansions of copy numbers across ruminant^46^ and primate lineages^8^. Although the copy numbers of *TSPY* tandem array and its reversed paralogous copy *TSPY2* within the inverted repeat 3 (IR3) have been well annotated in previous ggapless Y chromosomal assemblies^6,7^, the composite transposable elements flanking each *TSPY* gene (**Figure 1C**) obscured the resolution of the precise repeated unit structure of tandem arrays, thereby hindering reconstruction of their origin and expansion processes in the human lineage. We found that a non-coding RNA gene with unknown function, *FAM197Y* (Family With Sequence Similarity 197 Y-Linked Member), is situated downstream of *TSPY2* and also adjacent to nearly every copy of the human *TSPY* tandem array (**Figure 1C**), forming a composite unit. Homologous *TSPY* units consisting of *TSPY*, *FAM197Y* and their encompassed intergenic region can be also found as a single or dispersed unit in the chimpanzee or the bonobo (**Figure 3A**), as a six-copy tandem array in addition to one distal unit with the same orientation (in contrast to human) in the gorilla, but are either not forming a tandem array in the orangutan or organised as two dispersed units with a different orientation in gibbon (**Figure 3A**). This phylogenetic pattern suggests that the *TSPY/FAM197Y* unit structure likely originated in the Homininae ancestor. Epigenetic data further supported this organization, where adjacent tandem array units are demarcated by enriched DNA methylation, promoter-associated histone modification H3K4me3, and reverse-oriented binding sites of the CTCF (CCCTC-binding factor) protein (**Figure S17**). This suggests that each repeat unit represents an evolutionarily conserved structural unit with distinct epigenomic boundaries. Nevertheless, we detected no obvious transcriptive or regulatory evidence of *FAM197Y* genes, and their function warrants future investigation.

**Figure 3.**
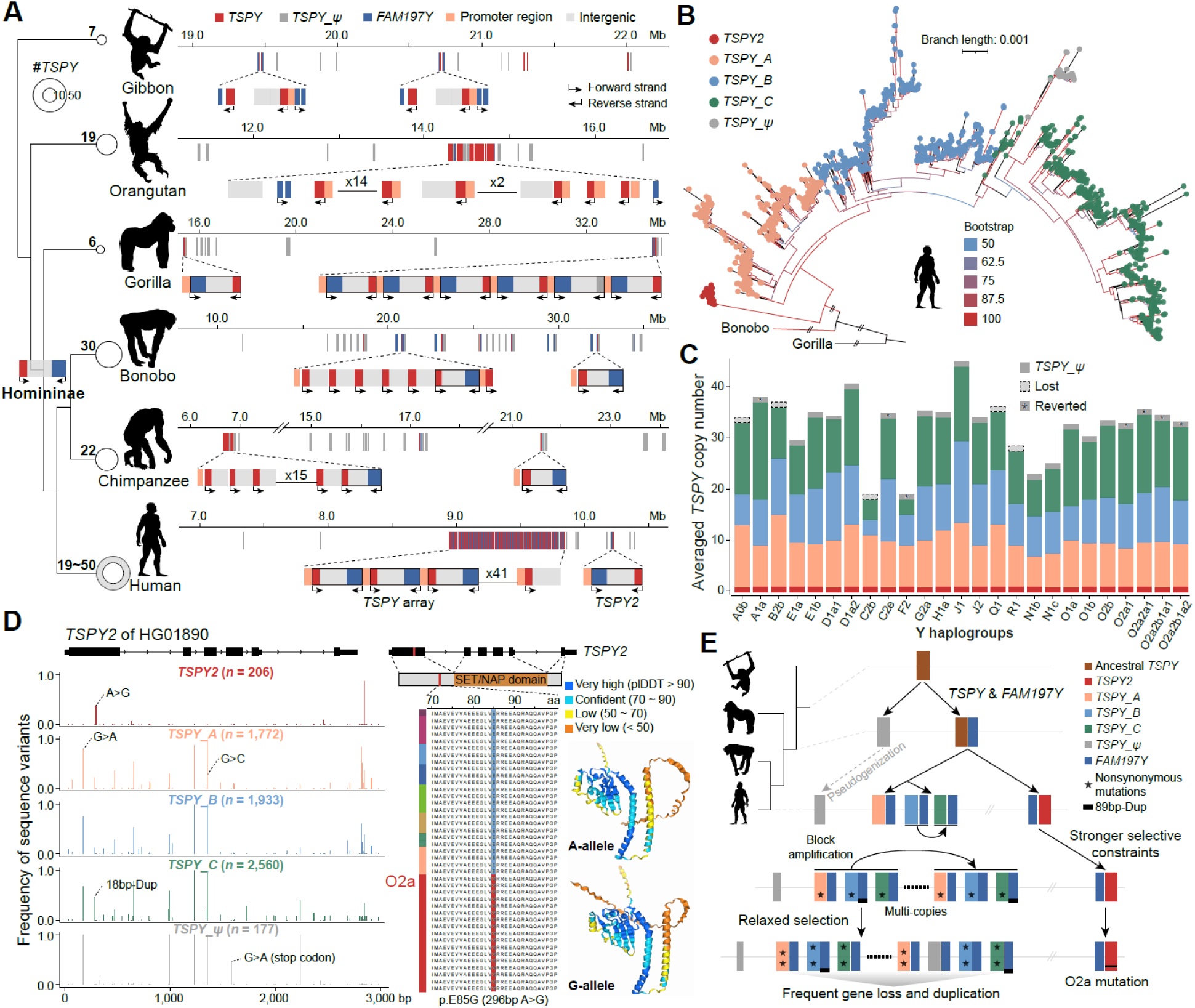
Origin and evolution of *TSPY* genes among great apes. **A)** Evolutionary origin of the *TSPY/FAM197Y* gene unit among great apes. The size of the circle is scaled to the number of *TSPY* gene copy numbers for each species on the phylogenetic tree, with two circles showing the range of *TSPY* CNVs within the studied human individuals in this work. We inferred that the *TSPY*/*FAM197Y* composite unit originated in the common ancestor of Hominidae, then underwent extensive expansion of copy numbers specifically in humans and formed two clusters, the *TSPY* tandem array and *TSPY2*. **B)** Maximum likelihood (ML) tree constructed with the *TSPY/FAM197Y* unit sequences including their intergenic region across human Y haplogroups. Four major clades can be identified: *TSPY2* (red), *TSPY_A* (orange), *TSPY_B* (blue), and *TSPY_C* (green). Pseudogenes (*TSPY_ψ*, gray) are embedded within the *TSPY_C* cluster. Branch colors indicate bootstrap supports. Y-linked homologous sequences of *TSPY*/*FAM197Y* of gorilla were used to root the tree. **C)** Average copy number of *TSPY* subgroups across Y haplogroups. In some haplogroups, the annotated *TSPY* pseudogene has lost its premature stop-codon mutation (G>A, ‘Reverted’; Figure 3D), while some other pseudogenes have completely lost (‘Lost’). **D)** Mutation profiles of *TSPY* genes using the *TSPY2* of an individual (HG01890) of A0b haplogroup as reference, which represents the basal lineage of modern human Y chromosomes. Only the alleles with MAF (Minor Allele Frequency) >= 0.05 in at least one *TSPY* sub-group are shown. Among them, the missense mutation specific to the O2a haplogroup (p.E85G, outside the SET/NAP domain) was predicted to affect the 3D protein structure of the TSPY. **E)** Evolution of the *TSPY* gene family. After forming the *TSPY*/*FAM197Y* unit in the Homonidae ancestor, the *TSPY* array underwent frequent gain and loss of copy numbers, whereas *TSPY2* remained conserved as a single copy, implicating stronger selective constraints on the latter. Frequent gene conversions probably have contributed to the homogenization and functional maintenance of the *TSPY* array. Notably, a O2a haplogroup specific missense mutation occurred in the *TSPY2*.

Previous study distinguished the human protein-coding from the pseudogene *TSPY* copies and resolved species-level differences through phylogenetic clustering of exonic^7^ or genic sequences^6^ (**Figure S18**), but did not determine whether sequence and functional distinctions exist between the highly similar copies of *TSPY* protein-coding genes. By expanding the phylogenetic analyses to the entire *TSPY/FAM197Y* sequences across all 206 individuals with available Y chromosomal sequences (**Table S9**), we divided their *TSPY* genes into four subgroups, including the *TSPY2* genes as a subgroup positioned at the basal clade, and three tandem array subgroups (*TSPY_A, B* and *C*) (**Figure 3B**). This finer classification proved to be robust when only using the *FAM197Y* or the intergenic region of each unit for phylogenetic analyses, while *TSPY* genic sequences alone can only resolve two clusters of *TSPY2* and *TSPY* array genes (**Figure S19A**). It indicates the importance of incorporating longer and more variable flanking sequences of *TSPY* for accurate phylogenetic inferences. PCA and pairwise divergence distributions corroborated this four-subgroup classification (**Figures S19B-C**). Multiple lines of evidence indicated that *TSPY2* experiences stronger functional constraints than array copies. Compared to other array genes, the *TSPY2* shows significantly higher sequence similarity to the bonobo orthologs (median Jaccard similarity of 0.319 vs. 0.306; *W* = 1327052, *P* < 2.22e-16, Wilcoxon rank sum test; **Figure S19D**), but lower (*W =* 11704*, P =* 9.74e-09, Wilcoxon rank sum test) evolutionary rates (measured by the ratio of nonsynonymous vs. synonymous mutation rates, dN/dS ratio; **Figure S20A**) in coding regions. Within populations, *TSPY2* exhibits no CNVs (**Figure 3C**), and substantially lower nucleotide diversity (0.0055 vs. 0.032; *t =* 4.157*, df =* 29*, P =* 0.00013, one-side paired *t* test). Importantly, *TSPY2* exhibits significantly higher testis expression (*t =* 3.943*, df =* 10*, P =* 0.00138, one-side paired *t* test; **Figure S20B**) compared to *TSPY* array genes, specifically in spermatogonia and spermatocyte cells (**Figure S21**). In contrast, the *TSPY* array shows extensive CNVs (especially of *TSPY_B/C* subgroups; **Figure 3C**), pronounced enrichment of coding sequence polymorphisms (**Figure 3D**), and even greater variations in intergenic and *FAM197Y* genes (nucleotide diversity: 0.0426 vs. 0.00207; *t* = 4.619, *df* = 59, *P* = 1.068e-05, one-side paired *t* test; **Figure S22**). These results imply *TSPY2* plays a more constrained role in spermatogenesis, while tandem array copies can tolerate greater structural and sequence variations.

Within the *TSPY* tandem array, functional diversification is evident through subgroup-specific mutations and dynamic evolutionary processes. Each individual carries one *TSPY* pseudogene (*TSPY_ψ*) clustered within the *TSPY_C* subgroup (**Figure 3B**), yet individuals from several haplogroups (C2e, F2 and O2a) have apparently restored this pseudogene to functional status through reverse mutation of the premature stop codon (G>A; **Figure 3D**), possibly via gene conversion with functional copies. There are also widespread gene conversions between array subgroups, with 18 of 61 common (Minor Allele Frequency (MAF) > 0.05) variant sites relative to the *TSPY2* reference that are shared between subgroup pairs, explaining the former difficulty of resolving the array gene identity using *TSPY* genic sequences alone (**Figure 19A**). Nevertheless, we identified some subgroup-specific SVs with potential functional consequences: an 18bp-duplication in the first exon of *TSPY* occurs with a much higher frequency in *TSPY_C* copies (45.2%) than other subgroups (15.7%; **Figure 3D**), and is predicted to significantly alter the protein structure (**Figure S23**). An 89bp-duplication in *FAM197Y* promoter (allele frequency 77.4%) was identified among the *TSPY_B* and *C* but not *TSPY_A* copies. Since this duplication also exists within the Y-linked *FAM197Y* homologs of chimpanzee, bonobo and gorilla, it likely originated in the ancestor of Homininae and became specifically lost in the *TSPY_A* subgroup (**Figure S24**).

Besides the subgroup-level variations, we identified some haplogroup-specific mutations with potential functional impacts. All examined O2a individuals, representing a haplogroup dominant in East Asia, carry a fixed nonsynonymous substitution in *TSPY2* coding region that is predicted to alter protein folding (change of the folding free energy, ΔΔG = −1.39 kcal/mol) (**Figure 3D**). Protein-protein interaction modeling revealed that this mutation modified the TSPY2 interaction affinity (**Figure S25**) with *AR* (Androgen receptor)^47,48^, *ERG* (ETS-related gene)^49^, and *FOXA1* (Forkhead box protein A1)^50,51^, which together constitute a core regulatory axis frequently dysregulated in prostate tumorigenesis. While these computational predictions require experimental validation, they suggest this O2a-specific *TSPY* variant may influence prostate cancer susceptibility or other androgen-regulated processes.

Overall, our evolutionary reconstruction revealed functional stratification of the *TSPY/FAM197Y* gene family since the Homininae ancestor (**Figures 3E** and **S26**), with *TSPY2* being much more conserved as a likely essential spermatogenesis regulator while the tandem array diversified into three subgroups with distinct structural variants and haplogroup-specific mutations. This diversification provides testable candidates for linking Y chromosome variation with male fertility and other male-specific diseases susceptibility across populations in future.

### Large recurrent inversion togglings in the Y chromosome ampliconic regions

While CNVs of *DYZ* satellites and *TSPY* genes account for dramatic Y chromosome length variation (**Figure 1B**), recurrent inversion toggling comprises the other major type of SV affecting ampliconic regions. Previous studies estimated that inversions impact 6% of the Y chromosome length by Strand-seq^18^ and influenced 54.6% of the MSY euchromatin region based on the study of 43Ys^7^, with however the large amplicon-rich AMPL7 region remained undercharacterised. For AMPL1-AMPL6, inversions can be readily identified by assembly sequence alignments because the orientations of the spacer sequences separating palindromes are unambiguous. Using this strategy, we identified the same set of recurrent inversions in previous work^7^, and revealed haplogroup specific stratification (**Figures 4A-B**). In contrast, the highly similar nested palindromic organization of AMPL7 (illustrated by the colored arrows in **Figure 1C**) precludes reliable inference by conventional alignment-based approaches. To address this challenge, we developed a sub-amplicon synteny based method that infers SVs without requiring whole-genome sequence alignments. This approach enabled the identification of eight previously uncharacterized large (>10 Kb) inversions within AMPL7 (**Figures 4D**). Together with the recurrent inversions identified in AMPL1-AMPL6, we identified a total of 24 large inversions across Y-linked euchromatic regions (**Tables S10** and **S14**). Nearly all inversions except one are located within the AMPL regions, whereas the remaining euchromatic regions exhibit remarkably high synteny across haplogroups (**Figures 4A** and **S27**).

**Figure 4.**
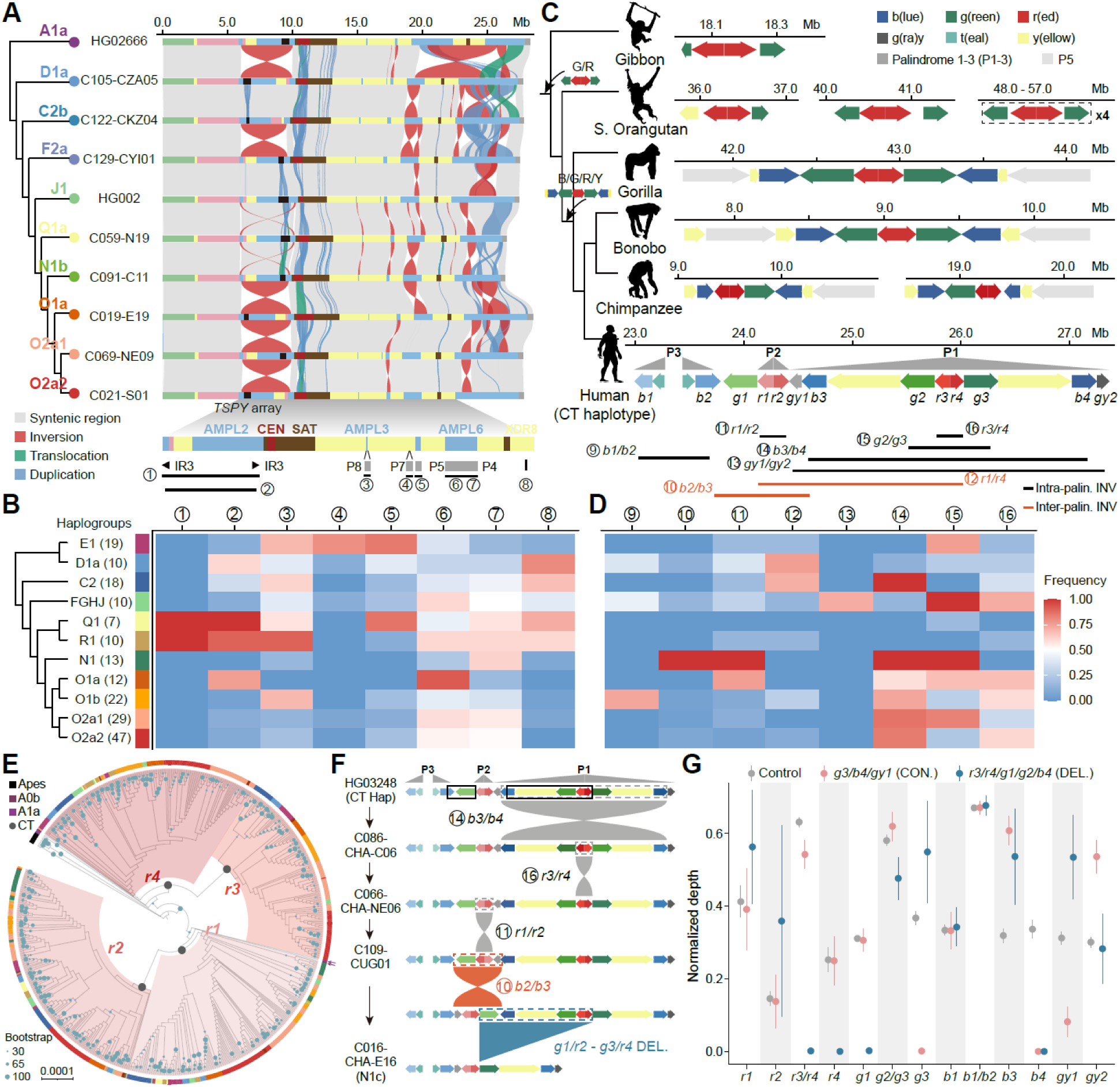
Large recurrent inversions in the Y chromosome ampliconic regions. **A)** Synteny of euchromatic Y-chromosome regions between individuals of major haplogroups, with colored lines between them showing different types of SVs (red for inversions, green for translocations, blue for duplications, and grey for syntenic regions). Horizontal colored bars denote the annotated Y-linked subregions according to Figure 1b, and black bars mark the *TSPY/FAM197Y* gene array. The red cross lines across AMPL1 and AMPL2 between individuals of J1, O1a and N1b haplogroups indicate two nested inversions (inversion 1 and 2). **B)** Frequencies of identified large inversions (> 10Kb, inversions 1-8) in the euchromatic regions (excluding AMPL7) within major haplogroups using the HG002 Y chromosomal sequence as reference. **C)** Evolution of the composition and synteny of amplicons of AMPL7 among great apes. Red and green amplicons are conserved across all apes, whereas blue amplicons originated in the common ancestor of Hominidae. In humans, additional amplicons were subsequently incorporated and rearranged to form three large palindromes. Four amplicons, red, green, blue and gray, for all CT haplogroups were further classified into two to four subgroups and numbered according to their order along the CT ancestral haplotype shown for the human. Inversions in AMPL7 were further inferred by comparing the synteny of the classified amplicon units against the CT haplotype. Two types of inversions, inversions within palindromes or between palindromes were distinguished by different colors. Only inversions with the frequency among all examined individuals of at least one haplogroup higher than 0.5 were shown for both heatmaps (**B** and **D**). **E)** Phylogenetic trees of red and green amplicons across Y haplogroups with well-annotated amplicons in AMPL7. Homologous sequences of the ape Y chromosomes were used as outgroups. The size of blue dots is scaled to the bootstrap support, and the gray dots mark the CT node within each subgroup. The color arrows represent the amplicons from A0/A1 individuals. **F)** The mutational trajectory of the common *b2/b3* Y-linked deletion, whose affected region is shown by the black box on the CT haplotype (from the sampled individual HG03248 in this work). The actual order of three intra-palindrome inversions can be changed. **G)** Normalized short-read depth of k-mers (31-mer) specific to each subgroup of colored amplicons. The X-axis indicates the amplicon subgroups defined from Figure 4E, whose depths of diagnostic k-mers were counted and normalized by the average read coverage of the XDR region. Three AMPL7 haplotypes were included to demonstrate the resolution for gene conversion and micro-deletion events by short-reads only: the Control haplotype without any detected gene conversions or micro-deletions; the gene conversion (CON.) haplotype that carries gene conversions of *g3>g2*, *b4>b3*, and *gy1>gy2*, predominantly observed in the C2 haplogroup (**Table S12**); and the deletion (DEL.) haplotype that was characterized by deletions affecting *r3/r4/g1/g2/b4,* predominantly in N1 haplogroups^53^. A lack of diagnostic k-mers for certain amplicon subgroups indicates gene conversions or structural variations. The elevated depth of *r1/r2/g3/gy1* in the DEL. group is attributable to an additional duplication in N1c, which cannot be estimated accurately by short-read depth alone.

The complexity of AMPL7 structure stems from its evolutionary history. Cross-species comparisons revealed that inverted pairs of red and green amplicons^52^ originated in the ancestor of apes, while blue and partial yellow amplicon pairs were acquired among great apes (**Figure 4C**), followed by human-specific duplication and rearrangements (**Figure S28**). These duplications expanded the AMPL7 specifically in humans, and also posed a tremendous analytical challenge due to the nearly identical sequences of amplicons from the same color family. To circumvent this, we classified most of the same-color amplicons, except for the yellow and gray ones (**Figure S29**), into two to four distinct subgroups (**Table S11**) based on their phylogenetic (**Figure S30**) and PCA clustering (**Figure S31**; **METHODS**). For example, red amplicons can be divided into four subgroups (*r1-r4*) that show different testis expression level for their embedded *DAZ* gene copies (**Figure S32**), consistent with their non-random pattern of deletions in patients with spermatogenic arrest^53,54^. As expected from the result of gene conversions, amplicons within the same palindrome (e.g., *r1* and *r2* units of P2) are more closely clustered to each other than to those in different palindromes (**Figure 4E**). Within each subgroup, most amplicons from the same haplogroup are also expectedly clustered together (**Figures 4E** and **S30**). Notably, amplicons from the African haplogroups, A0b and A1a, which represent the oldest known haplogroup diverged over 0.2 million years ago^28^, form a separate phylogenetic branch from the major ‘out-of-Africa’ modern human lineages (**Figures 4E** and **S30**), suggesting either distinct evolution trajectories or lineage-specific gene conversion events of AMPL7 between the basal vs. derived modern human populations separated by the ‘out-of-Africa’ dispersal. However, we are currently restricted to very limited available Y chromosomal sequence for the A0, A1 and B haplogroups; more individuals from these old haplogroups are needed to confirm this hypothesis.

Distinguishing the same-colored amplicons from one another per individual enabled us to use them as markers to characterise SVs within AMPL7, without relying on sequence alignments between individuals that can be readily confounded by cross-mapping and other types of SVs within AMPL7 (e.g., translocations and duplications in **Figure 4A**). It led to several important discoveries: first, we cataloged 175 individuals (**Table S11**) with well-annotated amplicons in AMPL7 into 76 distinct haplotypes (**Tables S12-13**), based on the copy numbers and the synteny, i.e., relative orientations, of subgroups of amplicons. The breadth of haplogroup sampling allowed us to reconstruct the ancestral AMPL7 haplotype configuration of the modern human populations, shared by the predominant ‘CT’ haplogroup that emerged after divergence from ancient A0/A1 haplogroups^28,55^ (**Figure S33A**; **METHODS**). This ancestral CT haplotype of AMPL7 provided a better reference for identifying inversions across individuals than the HG002-Y (**Figure 4E**), as the latter of which has lost the ancestral palindromic structure of P1 through multiple inversions (**Figure S34**). Second, we discovered that these newly identified inversions (with their breakpoint intervals annotated as the size of certain amplicons) (**Figure 4D**), and micro-deletions and duplications of amplicon units (**Figure S35**) emerged recurrently across different haplogroups (**Table S14**; **Figures S33B-C**). They expanded the catalogs of both common and rare structural mutations within AMPL7^7,18^, and provided an important reference for future molecular diagnosis with low-cost short reads (see below). To validate these rearrangements, we developed an SNP/Indel marker based strategy that does not rely on local sequence alignments or predefined amplicon subtypes (**Figure S36A**; **METHODS**). Amplicons of the same color class from all individuals were projected onto a common reference sequence to identify markers. Orientation-aware similarity dot-plots generated from these marker chains consistently identified the SVs inferred from amplicon synteny (**Figures S36B-C**), e.g., the recurrent *b2-b4* and *gr-gr* inversions as well as associated microdeletions, while providing improved breakpoint resolution (**Figures S36-39** and **Supplementary materials**). Notably, breakpoint positions varied among individuals carrying the same class of inversions, highlighting the structural complexity of the AMPL7 region. Finally, 16 of the 24 inversions (66.6%, shown in **Figures 4B-D**) independently reached high-frequency (> 50%) within specific haplogroups (**Table S14**), demonstrating extensive inversion togglings during the evolution of Y chromosome euchromatin region.

These recurrent inversion togglings likely arise by NAHR between the abundant inverted repeats or palindromes. For example, the two inversions spanning AMPL1 and AMPL2 were found to be present among all individuals of Q1 and R1 haplogroups with slightly different breakpoints that overlap with the inverted repeats IR3 (**Figures 4B** and **S40**). In contrast to autosomes, the impacts of inversion toggling on the already-nonrecombining Y chromosome are unclear, especially for most inversions within AMPL7 that do not disrupt the protein-coding regions. These inversions may promote subsequent morbid deletions and duplications by increasing the genome instability at the breakpoint regions^18,56^. Such impacts seem to be more pronounced for inversions between palindromes than those within palindromes of AMPL7, possibly due to disruptions of pre-existing palindromic structure and formation of new pairs of direct repeats (*g1* & *g3*; **Figure 4F**) by the former. Indeed, individuals with both inter- and intra-palindrome inversions showed 5.9-fold more amplicon deletions than those with only intra-palindrome inversions (*χ²* = 32.967, *df* = 1, *P* = 2.57e-08, Chi-squared test; **Figure S41**). The N1 haplogroup exemplifies this mutational cascade. Reconstruction of its evolutionary trajectory from the ancestral CT haplotype revealed multiple intra-palindrome inversions followed by the *b2*/*b3* inter-palindrome inversion and microdeletion that gave rise to its extant haplotype (**Figures 4F** and **S33C**).

The *b2/b3* microdeletion, present in 1.1% of the global population^24,57^, results in partial *AZFc* deletion and remains highly controversial regarding its impact on spermatogenic failure, with effects appearing haplogroup-dependent^53,57^. One probable cause of such debate has been the challenge to precisely characterize which specific amplicons are deleted in the individual patient. The classic STS (Sequence-Tagged Site) method cannot identify the exact deleted region, while our results revealed there are substantial variations of the boundaries of SVs between healthy individuals. We applied a k-mer based strategy which has been widely used in genomic genotyping or association studies^58,59^, based on our classification of amplicon subgroups (**Figure S42**) to enable cost-effective, precise identification of which specific subgroup of amplicons are deleted or have undergone gene conversion using only short-read sequencing data, without requiring expensive long-read sequencing or assembly. For example, for the individuals whose Y chromosomal assemblies were characterised by deletions of certain subgroups of amplicons (e.g., *r3/r4/g1/g2/b4*), we indeed cannot detect any diagnostic k-mers of the specific subgroups of amplicons from short-reads (on average 30x sequencing coverage) of the examined samples (**Figure 4G**), using the HG002-Y as reference. Interestingly, lack of diagnostic k-mers of certain amplicons can be also the result of gene conversion (e.g., in C2 haplogroups; **Figure S33**), for which we also managed to verify by the doubling of normalized k-mer abundance of another amplicon of the same color (**Figure 4G**).

This diagnostic k-mers approach also provided critical validation for the evolutionary divergence of AMPL7 during the out-of-Africa dispersal. To test whether the unique phylogenetic branching of the African A0b/A1a amplicons represented genuine evolutionary history rather than artifacts of low numbers of samples(**Figure 4E**), we analyzed five additional A0b/A1a individuals with short-read data available^60^. This confirmed the complete absence of *r2* and *b4* amplicon-specific k-mers in all A0b/A1a individuals (**Figure S43**), in contrast to the consistent presence among all examined CT haplogroup individuals. This suggests that red and blue amplicons either underwent sequence divergence specifically during the out-of-Africa migration, creating the distinct subtypes characterizing modern Eurasian populations, or experienced lineage-specific gene conversion within the ancestral African branches.

### Candidate Y chromosomal disease-associated structural variations

After identifying new molecular diagnostic markers of amplicons, we finally ask whether the newly assembled chromosomal sequences across haplogroups can help to recover the disease-associated genetic SVs that were previously missed due to the incomplete assembly of MSY regions. As a pilot study, we first focused on prostate cancer (PCa), the second most common cancer in men with reported increased risks associated with Y-linked mutations^20,61–63^. To investigate the somatic dynamics of Y-linked multi-copy arrays during PCa progression, we analyzed a matched cohort of 200 PCa and adjacent para-tumor pairs^51^. Using linear mixed-effects models (LMMs) adjusted for key clinical covariates, including patient age and Gleason score (a histopathological grading system reflecting PCa aggressiveness), we identified a significant reduction in *TSPY* copy number specifically in haplogroup O1a tumors relative to matched para-tumor tissues (32.5 vs. 33.8, FDR-adjusted *P* = 0.00389; **Figures 5A** and **S44; Table S15**). To determine whether this O1a-specific *TSPY* reduction was an isolated locus event or indicative of a broader structural instability on the Y chromosome, we extended our LMM framework to the massive DYZ1 and DYZ2 satellite arrays. Concordantly, haplogroup O1a displayed concurrently significant copy reduction in both DYZ1 (estimated reduction of 506 copies, FDR-adjusted *P* = 0.00117) and DYZ2 arrays (estimated reduction of 691 copies, FDR-adjusted *P* < 0.0001; **Figure 5B**). Similar profound structural collapses of DYZ arrays were robustly detected in other expanded haplogroups, including C2e, O2a1 and O2a2 (FDR-adjusted *P* < 0.001; **Figure S45**). This parallel collapse of *TSPY* and DYZ arrays exclusively within specific genetic backgrounds reflects genuine haplogroup-modulated somatic variations within the prostate tumor. While this copy reduction may partially reflect generalized tumor genomic instability, we speculate that localized destabilization of *TSPY* array and Yq12 heterochromatin may actively contribute to prostate pathogenesis. This might be associated with disordered ribosome biogenesis, as integration of PCa transcriptomic data revealed that genes whose expression levels correlated with somatic DYZ copy alterations were highly enriched in ribosomal function (**Table S16**), consistent with a previous study of HSat3 arrays participating in ribosomal DNA regulation^64^.

**Figure 5.**
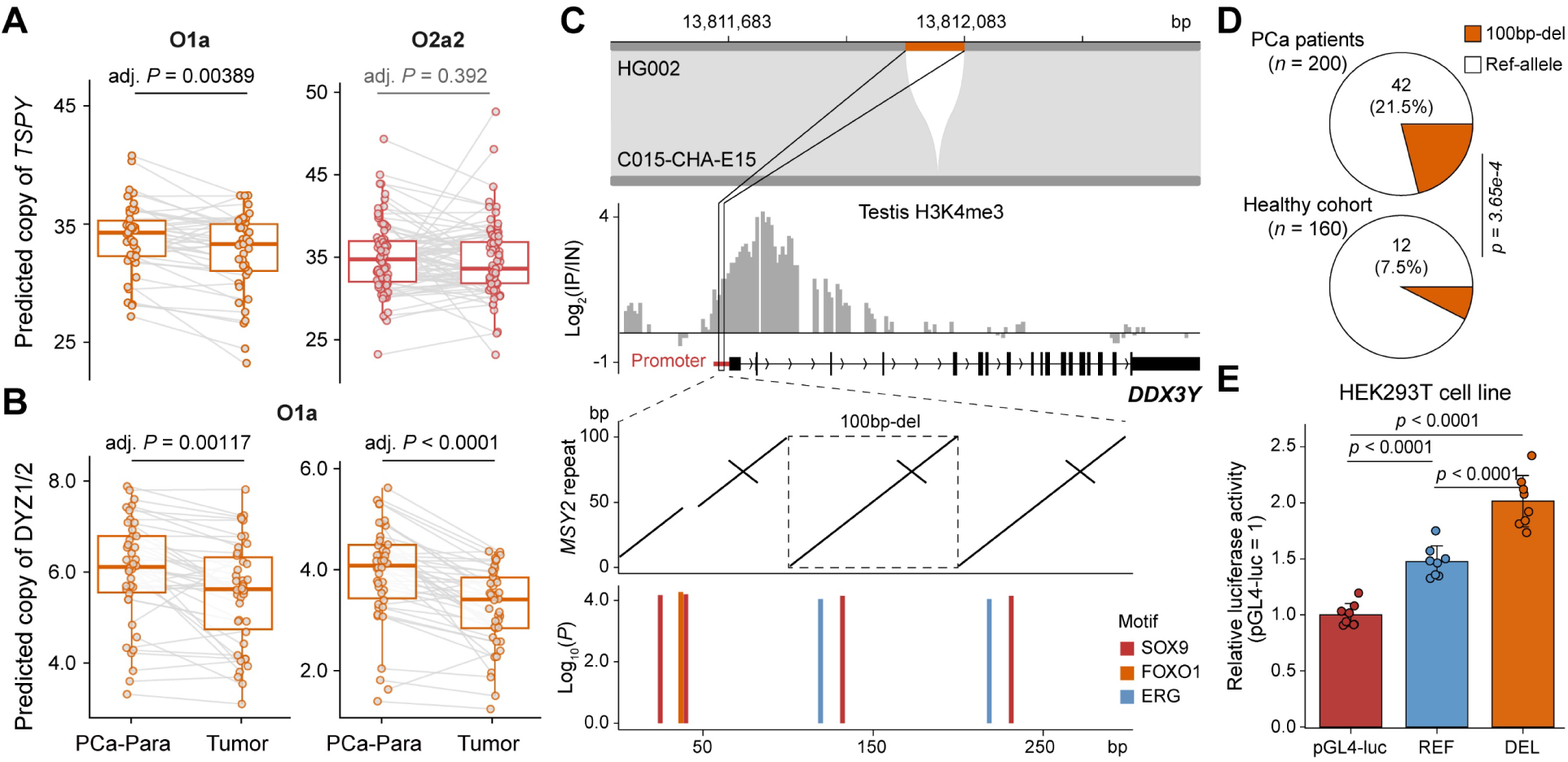
Candidate Y-linked prostate cancer related variants. **A)** The comparison of short-read estimated copy number (CN) for *TSPY* gene among healthy cohort in this study, prostate cancer (PCa-Tumor) and adjacent normal (clinical) tissue (PCa-Para) for O1a and O2a2 haplogroups. **B)** A higher DYZ1/2 copy number in Pca-para compared to Pca-tumor samples. **C)** A 100bp deletion in the promoter of the *DDX3Y* gene. This deleted segment is part of a tandem duplication unit known as the *‘MSY2’* repeat^80^ and is predicted to harbor binding motifs for multiple important transcription factors. **D)** This deletion shows a higher proportion among individuals with prostate cancer (*n* = 200) compared to the healthy cohort (*n* = 160). **E)** The x-axis shows the control construct (pGL4-luc), the 100-bp deletion allele (200-bp sequence lacking the deleted fragment), and the reference allele (300-bp sequence containing the deleted fragment). The y-axis indicates normalized luciferase activity. Reporter assays were performed in cultured HEK293T (human embryonic kidney 293T) cells.

Beyond the *TSPY* and DYZ array, numerous variants across other euchromatic regions of the Y chromosome (**Figure S46; METHODS**) exhibit marked differences in frequency among populations and haplogroups. In PAR, we identified four genes *CRLF2* (Cytokine receptor-like factor 2), and *IL3RA* (Interleukin-3 receptor subunit alpha), *SHOX* (Short stature homeobox) (**Figure S47**), *ASMTL* (Acetylserotonin O-methyltransferase-like) harboring SVs (> 50bp) with substantial population differentiation (*Fst* > 0.3; **Figure S48**). *CRLF2* and *IL3RA* are two immune-related genes whose mutations are linked to hematopoietic malignancies such as B-cell acute lymphoblastic leukemia^65–67^. Mutations in *SHOX* were reported to be associated with idiopathic short stature and Léri-Weill dyschondrosteosis^68^, and we found the European population carry a 54bp-deletion in the last intron with a much higher allele frequency than other populations (**Figure S47**). *ASMTL* has been reported to be related to melatonin biosynthesis regulation^69^. Notably, we identified a high-frequency 78bp-deletion specifically in Africans (38.0% vs. 0.0% in other populations; **Figure S49**) within the promoter region that has been demonstrated to significantly downregulate the *ASMTL* expression in lymphocyte cell lines^70^, suggesting a potential regulatory impact.

In MSY, we identified three protein-coding genes *VCY1B* (Variable Charge Y-Linked 1B), *NLGN4Y* (Neuroligin 4 Y-Linked) and *DDX3Y* (DEAD-Box Helicase 3 Y-Linked) with fixed (*Fst* = 1.0) allele differences across multiple haplogroups compared to E1, a basal haplogroup of CT lineage as reference (**Figure S48**). We identified an intronic deletion in *NLGN4Y* fixed in the D1a haplogroup (**Table S17**), the most dominant haplogroups among Tibetans. This gene was implicated to be associated with autism spectrum disorder^71^, and our result is consistent with the previous study^72^ showing this deletion has a high frequency among Tibetans. The *VCY1B* is involved in regulation of ribosome assembly during spermatogenesis^73^ and harbors a fixed 87bp-deletion at 238bp downstream of the gene that has been present since the common ancestor of J/Q/R/N/O haplogroups. Notably, *DDX3Y* has a 100bp-deletion in its promoter in all examined O1a individuals (**Figure S50**). This gene is located in *AZFa*, and was implicated in human non-obstructive azoospermia^74,75^ and prostate cancer progression^76,77^. The deleted interval belongs to the testis-specific transcription initiation sites^78^ of *DDX3Y*, and encompasses predicted binding motifs including several key transcription regulators of testis development and prostate cancer, e.g., *SOX9* (SRY-Box Transcription Factor 9), *FOXO1* (Forkhead Box O1) and *ERG* (**Figure 5C**), suggesting potential regulatory consequences. We further observed that the O1a haplogroup, which carries this fixed promoter deletion, is significantly enriched in the prostate cancer cohort^51^ (21.5% vs. 7.5%, *χ²* = 12.706, *df* =1, *P* = 3.65e-04, Chi-squared test; **Figures 5D** and **S50**) compared to healthy APG cohort, even with populations controls from Eastern China, a region with one of the highest prostate cancer incidence rates in China^79^. To assess the potential functional consequence of this promoter variant, we performed a dual-luciferase reporter assay in HEK293T cells. The construct sequence carrying the derived allele (100-bp deletion) exhibited significantly higher luciferase activity (*t* = 5.892, *df* = 7, *P* = 0.000302, one-side paired *t* test; **Figures 5E** and **S51**; **Table S18**) than the reference allele, indicating that the deletion alters the transcriptional activity of the *DDX3Y* promoter. Nevertheless, the deletion is completely fixed within the O1a haplogroup, its potential contribution to prostate cancer susceptibility or spermatogenesis disorder cannot be disentangled from the underlying haplogroup background. Further studies using larger and geographically matched cohorts will be required to determine whether this haplogroup-specific structural variant contributes to disease risk.

Together, these newly identified haplogroup-specific SVs provide targets for future functional studies investigating how Y chromosome variation contributes to population differences in disease susceptibility.

## DISCUSSION

The Y chromosome has historically been underrepresented in genome sequencing and population genomic analyses^81^, largely because its long, complex and repetitive regions posed exceptional challenges for assembly using Sanger and short-read sequencing technologies. As a result, the human Y chromosome was the last chromosome to be completely resolved^6,44^. Recent advances in long-read sequencing and Hi-C technologies have transformed this landscape, generating complete or near-complete Y chromosome assemblies for humans and other great apes^6–8^ and substantially expanding our understanding of Y chromosome structure, evolution and functional content. However, the number of gapless Y chromosome assemblies remains limited relative to the extensive diversity of human Y haplogroups, leaving many population-specific structural variants and evolutionary patterns insufficiently characterized. Here we present a comprehensive population-scale view of Y chromosome structural variation through high-quality assemblies spanning major human haplogroups. This expanded dataset reveals extensive cryptic diversity within human Y chromosome’s most repetitive regions, including ancient sequence variants, lineage-specific structural rearrangements, and population-stratified SVs with profound implications for understanding both evolutionary dynamics and disease associations.

We traced the evolutionary origins of the repeated units or composite structures of Yq12, *TSPY* family and AMPL7 to their ancestral sequences shared with other apes (**Figures 2E**, **3E** and **4C**). We focused on these regions not only because they represent the largest repetitive structures in the human genome and were more fragmented in previous assemblies, but also because similar complex regions appear throughout the human genome (e.g., centromere satellites and rDNA) and sex chromosomes of other vertebrate species (e.g., palindromes^14,82,83^). Dissecting their sequence and structural heterogeneity across human populations since their origins from the ape ancestor therefore bears general implications for the mechanisms of transient sequence and functional evolution, before the expected complete degeneration in the absence of recombination^84^.

Our analysis revealed that the alternating DYZ satellite unit, which specifically originated and expanded in humans to form Yq12, are derived from ancestral satellites of the same family in centromeric regions of acrocentric chromosomes shared across apes (**Figure 2E**). This evolutionary connection likely explains why 70% of *de novo* Y/autosome translocations involve Yq12 translocations onto acrocentric chromosomes, with few having a phenotypic impact^85–87^. It also accounts for the spatial proximity of Y-linked heterochromatin to nuclear organiser regions (NORs) on acrocentric chromosomes during interphase^88^. This nuclear architecture association may contribute to the reported Y-linked regulatory variation (YRV), where Y chromosomal heterochromatin is assumed to impact global gene expression levels through competing chromatin modulators with other chromosomes (the ‘heterochromatin sink’ hypothesis)^89^. In *Drosophila*, males with different Y chromosomes but identical genetic backgrounds elsewhere in the genome show differential expression levels of autosomes and X-linked genes, demonstrating that Y heterochromatin variation can influence genome-wide transcription^90–92^. However, evidence for YRV in humans remains mixed. One study of XYY males found increased overall expression of transposable elements in blood^93^, while another recent study in lymphoblastoid and fibroblast cell lines found no evidence of heterochromatin sink^94^. Our discovery that Yq12 encodes numerous lnRNAs and piRNAs suggests an additional source of YRV, particularly in testis (**Figures 2G** and **2H)**. Furthermore, DYZ1 satellites are predicted to be enriched for binding motifs of *TEAD1/4* of the YAP/TEAD signaling pathway^64^ and prostate cancer related TFs (**Figure S52**), including *HSF1/4*^95^*, EVX1*^96^ and *Nfact2*^97,98^. The functional consequences of the drastically different copy numbers of DYZ satellites between individuals that either encode non-coding RNAs or provide binding sites of TFs are to be examined in future.

Although massive tandem and inverted repeats on the Y chromosome were thought to maintain highly homogenized sequences through gene conversions^4,82^, a critical finding of this work is the classification of DYZ, *TSPY* and amplicon units into distinct subgroups with different evolutionary ages (**Figures 2B**, **3B** and **4E**). This classification revealed their non-uniform chromosomal distribution and, more importantly, suggests potentially different functional significance of these highly similar units, whose mutations likely have different pathogenic effects. For example, *DYZ* copy numbers are far more conserved near Yq12 boundaries than within the central region (**Figure 2F**), paralleling the pattern for *TSPY2* vs. *TSPY* tandem arrays (**Figure 3C**). The discovery of inverted array pairs flanking Yq12 boundaries provides new mechanistic insights into how boundaries of such a large heterochromatin region are defined, not through canonical chromatin insulators but through specific satellite sequence arrangement^7^. Further studies of male infertile or cancer patients should examine whether mutations in these boundary-specific units occur more frequently than in their internal counterparts. Overall, our analyses of the completed Y chromosomal sequences of human populations revealed their evolution history and cryptic divergence among large numbers of repeats that provide a more comprehensive understanding of Y chromosomal functions. As precision medicine increasingly recognizes the importance of population-specific genetic architecture, the Y chromosome can no longer be dismissed as a genetic wasteland but must be integrated into our understanding of human health and disease.

## CODE AND DATA AVAILABILITY

The workflow of assembly and annotation for all 160 Y chromosomes are available at Github (https://github.com/Asian-Pan-Genome/APGp1). All related code scripts have been deposited at GitHub (https://github.com/Asian-Pan-Genome/APGp1-Y). The assemblies and raw sequencing data for all Y chromosomes generated in this study are part of the APGp1 (Asian Pan-Genome phase1) project and have been deposited in the National Genomics Data Center (https://ngdc.cncb.ac.cn) under BioProject accession PRJCA030428. Detailed information is provided in the *Data Availability* section of the APGp1 flagship manuscript (Wu et al. 2026). The public datasets, including reference genomes, Y assemblies, Iso-seq, bulk RNA-seq, scRNA-seq and prostate cancer cohort, were summarized in **Table S21**. Other supplementary materials were available on GitHub.

## ACKNOWLEDGMENTS

We thank all members of APGp1 project and sampling participants related to this project. This work was funded by the National Key Research and Development Program of China (2024YFA1802500 and 2023YFA1800500), and National Natural Science Foundation of China (73432170415) to Q.Z., the International Institutes of Medicine of Zhejiang University (KY2022-098), National Key Research and Development Program of China (2024YFA1802500), Basic Research Center Program (32388102), the New Cornerstone Science Foundation through the XPLORER PRIZE and the K.C.Wong Education Foundation to G.Z..

## AUTHOR CONTRIBUTIONS

Project design: Q.Z., G.Z.; Project supervision: Q.Z., G.Z., D.W.; Genome assembly, polish and evaluation: D.W., C.Y., J.L., Q.N., Q.C.; Genomic analysis: J.L., Q.N., Y.Z. Q.C.; Manuscript writing and revision: Q.Z., G.Z., J.L., Q.N.. All authors have reviewed and approved the manuscript.

## METHODS

### Y chromosome assembly and evaluation

The primary assemblies of 160 Y chromosomes were derived from the assembly effort of APGp1 (Asian Pan-Genome phase1) project^31^ (**Table S1**). In brief, Hifiasm (v0.16.1)^97^ and Verkko (v1.4.1)^98^ with trio-mode were used to generate primary contigs, with Verkko-derived contigs being used to improve the continuity of Hifiasm-derived contigs^31^. To ensure the high integrity and accuracy of Y assemblies, we manually examined all potential mis-assemblies, including abnormally short (e.g., over 1Mb shorter than that of HG002-Y) PAR1 (pseudoautosomal region 1) (3/160), misplacement of PAR2 fragments (12/160), and heterochromatic regions that were severely (e.g., over 10Mb shorter than the contig size of Hifiasm or Verkko primary assembly) shortened or incorrectly assigned for maternal haplotypes (Yq12; 15/160) (**Table S3**). These issues were largely attributed to unusual genomic features, such as the high level of GA-microsatellite density in PAR1 and that of GC-content variation in Yq12^5^, both of which are known to introduce bias in Pacific Biosciences (PacBio) high-fidelity reads (HiFi) sequencing^99^. Mis-assemblies involving PAR2 were primarily caused by mis-joined contigs that included tandem repeats of Yq12. In such cases, instead of using complementary contigs from Hifiasm and Verkko that might introduce unexpected errors, we selected and connected the longer contigs generated by either with RagTag (v2.1.0)^100^ with the HG002-Y^5^ genome as reference. All the draft assemblies were then projected to correct structural variants (SVs; at least 50bp long) identified from both phased ONT and HiFi reads using Sniffles2^101^. Subsequently, base-level accuracy was enhanced through two rounds of NextPolish2 (v0.2.1)^102^ using PacBio HiFi alignments and short-read derived k-mers processed by yak (v0.1-r69) (https://github.com/lh3/yak).

The size of Y chromosomes were estimated by mapping the HiFi and PCR-free (MGISEQ) short reads to the HG002-Y reference and extracting Y-linked reads using SAMtools (v1.9)^103^, following by size estimation using GenomeScope2.0^104^ with settings: -k 21 -p 1. Size estimates from HiFi and short reads were compared with the assembly sizes across 85 and 75 samples without/with assembly gaps (**Figure S53**). Given the challenges of mapping and assembly in complex regions such as PARs, (peri)centromeres, and Yq12, our focus was on evaluating the correlations between estimated and assembled sizes rather than their absolute concordance, as a measurement of assembly integrity. The strong correlation (short-reads: *R*^2^ = 0.63, *P* = 4.42e-19; HiFi reads: *R*^2^ = 0.65, *P* = 1.96e-20) between the assembled and predicted sizes (**Figure S53**) supported the completeness of our assemblies. Although assembly gaps can impact the size of Yq12, no significant size differences (*P* > 0.05) were observed between gapped and gapless samples (**Table S4**).

To assess the base-level accuracy and structural continuity of the resulting Y chromosome assemblies, we calculated the scores of quality value (QV)^35^ and Genome Continuity Inspector (GCI v1.0)^105^ (**Table S2**). QV scores estimate the consensus base accuracy from HiFi and short reads, and are calculated using Merqury (v1.3)^106^. Briefly, hybrid k-mer (k = 21) spectrum were constructed from both MGI and HiFi reads using Meryl (v1.4.1), followed by union-sum merging and default execution. The assembly integrity were assessed using GCI which integrates both binned HiFi and Oxford Nanopore ultralong reads (ONT) read alignments generated via Minimap2 (v2.26)^107^ and Winnowmap2 (v2.03)^108^ to detect structural inconsistencies and report base-level continuity..

To identify potential mis-assemblies, we examined the nucleotide-level discordance using NucFreq (v0.1)^109^, and the structural assembly using Flagger (v0.3.2)^110^ (**Table S5**). For NucFreq, the alignments of binned HiFi reads were filtered using SAMtools (v1.9) with parameters -bh -F 2308 to perform secondary and supplementary alignments. Nucleotide frequency profiling was generated using NucPlot.py with default settings. Genomic regions were flagged when ≥10% of reads supported for the second most frequent base at ≥5 positions within each 500bp sliding window, as determined by hetDetection.R^111^. Flagger classifies the potential structural assembly mistakes into categories of being collapsed, duplicated, or erroneous. To correct for HiFi platform-specific sequencing biases in satellite-rich regions^110^, coverage thresholds were adjusted to 0.55 for Revio and 1.2 for Sequel II in HSat2/3 arrays. To integrate the results of both NucFreq and Flagger, BEDtools (v2.31.0)^112^ was used to compute their intersections and unions (**Table S5**). The more likely mis-assembly regions (the intersected regions) account for a small proportion (mean: 0.085%), and the union flagged regions accounted for a median of 1.32% (ranging from 0.17 to 29.0%), with eight samples exhibiting an estimated mis-assembly rates above 10%, mainly because of the highly fragmented assembly of Yq12. In fact, the Yq12 region comprises the highest proportion (52.1%) of collapsed errors and biased read coverage (**Figure S3**), consistent with its complex and long repeated structures^3,5^. Elevated error rates were also observed in two additional complex regions: AMPL7 (ampliconic region 7), which comprises six palindromic amplicon families^51^, and centromeres with higher-order satellite repeat (HOR) arrays^41^. Assembly quality in complex regions (e.g., centromere (CEN) and Yq12) was further assessed using VerityMap (v2.1.2)^113^ with HiFi reads in ‘-d hifi-diploid’ mode, and GAVISUNK (v1.0.0)^114^ using the ONT reads under default parameters (SUNK_len = 20) (**Figure S54**). Several large and rare (singleton) rearrangements were validated using HiFi and ONT read depth and synteny analysis against the primary contigs to rule out assembly artifacts (**Figure S55**).

Phasing accuracy in the PARs of chromosomes X and Y was assessed using yak (v0.1-r69). Parental 21-mer databases were first constructed from short-reads using ‘yak count’. Switch and hamming error rates were then calculated with ‘yak trioeval’ for PAR sequences. We applied the VGP (Vertebrate Genome Project) telomere detection pipeline^35^ to identify telomere-associated repeat motifs on the Y chromosomes, retaining only motifs located within 1Kb regions at both chromosomal ends. About 88.8% of Y-chromosomal ends can be annotated for telomeres, supporting the integrity of our assemblies (**Table S19**).

### Y chromosome annotations

Gene annotations were transferred from that of the HG002-Y using Liftoff (v1.6.3)^115^ with the following parameters: -sc 0.95 -copies -polish -exclude_partial. Premature stop codons and frameshift mutations were curated in each assembly to identify pseudogenes, especially in the multicopy Y-linked gene families (**Figure S2**).

Each Y chromosome assembly was firstly soft-masked using RepeatMasker (v4.1.2; Dfam 3.3) to reduce the alignment bias of repetitive sequences. The primary Y chromosome subregion annotations were generated using nf-LO (v1.8.0)^116^ with Minimap2 (v2.26) alignment to the HG002-Y reference to produce a preliminary chain file for each individual. The resulting chain files were refined with chain splitting, PAF format conversion, and alignment filtering following previous workflow^43^. Filtered alignments were then proceeded to convert subregion coordinates from the annotation of HG002-Y using CrossMap (v0.7.4 ^117^. Unmapped and redundant entries were removed, and final annotations were consolidated with custom scripts. Further manual inspection and trimming via alignment visualization were performed to ensure consistency and accuracy. Self-alignments and alignments to HG002-Y were both conducted using Lastz (v1.04.00; https://github.com/lastz/lastz). For self-alignments, initial ungapped anchors were first identified (--ungapped --filter=identity:80 --filter=nmatch:400 -- hspthresh=36400), followed by alignments filtered at ≥80% identity and ≥1000 matched bases. Strand orientation and boundary artifacts were corrected, and dot-plots were generated using adapted EDFig3a_dotplot_idy.R^5^ for visualizing structural features, detecting potential mis-assemblies and correcting the coordinates of subregion boundaries. The subregion boundaries were further manually refined for the Y chromosomes which exhibit large inversions or deletions.

The boundaries of PAR1 and PAR2 on the X and Y chromosomes were refined by pairwise alignments with the HG002-Y using Lastz (v1.04.00) for each sample. For the PAR1, we further analyzed the distribution of single-nucleotide polymorphisms (SNPs), small deletions/insertions (INDELs; < 50bp) and structural variants (SV deletions/insertions; ≥ 50bp) based on the Minigraph-Cactus graph pangenome^17,29^ (see below). Variants located within the first 3Mb of HG002-Y were extracted, and their densities were computed in non-overlapping 10Kb windows to evaluate variation patterns around the canonical PAR1 boundary^30^. Consistent with previous findings^31^, a reduction in SV polymorphism was observed within the ∼500Kb from the canonical PAR1 boundary, which however, cannot exclude the possibility of ongoing X-Y recombination, as also highlighted by Bellott and colleagues^32^. To explore potential XY genetic exchanges at a finer resolution, we examined the shared SNPs and SVs between X and Y chromosomes (**Figure S8**). Specifically, the 1Kb regions flanking the canonical PAR1 boundary (± 500bp) were aligned using MAFFT (v7.505)^33^. Following the logic of Bellott et al.^32^, we defined the PAR1 boundary as a precise 116-bp interval. This interval is anchored between the last shared derived SNP located just distal to the canonical PAR1 boundary and the first proximal fixed difference.

Inverted repeats (IRs), major palindromes (P1-P8), and six amplicon families (red, green, blue, yellow, teal and gray) located within the AMPL7 were initially identified by converting the GRCh38-Y annotations^51^. Specifically, the amplicon sequences of GRCh38-Y were aligned to each Y chromosomal assembly using Winnowmap2 (v2.03). The resulting alignments were converted to BED format using BEDTools (v2.31.0), merged and refined with custom scripts to resolve overlaps and extend region boundaries. The coordinates of IRs and palindromes were further refined using Palindrover (https://github.com/makovalab-psu/T2T_primate_XY)^34^, which identified the palindromic structures based on ≥ 98% sequence identity and ≥ 2Kb length with Lastz (v1.04.00) based alignments. Given the structural complexity of the six colored amplicon families within the AMPL7 region, dot-plots were generated for both self-alignments of 160Y chromosomes and pairwise comparisons with HG002-Y. Amplicon boundaries were manually refined based on these alignments, and internal breakpoints of associated palindromic structures were identified to ensure consistent structural annotation (**Figure S2**).

As DYZ1 is known to belong to the HSat3 subfamily 6A^41^, we identified the DYZ1 blocks and their HSat3 (Human Satellite 3) homologs across the genomes of 160Ys and great apes^34,35^ by searching for HSat3 repeats using the script Assembly_HSat2and3_v2.pl (https://github.com/altemose/chm13_hsat)^41^. The ape genomes include Gorilla (mGorGor1), Chimpanzee (mPanTro3), Bonobo (mPanPan1), Sumatran orangutan (mPonAbe1), Bornean orangutan (mPonPyg2) and Siamang gibbon (mSymSyn1). The more detailed annotation followed the pipeline published (https://s3-us-west-2.amazonaws.com/human-pangenomics/T2T/CHM13/assemblies/annotation/chm13v2.0_censat_v2.1.bed)^43^ for annotation of the human T2T-CHM13 assembly. For the great apes, we annotated the HSat3 subfamily with an alignment-free approach, via the pairwise distances between identified ape HSat3 and human T2T-CHM13 repeats using kmer-db (v1.11.1)^36^ with a k-mer size of 9. For HSat1 annotations, regions identified as ‘*SAR*’ by RepeatMasker were annotated as HSat1A, and regions annotated as ‘*HSATI*’ by RepeatMasker, combining the downstream or upstream *AluY* repeat with a same orientation plus (AT)n repeats were annotated as HSat1B (DYZ2) units. These combined methodologies allowed us to determine the genomic locations of HSat3 (including DYZ1), HSat1B (DYZ2), and their homologous sequences in both human and great ape genomes. To precisely annotate the boundaries of individual DYZ repeat units, a profile HMM-based search was performed using the nhmmer from the HMMER suite (v3.4)^37^. Consensus sequences for the DYZ1 and DYZ2 (https://github.com/Asian-Pan-Genome/APGp1-Y) were used as queries, with aligned hits retained only if they met a stringent E-value threshold of 1.60E-150. Only matches yielding an E-value of zero were selected as reliable DYZ repeat units for downstream analyses. Additionally, solo-*Alu* insertions occurring within the Yq12 region^31,38^ were examined across all individuals (**Figure S56**). Such insertion loci displayed different DNA methylation levels from nearby regions.

### Haplogroup phylogeny of Y chromosomes

Initial Y haplogroups of all 160 samples were assigned using phylogenetically informative SNPs from the ISOGG database (http://www.isogg.org/tree/). Paired-end reads were quality-trimmed with fastp (v0.23.4; -u 30 -q 20)^39^, uniformly downsampled to 33x coverage per sample using seqkit (v2.8.2)^40^, and aligned to the T2T-CHM13v2.0 reference genome^43^ using BWA-MEM (v0.7.18)^41^. Alignments were filtered, duplicate-marked, and processed with SAMtools (v1.9; view -bh -q 30 -F 2048 -F 256) and sambamba (v1.0.0)^42^. Variant calling was performed per sample using GATK HaplotypeCaller (v4.1.8.1)^43^ in GVCF mode followed by joint genotyping, retaining only biallelic SNPs that passed GATK Best Practices filters^44^. Variants with genotype missing rate > 0.2 or minor allele frequency (MAF) < 0.01 were excluded using VCFtools (v0.1.16)^45^. The filtered SNP set was lifted over to the hg19 reference genome using the LiftoverVcf program from Picard (v3.1.1; https://broadinstitute.github.io/picard/). A total of 8,092 SNPs located in the 10.4Mb NGS (Next-generation sequencing) accessible regions^46^ were subsequently analyzed with yhaplo (v2.1.14)^47^ for haplogroup assignment (**Table S1**). Haplogroups of three samples were manually adjusted based on the assembly alignment-based phylogeny (**Figure S57**) (see below). Haplogroup information for samples from the HPRCy1/HGSVC2 (Human Pangenome Reference Consortium Year 1/Human Genome Structural Variation Consortium Phase 2) was obtained from the original study^31^.

### Assembly alignment based phylogeny of MSY

A pangenome graph of 206 Y chromosomes (**Table S1**), including 160 samples in this study, 44 HPRCy1/HGSVC2 samples^31^, HG002-Y^5^, and CN1-Y^48^, was constructed using the Minigraph-Cactus pipeline^29^ with the HG002-Y as reference. The resulting HAL (Hierarchical alignment) format alignment was converted to MAF (Multiple alignment format) and syntenic blocks were extracted in 10Kb windows using the hal2maf tool from Cactus (v2.7.2)^50^ with the following parameters: --onlyOrthologs --noDupes --unique --maxBlockLen 10000 --noAncestors. Blocks shorter than 1Kb, and with fewer than 10 parsimony-informative sites, or with excessive missing data (> 10%) were excluded. Only syntenic blocks from the X-degenerated regions (XDRs), X-transposed regions (XTRs), and AMPL1-6 regions were retained, while the highly duplicated regions (e.g., IR3, *TSPY* array, CEN, AMPL7, Satellite regions and Yq12) were excluded due to unreliable alignment (**Figure S58**). In total, 1,657 blocks were used for phylogenetic inference (**Table S20**). A concatenated maximum likelihood (ML) tree was inferred using IQ-TREE (v2.1.4)^51^ with the following parameters: -bb 1000 -m TEST; and a coalescent-based MSY tree was reconstructed with ASTRAL-III (v5.7.1)^46^ from individual gene trees of all blocks (**Figure S59**). For both ML and coalescent trees, the genome from an A0b individual (one of the earliest-diverging Y haplogroups^52^, HG01890) was used as the root. Topological discordance between the two trees, the topology of three internal nodes (DE, C and FT) was quantified using DiscoVista (v1.0)^53^. Divergence times of internal Y-chromosomal nodes were estimated using the MCMCtree program of PAML (v4.10.7)^135^ under the GTR model (**Figure S60**).

### Evolutionary analysis of the Yq12 DYZ repeats

To obtain a better synteny of DYZ arrays between 160Ys and HG002-Y, two masked reference genomes were generated based on HG002-Y DYZ annotations^54^, in which either DYZ1 or DYZ2 regions were masked, respectively. The resulting alignment files were merged to obtain the final alignments for DYZ1 and DYZ2 arrays. To minimize the ambiguity caused by repetitive sequences and multiple alignments, only the alignment with the highest mapping quality was retained for each query segment. The whole chromosome synteny was visualized using NGenomeSyn (v1.43)^31^. We further partitioned the Yq12 region into three categories, proximal/distal terminal and inverted blocks, and the internal array, based on two peripheral inversions that were speculated to be shared across all human Y chromosomes^35^. Additional inversions were detected in several haplogroups (**Figure S61**), which were excluded from the downstream normalization analysis of DYZ block distribution (**Figure 2C**).

Phylogenetic analyses were first performed using DYZ1 and DYZ2 repeat units from HG002-Y and pericentromeric satellites (HSat3 and HSat1B) from T2T-CHM13v2.0 autosomes and great apes^55^ (**Figure 2B**). For DYZ1 phylogeny, a k-mer distance based approach was adopted. To minimize biases in k-mer frequency estimation caused by heterogeneous block sizes, HSat3 regions from Yq12 and from the acrocentric autosomes of human and apes were segmented into approximately 10Kb bins. Pairwise sequence distances between bins were then calculated using Mash distance as implemented in kmer-db (v1.11.1). Based on the resulting distance matrix, a neighbor-joining (NJ) tree was constructed with FastME (v2.0)^56^. Multiple k-mer sizes (7, 9, 11, 15, 21 and 25) were tested, and the tree inferred using 9-mer was selected for visualization. For the DYZ2 phylogeny, we focused specifically on the *HSATI* and *AluY* elements within DYZ2 copies, after excluding the AT-rich simple-repeats due to their high variability across units. The DYZ2 dataset included all 74 *HSATI*-*AluY* units located in the Yq12 terminal and inverted blocks of the HG002-Y assembly, along with a randomly chosen subset of 1,000 units from the internal array to represent the major expanded units. To investigate the evolutionary origin of DYZ2, we incorporated 510 *HSATI* and *Alu*Y sequences from the T2T-CHM13v2.0 acrocentric chromosomes (chr13, 14, 15, 21 and 22), as well as a randomly chosen subset of 371 homologous sequences from bonobo, chimpanzee, and gorilla genomes as outgroups. Sequence alignments were performed using MAFFT (v7.505) with default parameters. The ML tree was constructed using IQ-TREE (v2.1.4) with ModelFinder determined substitution models and 1,000 ultrafast bootstrap replicates (-bb 1000 -bnni -m TEST). All the trees were visualized by iTOL (v7.4)^57^.

To delineate the subgroups of DYZ1 and DYZ2 within the Yq12 region, we expanded our analysis to include sequences from all 85 gapless Y chromosome assemblies from this study, together with the HG002- and CN1-Y chromosomes (**Figure S10**). Redundant sequences (identity equal to 100%) were removed using VSEARCH (v2.29.0)^58^, and length-based filtering was applied to exclude fragmented units (DYZ1 < 1,000 bp; DYZ2 *HSATI*-*AluY* < 800 bp). High-confidence core subtype units were identified by grouping similar sequences (--cluster_fast) with identity thresholds of 0.99 for DYZ1 and 0.993 for DYZ2. For each group, the most representative sequence was selected for phylogenetic reconstruction. For the DYZ2, we used the outgroup dataset (T2T-CHM13v2.0 acrocentric HSat1B repeats and their great ape homologs) adopted in the above HG002-Y phylogeny. To verify the topological consistency, we also constructed separate trees using the *HSATI* or *AluY* sequences (**Figures S10D-F**). All ML trees were constructed using the GTR+F+G4 substitution model which exhibits the best performance in the above HG002-Y phylogeny. To complement the phylogenetic analysis and further resolve sequence heterogeneity without relying on multiple sequence alignment, we employed a k-mer based dimensionality reduction approach. For each dataset, including the DYZ1 units, the DYZ2 composite units (*HSATI*-*AluY*), and the isolated *HSATI* and *AluY* components of DYZ2, we constructed feature matrices based on the 15-mer frequencies constructed using kmer-db (v1.11.1). Principal component analysis (PCA) was performed on these matrices using the PCA module from the scikit-learn library. The first two principal components (PC1 and PC2), which capture most of the variance, were extracted to visualize the clustering landscape of the repeat units (**Figures S10B**, **C** and **G-I**).

Based on the integration of phylogenetic topology, PCA, and genomic spatial distribution, we stratified the DYZ1 and DYZ2 repeat units into three distinct lineages, designated as age groups G1, G2, and G3. The DYZ1 G1 subgroup was defined as the oldest group based on three lines of evidence: (1) it formed a clearly distinct cluster separated from other DYZ1 units in the PCA space; (2) it is located the basal position within the Yq12-specific HSat3-A6 clade in the k-mer tree (**Figures 2B** and **S10**). The DYZ1 G2 subgroup was identified as a cluster immediately adjacent to G1 in the alignment-based phylogenetic tree of all gapless samples. The remaining units, which constitute the vast majority of the internal array, were classified as the DYZ1 G3 subgroup. Similarly, the DYZ2 G1 subgroup was characterized by its distinct separation in PCA and its phylogenetic position within HSat1B clusters from autosomes (**Figure S10**). The DYZ2 G1 units were also enriched in the most distal block (**Figures 2C** and **S12A**), and the differentiation between DYZ2 G2 and G3 was further resolved based on the divergence of their internal *AluY* elements, as evidenced by specific clustering patterns in both the *AluY*-based phylogenetic tree and PCA analysis (**Figure S10**).

We defined a ‘block’ as a contiguous stretch of DYZ1 or DYZ2 repeat units that appear in an alternating manner within the Yq12 region. This organization was observed across all analyzed samples, which were all initiated with a proximal DYZ1 block and terminated with a distal DYZ2 block. We only selected the gapless assemblies from this study and excluded the samples containing fewer than 30 or more than 40 blocks for distribution normalization analyses. As a result, a final set of 27 samples were retained for the downstream later analysis. For each retained sample, we calculated the proportion of each age group (G1, G2 and G3) within each classified block. To facilitate a global comparison of subgroup distribution patterns across Y chromosomes of varying lengths, the entire Yq12 array structure was normalized to a uniform scale and partitioned into 40 bins. The proportion of each subgroup was then averaged across these windows for all samples to generate the composite distribution profile **(Figure 2C)**. As for individual blocks, we only kept those with lengths falling within the 95% confidence interval. The position of each unit was subsequently normalized (scaled 0-1) by the total length of its respective block to summarize the spatial distribution of subgroups **(Figure 2D)**.

Sequence divergence among the DYZ1 and DYZ2 units was quantified by analyzing individual repeat unit sequences extracted from each Y chromosome assembly using BEDtools (v2.31.0). For each dataset, the extracted units were aligned against the consensus sequence to prepare input files. A custom script was employed to align sequences and calculate mismatch-based divergence metrics between each unit and the consensus (https://github.com/Asian-Pan-Genome/APGp1-Y/) generated using EMBOSS CONS^38,59–61^.

### Expression analysis of Y-linked genes and repeats

Published Iso-seq data of humans^62^ were mapped to the respective reference genomes using Minimap2 (v2.26) with following parameters: -ax splice -uf --splice-flank=no --secondary=no - C5, and the resulting alignment file was sorted using SAMtools (v1.9). To annotate the Yq12 lncRNA, we selected the alignment hits and collapsed them into isoforms using collapse_isoforms_by_sam.py script from Cupcake (v28.0.0; https://github.com/Magdoll/cDNA_Cupcake) with following parameters: --dun-merge-5-shorter. The quality of the isoform annotation was assessed using SQANTI3 (v3-5.1)^63^ and HG002-Y annotation file. Low-quality isoforms were filtered out based on the criteria of ‘perc_A_downstreamTTS’ greater than 59 and ‘RTS_stage’ being TRUE. A total of 90 high-quality isoforms that were labeled as ‘non-coding’ by SQANTI3 and had a length greater than 200 bp were considered to be candidate lncRNAs. The remaining high-quality isoforms labelled as ‘coding’ were aligned against the non-redundant protein sequences (nr) from NCBI, and the isoforms that did not align to any nr sequences were also considered to be candidate non-coding RNAs (**Table S8**). The visualization of confident lncRNAs was performed using IGV (v2.12.3)^64–66^.

To minimize interference from other small RNA species in the human genome, published piRNA reads^67^ were first aligned to a curated set of small RNA sequences from T2T-CHM13v2.0 gene annotations, including snoRNA, snRNA, Y_RNA, telomerase RNA, vault RNA, RNase_MRP_RNA, RNase_P_RNA, scRNA, and tRNA. The alignment was performed using Bowtie (v1.3.1)^68^ with the parameters: -a --best --strata -v 1. Reads that did not align to any of these small RNAs were retained and subsequently aligned to the T2T-CHM13v2.0 human genome using the same parameters. We extracted the alignments mapped to HG002-Y and calculated CPM (Counts Per Million) expression using bamCoverage from DeepTools (v3.5.3)^69^. The DYZ2-encoded piRNAs were further validated by sequence searches against the human piRNA database (piRBase; hsa v3.0)^70–73^ using BLASTN (v2.15.0+) with an E-value cutoff of 1e-4. The base-pair level piRNA coverage across the HG002-Y was calculated from the aligned BAM (Binary Alignment Map) files. Individual DYZ2 repeat units extracted from the genome were aligned to the DYZ2 consensus sequence to establish a nucleotide-level correspondence. Using these alignment coordinates, the genomic piRNA counts for each unit were projected onto their corresponding positions in the consensus sequence. This process was performed independently for each DYZ2 subgroup (G1, G2 and G3) to generate lineage-specific expression profiles. We calculated the mean piRNA abundance within 100 bp windows moving in 10bp steps across the consensus length (**Figure 2H**).

The published bulk RNA-seq data from the testes of humans^74^ were mapped to the human T2T-CHM13v2.0 using STAR (v2.7.10b)^75^ using following parameters: -- winAnchorMultimapNma× 100 --outFilterMultimapNma× 100 --outFilterScoreMinOverLread 0.3 - -outFilterMatchNminOverLread 0.3, and the resulting alignment file was sorted using SAMtools. The StringTie software (v2.2.1)^76^ was used for estimating the expression levels. We acquired the published single-cell RNA sequencing (scRNA-seq) datasets derived from multiple human testis^77^. Read alignment to the complete T2T-CHM13v2.0 reference genome was performed using CellRanger (v9.0.1)^78^. To quantify the expression of repetitive elements, we employed the scTE (v1.0)^79^ using the aligned BAM files generated by CellRanger. And to capture the transcriptional dynamics of *TSPY* gene families, we distinctly annotated *TSPY/FAM197Y* array genes as each composite repeat and measured their expressions using scTE (v1.0). The single-cell gene expression matrices were processed using Scanpy (v1.11.0)^80^. Potential doublets were identified and removed using Scrublet^81^. We applied quality control filters to retain genes expressed in at least one cell and cells with a minimum of 800 detected genes, resulting in a final high-quality dataset of 34,741 cells. To mitigate the batch effects arising from different donor ages, dataset integration was performed using Harmony^76^ with default parameters. For dimensionality reduction and visualization, we constructed a nearest-neighbor graph (n_neighbors = 10, n_pcs = 50) based on the Harmony-corrected PCA space (use_rep = ‘X_pca_harmony’) and generated a Uniform Manifold Approximation and Projection (UMAP) embedding (min_dist = 0.3, spread = 0.8). Cell clusters were annotated and assigned to specific germ cell or somatic cell types based on canonical marker genes established in the related published human testis atlas^31^. The count matrix generated by scTE (v1.0) was mapped back to the individual cells. The expression of repeat elements was normalized against the total gene counts of each cell (**Figure S14**). To estimate the DYZ1-derived lncRNA expression level, we calculated the cumulative expression for each lineage by summing the raw counts of all repeat units belonging to G1, G2 and G3, respectively. These aggregated counts were subsequently log_2_(count + 1) transformed to represent DYZ1 lncRNA expression levels. For comparative analysis, all lncRNAs located within the euchromatic regions of the MSY regions were processed and normalized using the identical strategy (**Figure 2G**).

### Analyses of the *TSPY/FAM197Y* gene family

*TSPY* genes were annotated using Liftoff (v1.6.3) based on the HG002-Y gene annotation^34^. Premature stop codons were examined in each assembly to identify pseudogenes within the *TSPY* gene array. To delineate the boundaries of *TSPY/FAM197Y* units (hereafter named ‘*TSPY* units’ for simplicity) on HG002-Y, we first aligned the flanking sequences (± 10Kb) of the standalone *TSPY2* unit to the *TSPY* unit array using Minimap2 (v2.26; -cx asm10). The coordinates of the best continuous alignments were defined as the boundaries of the *TSPY2* unit. The resulting ∼20.4Kb *TSPY2* unit was subsequently aligned to each Y chromosome assembly, including those of great apes^34^, using Minimap2 (-cx asm20) to identify all *TSPY* units. For great apes, *TSPY* gene coordinates were further refined based on their published gene annotations^82^. CTCF (CCCTC-binding factor) binding sites across the full *TSPY* array in HG002-Y were predicted using the FIMO program from the MEME Suite^83^.

We first reconstructed the ML tree of *TSPY* genes using their full-length DNA sequences with IQ-TREE (v2.1.4) with the parameters: -bb 1000 -bnni -m TEST. The protein-coding *TSPY* genes in each ape species were clearly distinguishable from pseudogenes, except for those embedded within the *TSPY* array. We further incorporated flanking sequences (e.g., intergenic sequence and/or *FAM197Y* genes) of *TSPY* genes into the phylogenetic analysis (**Figure S19A**). All the trees were visualized by iTOL (v7.4). To validate the classification of *TSPY* subgroups (**Table S9**), we measured the pairwise Jaccard similarity among all subtypes using kmer-db (v1.11.1). To assess the divergence level between the *TSPY* subgroups (*TSPY2* and *TSPY_A/B/C*), all units were mapped to the *TSPY2* unit from HG01890, one of the earliest-diverging Y haplogroups (A0b), using MUMmer (v4.0.0rc1)^84^ with the parameters: nucmer – mum && delta-filter -i 95 -o 95 && show-snps -r -T. The SNP outputs from all alignments were merged and converted to VCF (Variant Call Format) format using customized scripts. Singleton and non-biallelic variants were removed prior to analysis. PCA analysis was performed using VCFtools (v0.1.16) and PLINK (v1.90)^85^. Multiple sequence alignments between the *TSPY* subgroups were generated using MAFFT (v7.505) with default parameters and visualized using AliView (v1.28)^86^. For the selection constraints, we calculated the *dN/dS* ratio for all subtypes of *TSPY* genes (**Figure S20**) under the free-ratio model implemented in the PAML package (v4.10.7), using the gorilla *TSPY* genes as outgroups. Functional motif enrichments were identified with FIMO from the MEME Suite.

The predicted protein structures of *TSPY* subgroups were obtained using AlphaFold3^87^. To assess the potential impacts of the O2a specific mutation (p.E85G) on protein-protein interactions, we employed DeepTrio (v1.0.0)^88,89^, a structure-informed deep learning framework. Interaction modeling focused on *TSPY2* (reference and mutant types) and three proteins associated with male germline biology or prostate cancer: *AR* (Androgen receptor)^90^, *ERG* (ETS-related gene)^91,92^, and *FOXA1* (Forkhead box protein A1)^93^. Protein-protein interactions were predicted using the visual_DeepTrio.py script with default settings and the pre-trained human model. For each protein pair, residue-level interaction scores were generated and normalized. Differences between reference-type and mutant TSPY2 proteins were estimated to identify residues with altered interaction potentials. To assess whether the O2a haplogroup-specific exonic variant in *TSPY2* (HG002-Y:10,226,949, T>C) affects transcription factor binding, we compared the first exon sequences of HG002-Y (T allele) and CN1-Y (C allele), which are identical except at this single site. Motif analysis was conducted using FIMO with the HOCOMOCOv11 human motif database^92^, applying stringent filters (*P-value* < 0.0001 & *Q-value* < 0.01). We focused on motifs overlapping with a 25bp interval centered on the variant site (**Figure S25**). Motifs uniquely present in either allele, or exhibiting differential binding scores, were identified and compared to evaluate potential regulatory effects of the variant.

To estimate the copy number of *TSPY* genes in the prostate cancer cohort^94^, we first assessed the accuracy of short-read–based copy number estimation using the gapless Y chromosome assemblies. Paired-end short reads from the samples in this study were aligned to the T2T-CHM13v2.0 reference genome using BWA-MEM (v0.7.18) with default parameters. Alignments were sorted and indexed with SAMtools (v1.9), followed by duplicate masking using Picard (v3.1.1). Copy number estimation was based on the normalized read depth. For the *TSPY* array, the average sequencing depth across the array was normalized to the depth of X-degenerate regions (XDRs), which permits unique mapping of short reads, and subsequently scaled relative to the known *TSPY* copy number in the HG002-Y reference assembly. The concordance between the copy numbers estimated from short-read data and those derived from gapless assemblies was evaluated and visualized (**Figure S44**). The same pipeline was applied to estimate *FAM197Y* copy numbers in the prostate cancer cohort.

### Analysis of Y-linked structural variations

To identify the large inversions (> 10Kb) in the euchromatic regions of MSY, we first compared each assembly to the HG002-Y reference using Mummer (v4.0.0rc1) with the following parameters: nucmer --maxmatch -c 500 -b 500 -l 1000 & delta-filter -m -i 95 -l 1000. The software Syri (v1.7.0)^95^ was then used to detect the rearrangements, including inversion, duplication and translocation, for the 206 Y assemblies relative to HG002-Y. As the Syri software requires chromosome-level assemblies, we used the RagTag (v2.1.0) to produce a pseudo-chromosome for the HPRCy1/HGSVC2 samples with fragmented Y assemblies, as was done for some in this study. The rearrangements spanning the junctions of the scaffold junctions were excluded. We further used PAV (v2.3.4)^96^ to identify inversions for cross-validation. The presence of large inversions (≥ 50Kb; **Table S10**) was further confirmed by visual inspection of syntenic plots of each assembly with HG002-Y reference, generated by plotsr (v1.1.1)^173^. For the two large inversions spanning AMPL1 and AMPL2 (**Figure S40**), we refined their breakpoints combining the inversion coordinates detected by both Syri and PAV, and the homologous gene position based on gene annotation liftover from HG002-Y.

The variant call set was generated from the Minigraph-Cactus graph pangenome workflow^17,29^ using 206 whole Y chromosomes with HG002-Y as reference. Following variant decomposition from the MC graph, an allele-collapsing strategy^97^ was applied to reduce redundancy in the SV representation. Briefly, for each site, all alternative alleles were clustered using VSEARCH (v2.29.0) with the following parameters: --id 0.9 --strand both --maxseqlength 100000 --minseqlength 2. Within each primary cluster, sub-clusters were further iteratively defined when an allele differed in length by ≥ 50bp from the longest allele. For downstream analyses, all alleles within a cluster that included the reference allele were annotated as reference alleles. This strategy yielded non-redundant SV allele clusters for each variant site. Across the euchromatic regions of the Y chromosome, we identified a total of 84,626 SNPs, 28,141 INDELs (insertions and deletions; <50 bp), and 1,015 SVs (insertions and deletions; ≥50 bp; **Figure S46A**). No more than ten variants were near (+/− 1Kb) assembly gaps, indicating assembly inconsistency having very limited effects on this variant set. Compared with the previously reported pairwise PAV-based variant catalog^98^, our graph-derived call set recovered 60.1% of SNPs, 45.5% of INDELs, and 50.8% of SVs while identifying numerous additional variants, due to differences in both sample composition and variant discovery strategy. Relative to HG002-Y, each individual carried on average 50 and 47 SVs, and 2,078 and 7,734 small variants (SNPs and INDELs) within the PAR and MSY, respectively. The PAR harbored a substantially higher density of common variants and nucleotide diversity (median value of 10Kb windows: 4.65e-04), whereas the non-recombining MSY was enriched for rare variants and singletons (**Figure S46B**) and exhibited low nucleotide diversity (median value of 10Kb windows: 8.31e-05), consistent with long-term recombination suppression. Within the MSY, variant density varied markedly among sequence classes, with ampliconic regions showing the highest burden of SVs, whereas X-transported regions exhibiting the lowest variant density (**Figures S46C-D**). Despite the availability of more than 200 high-quality Y chromosome assemblies, the accumulation curve of detected variants has not yet reached saturation, suggesting that additional Y-chromosome diversity remains to be discovered. The Hudson Fixation Index (*HFst*)^99^ was measured to quantify SVs stratification between populations or haplogroups. For small variants (SNPs and INDELs), multiallelic sites were first normalized into biallelic records using ‘bcftools norm’^100^, followed by *Fst* calculation with customized scripts. The deletion in the DDX3Y promoter region was genotyped for the prostate cancer cohort using PanGenie v4.2.1^34^. The nucleotide diversity was calculated using bcftools and vcftools, based on biallelic SNPs. To predict the regulatory effects of target SVs, we scanned each allele sequence of the transcription factor (TF) binding motifs using the FIMO program from the MEME Suite. The gene expression data of lymphocyte cell lines from the Multi-Ancestry Analysis of Gene Expression (MAGE)^101^ were integrated for samples with desired mutations, allowing SV alleles to be linked to transcript abundance levels.

To classify the amplicons of AMPL7, all amplicon sequences of the HG002-Y reference were aligned to ape Y chromosome assemblies^102^ using Minimap2 (v2.26; -cx asm20) to identify homologous regions. For each amplicon, the highest-scored alignment was retained to define its coordinates, and overlapping alignment intervals belonging to the same amplicon family were merged. For red, blue, and green amplicons, which have homologous sequences in the bonobo Y chromosome, the corresponding bonobo sequences were used as the outgroup to root the phylogenetic trees. For amplicon families (teal and yellow) lacking intact homologous sequences (shared homologous sequences are larger than 80%) in non-human primates, trees were rooted with the genome of an individual (HG01890) from the A0b branch. Amplicons with incomplete structure (< 80% of the average length within each family), likely resulting from assembly artifacts, were excluded from downstream analyses. Multiple sequence alignments of each amplicon family were generated using MAFFT (v7.505) and subsequently trimmed with ClipKit (v2.3.0) under the gappy mode^103^. ML trees were then constructed using IQ-TREE (v2.1.4) with following parameters: -bb 1000 -bnni -m TEST. Tree visualization and annotation were performed using iTOL (v7.2). Based on the resulting clustering patterns, amplicons were classified into subgroups. Four families, including red, blue, green and gray, exhibited well-supported sub-clusterings (**Figure S30**), whereas teal and yellow families showed no clear phylogenetic sub-clusterings (**Figure S29**). To validate the accuracy of subgroup classification, we compared the phylogeny-based assignments with gene-based annotations transferred from HG002-Y using Liftoff (v1.6.3). Because the *DAZ* (Deleted in Azoospermia) genes generally exhibit a one-to-one correspondence with each red amplicon subtype (e.g., *DAZ1/2* within *r1/r2* and *DAZ4/3* within *r3/r4* in HG002-Y; **Figure S62**), *DAZ* paralog annotations served as an internal consistency control. Across all red amplicons investigated, 93% showed consistent correlation between subgroup and gene annotation. Of the 46 inconsistencies, 26 (57%) exhibited amplicon gene conversions and involved apparent *DAZ* mis-annotations by Liftoff. For the remaining discrepancies, annotation accuracy was further assessed by comparing sequence identity to the corresponding HG002-Y *DAZ* paralogs as defined by each method. In cases where Liftoff and phylogeny-based annotations disagreed with each other (e.g., *DAZ2* versus *DAZ3*), the annotation with higher sequence similarity to the HG002-Y reference was considered more reliable.

### Ancestral haplotype inference and rearrangement analysis

Using the phylogeny-based amplicon subgroup annotation achieved above, we inferred the copy number and orientations of amplicons within AMPL7 structures of 175 haplotypes which show no obvious structural disruption of amplicons, abnormal HiFi read depth or assembly inconsistencies according to our manual examinations (**Table S11**). In total, 76 unique haplotypes were identified, including 30 non-singleton haplotypes (**Table S13**). Haplotypes were sequentially numbered based on sample composition and consistently referenced in all subsequent analyses; for example, Hap1 denotes the most frequent haplotype, comprising 16 individuals. To infer evolutionary relationships among haplotypes, ancestral haplotype states at internal nodes of major Y haplogroups (from CT to NO) were reconstructed using ANGES (v1.01)^104^, with amplicon subtype order and the concatenated phylogenetic tree as inputs. Because divergence among AMPL7 amplicons postdated the split of CT haplogroups (**Figure S33A**), the inferred ancestral haplotype at the CT node (Hap6; *n* = 6; **Tables S12-13**) was selected as the reference for rearrangement analyses.

Pairwise rearrangement distances and structural changes, including inversions, translocations, and deletions, were inferred using the DCJ-indel (double-cut-and-join with insertions and deletions) model implemented in UniMoG^103^. As current DCJ-based frameworks do not explicitly model duplications or gene conversion events, each haplotype was first reverted to a pre-duplication and pre-conversion state prior to DCJ analysis. After distance estimation, duplication and conversion events were reintegrated into the distance matrix to capture the full spectrum of structural rearrangements. This two-step strategy enabled incorporation of complex rearrangements that are otherwise not supported by standard DCJ models. Based on the revised DCJ distance matrix, a biologically informed neighbor-joining (BioNJ) tree was constructed using FastME (v2.0). This tree served as a reference for manual refinement of rearrangement events across haplotypes. Finally, all inferred rearrangements of each haplotype, including inversions, conversions, deletions and duplications, were compiled for each haplotype relative to the ancestral CT haplotype (Hap6; **Table S12**). Inversions were further categorized into intra-palindromic and inter-palindromic types, with the latter spanning two distinct palindromes. Haplotype networks for all 76 unique haplotypes, as well as for the subset of 30 non-singleton haplotypes, were constructed using PopART (v1.7)^104^ to visualize structural diversity and inferred evolutionary trajectories within AMPL7 (**Figure S35**; **Table S13**).

### SNP/Indel marker-based analysis for validating AMPL7 structural rearrangements

The traditional technologies used for SV validation, e.g., Hi-C and bionano sequencing, were less effective for the AMPL7 regions due to ambiguous alignments (**Figure S63**). To provide a validation independent of sequence alignment and sub-amplicon classification, we developed a marker-based algorithm for the highly repetitive AMPL7 region (**Figure S36**). Amplicons of the same color class from all samples were first projected onto a unified reference sequence, allowing the identification of SNP/Indel markers defined by their reference coordinates and allele states. This process was achieved by the combined pipeline of nucmer, delta-filter and show-snps, as described above. For each amplicon, these markers were ordered according to their reference coordinates to generate allele-marker chains. To prevent local mutation hotspots from disproportionately skewing the similarity metrics, we only retain one representative marker per 100-bp interval. Sliding window strategy (20Kb of window size and 10Kb of step size) was then applied along the AMPL7 coordinates, and the proportion of shared markers between homologous windows from the same amplicon families was calculated as the local similarity score. For instance, each window from a red amplicon (e.g., r1 of sampleA) would have four homologous windows (namely, r1-r4 of sampleB). Finally, the syntenic relationships were visualized using a dotplot matrix. To optimize the visual representation of marker density and minimize background noise, the dot size was scaled to the log_2 logarithm of the total marker count within each window. Consequently, syntenic collinearity and structural rearrangements, such as inversions between palindromic amplicons, were delineated by the presence of dense, highly similar marker clusters forming distinct diagonal or anti-diagonal paths, respectively. This marker-based approach provides substantially higher resolution for validating structural rearrangements inferred from sub-amplicon synteny analysis and refining breakpoint intervals within complex palindromic amplicons (**Figure S37-39**).

### Detecting amplicon micro-deletions by short-read data

For each amplicon family, all amplicons from the 175 individuals were aligned to the corresponding reference amplicons (*r1*, *b2*, *g1*, and *gy1*) from HG03248 (E1a), which exhibits a haplotype similar to the inferred CT ancestor. Variants were identified using MUMmer (v4.0.0rc1), and SNP and INDEL calls from all amplicon subgroups were merged and converted to VCF format using customized scripts. Singleton and non-biallelic variants were filtered prior to downstream analyses. The PCA was performed separately for each of the four amplicon families to validate phylogenetic sub-clustering (**Figure S31**). Red amplicons exhibited more dispersed clustering than the other families. Diagnostic sites for each amplicon subgroup were defined as positions where the frequency of the ‘alternative allele’ differed from the reference (*r1*, *b2*, *g1* and *gy1* of HG03248) by more than 0.90 within a specific subgroup (e.g., *r1* or *g3*) or a subgroup pair (e.g., *r3/r4*), relative to all other subgroups (**Figure S42**). To validate these sites using short-read data, we generated pairs of 31-mer sequences representing the reference (‘Ref’) and alternative (‘Alt’) alleles. For instance, if the *r1* amplicon carries an alternative allele at a frequency of 0.99 (relative to the HG03248 *r1* reference), while *r2*, *r3*, and *r4* amplicons maintain low alternative allele frequencies (e.g., 0.01), the Alt-derived k-mer serves as an *r1*-specific diagnostic marker. For samples with short-read alignments to T2T-CHM13v2.0, ‘Ref’ and ‘Alt’ k-mer counts at these diagnostic sites were normalized by the expected depth (**Figure 4G**), which was estimated from control haploid regions (XDR3, HG002-Y: 12,656,463 - 14,891,106), to account for variation in sequencing depth.

