## Supplementary Figures for "Origin and structural evolution of the complex genomic regions of human Y chromosome"

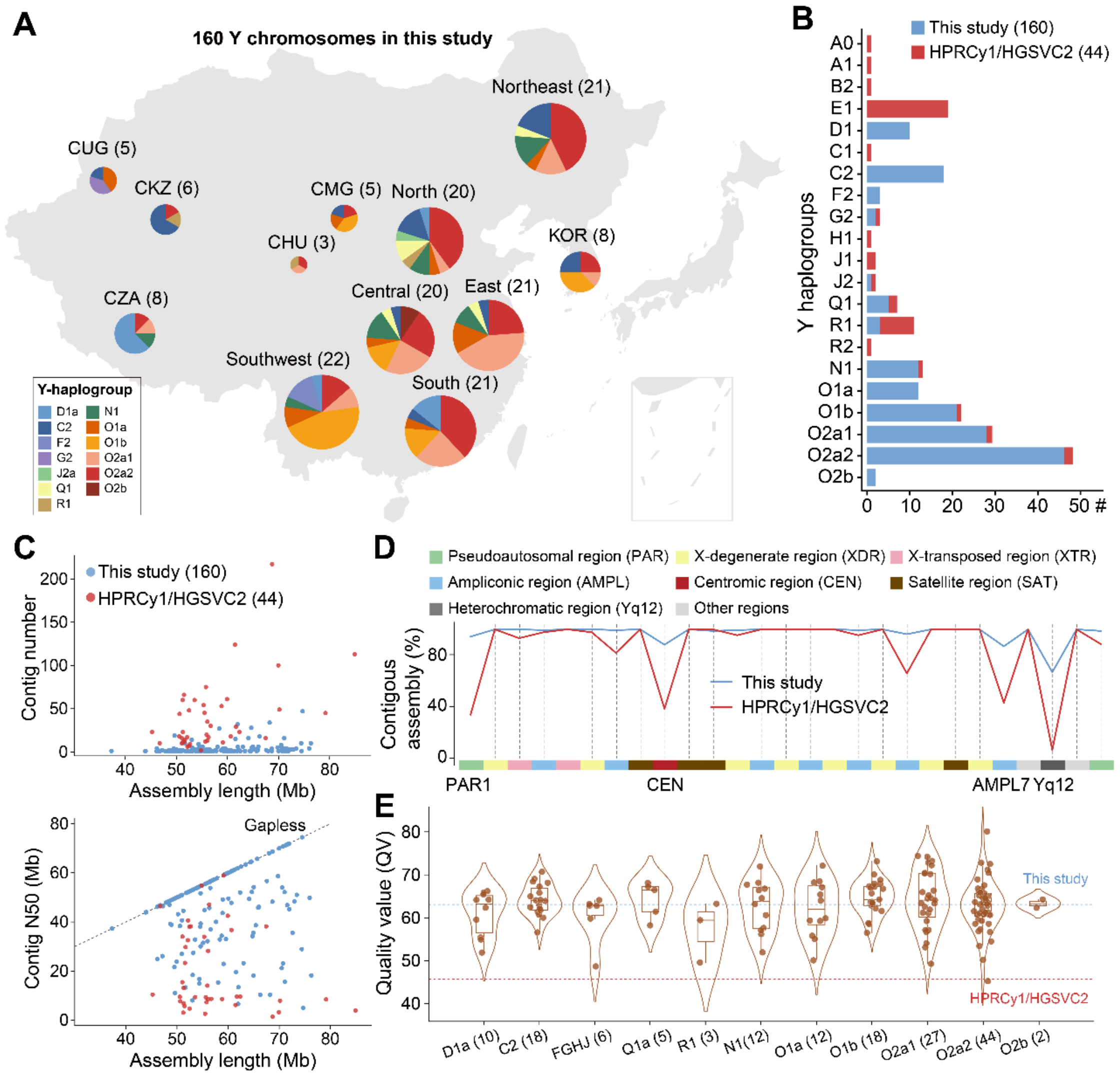


**Figure S1. Sampling and Quality assessment of 160 Y chromosome assemblies in this study.**

**A)** The geographical distribution of 160 East Asian haplogroups. CUG: Chinese Uyghurs; CKZ: Chinese Kazakhs; CZA: Chinese Tibetans; CHU: Chinese Hui; CMG: Chinese Mongolians; KOR: South Koreans. **B)** The major haplogroup comparison of 160Ys ($n$ = 160) and HPRCy1/HGSVC2 ($n$ = 44) samples. **C)** Assembly length versus the number of contigs for Y chromosomes. Blue dots represent the 160 samples in this study, while red dots represent HPRCy1/HGSVC2 samples[1]. Assembly length versus contig N50 for Y chromosomes. The dashed line marks the theoretical maximum for gapless assemblies. 160Y assemblies show longer contigs with higher continuity. **D)** Subregion-level assessment of contiguity for 160Ys (blue) and HPRCy1/HGSVC2 (red) assemblies. The y-axis represents the proportions of samples with gapless assembly in each subregion. Continuity is evaluated across Y-chromosomal subregions: Pseudoautosomal region (PAR), X-degenerate (XDR), X-transposed (XTR), Ampliconic (AMPL), Centromeric (CEN), Satellite (SAT), Heterochromatic (Yq12) and others. The assemblies in this study demonstrate consistently higher contiguity across most subregions. **E)** Violin plots of QV (Quality value) for 160 samples grouped by haplogroup. The blue dashed line indicates the mean QV of 160 samples in this study; the red dashed line shows the mean QV of HPRCy1/HGSVC2 assemblies for comparison.

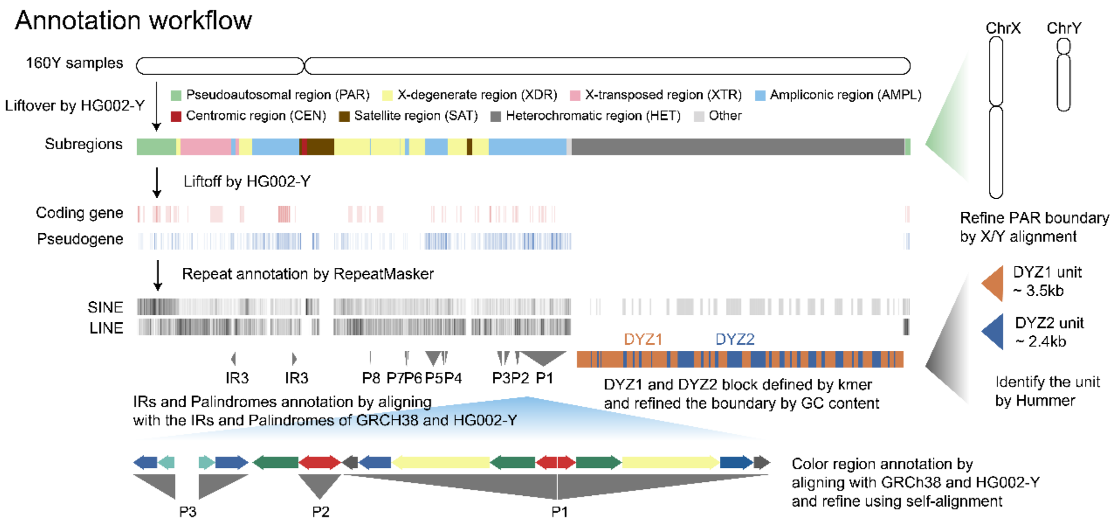


**Figure S2. Annotation workflow for 160Y chromosomes in this study.**
Subregions and genes were annotated using coordinate liftover from the HG002-Y reference[2]. Repeat elements, including SINEs and LINEs, were annotated using RepeatMasker (CONS-Dfam 3.3). DYZ1 and DYZ2 satellite blocks were defined based on kmer composition and refined by GC content, with repeat unit identification by HMMER[3]. Inverted repeats (IRs), palindromes and amplicon families were annotated through their alignments with GRCh38 and HG002-Y references, followed by self-alignments for refinement (**METHODS**). Complex repeat-rich regions such as the *TSPY* array and AMPL7 were manually curated. Additionally, the PAR boundary was refined through alignment between chrX and chrY.

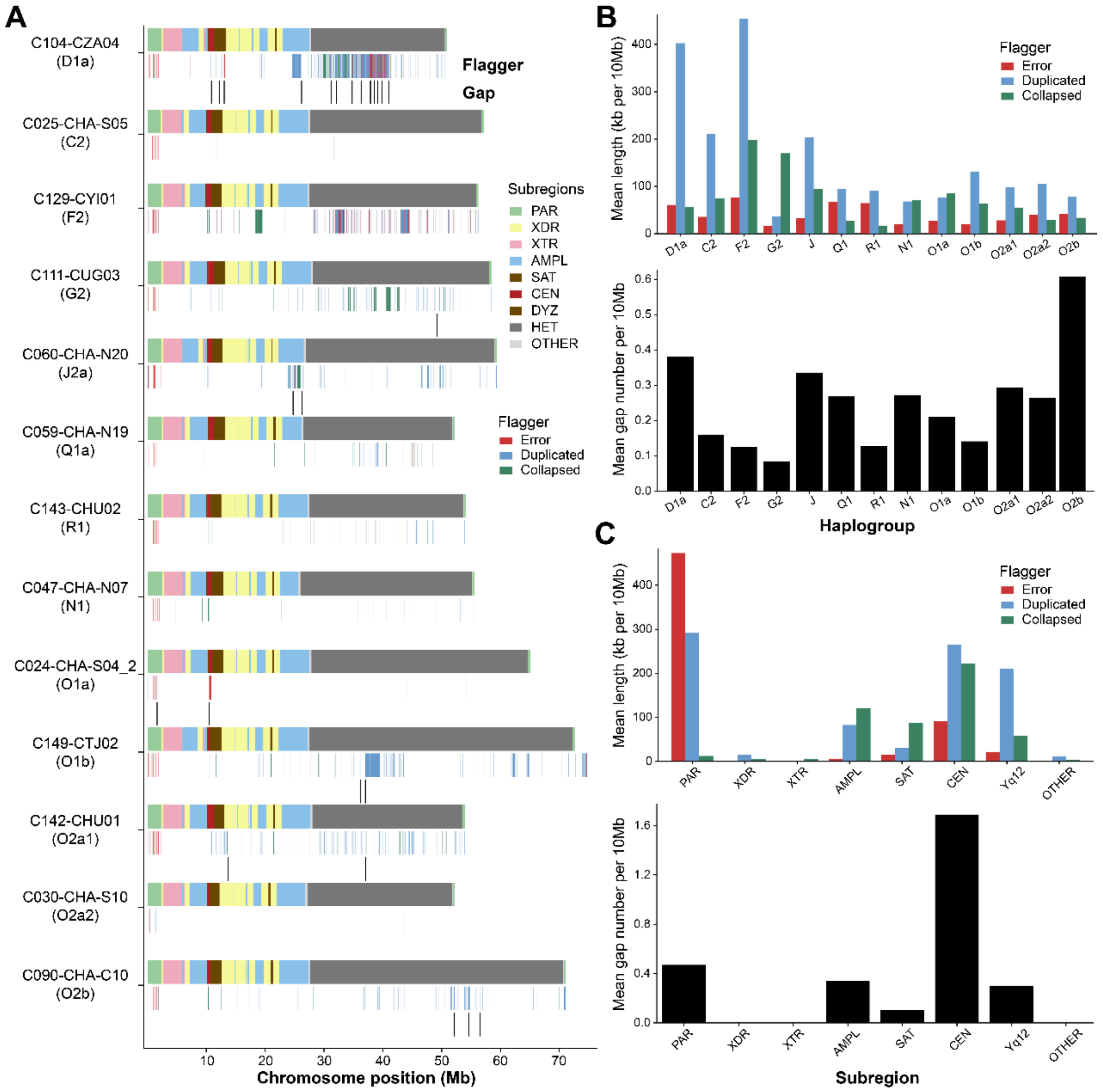


**Figure S3. The assembly quality evaluation for 160Y chromosomes.**

**A)** The flagged and gapped regions for 13 randomly selected Y chromosomes, each for one major haplogroup. The averaged normalized (per 10Mb) length of flagged regions and gap number for 13 major haplogroups **(B)** and eight major subregions **(C)**. The definition of flagged regions follows HMM-Flagger[4]. Error: the blocks with low read coverage. Duplicated: the blocks which are potentially a false duplication of another block. They should mainly include low-MAPQ alignments with half of the expected coverage. Collapsed: Two or more highly similar haplotypes collapsed into one block. The remaining represents the well-assembled haploid regions.

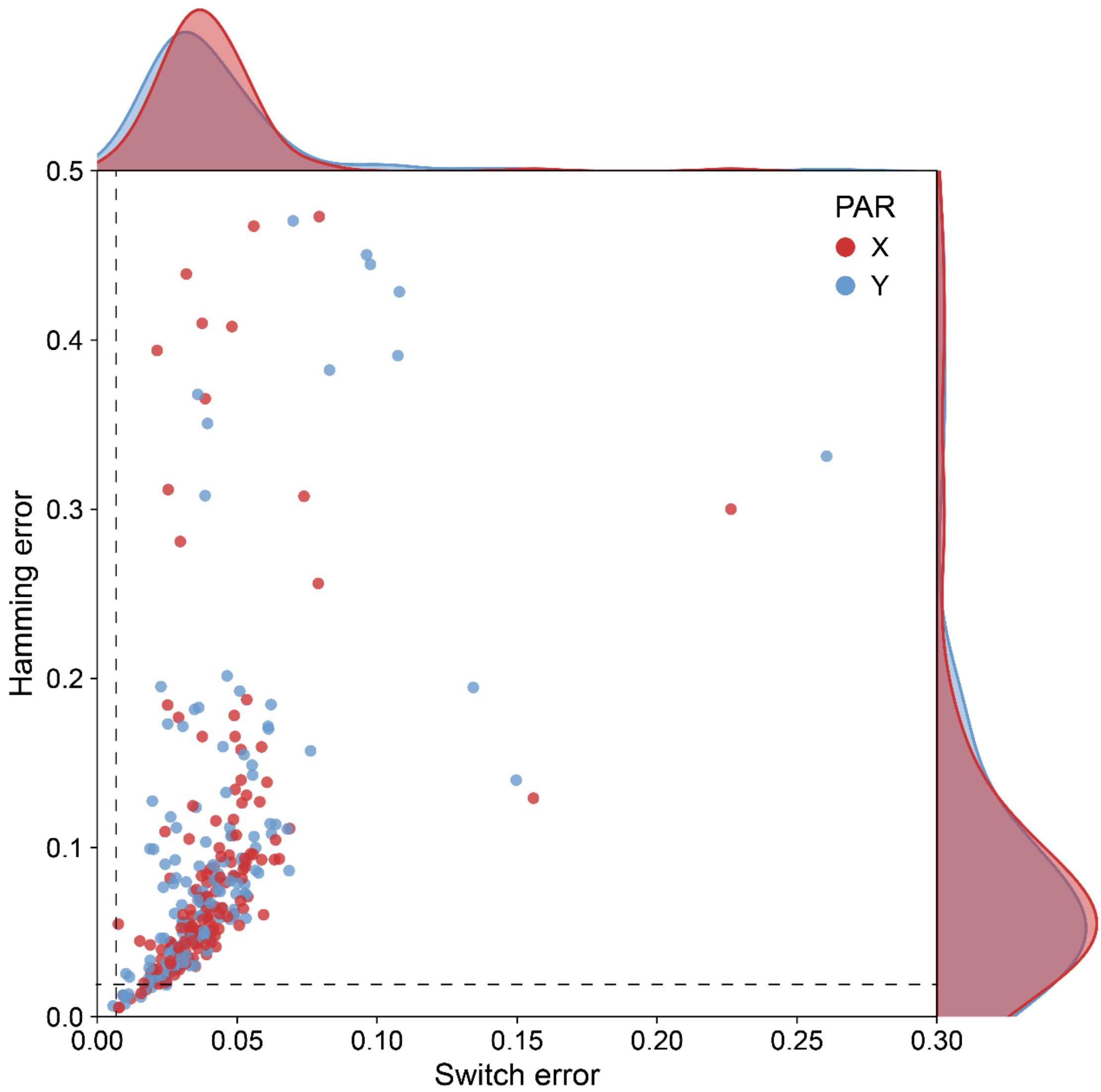


**Figure S4. Switch and hamming error for the PAR regions.**
On average, switch error rates of 4.03% and 4.05% were estimated for the PAR regions of chromosomes X and Y, respectively, compared to 0.64% (maternal haplotype) and 0.34% (paternal haplotype) for the whole genome (dotted lines)[5]. Similarly, the average hamming error rates were 9.20% and 9.27% for the PARs of chromosomes X and Y, respectively, versus 1.68% (maternal haplotype) and 0.68% (paternal haplotype) at genome wide.

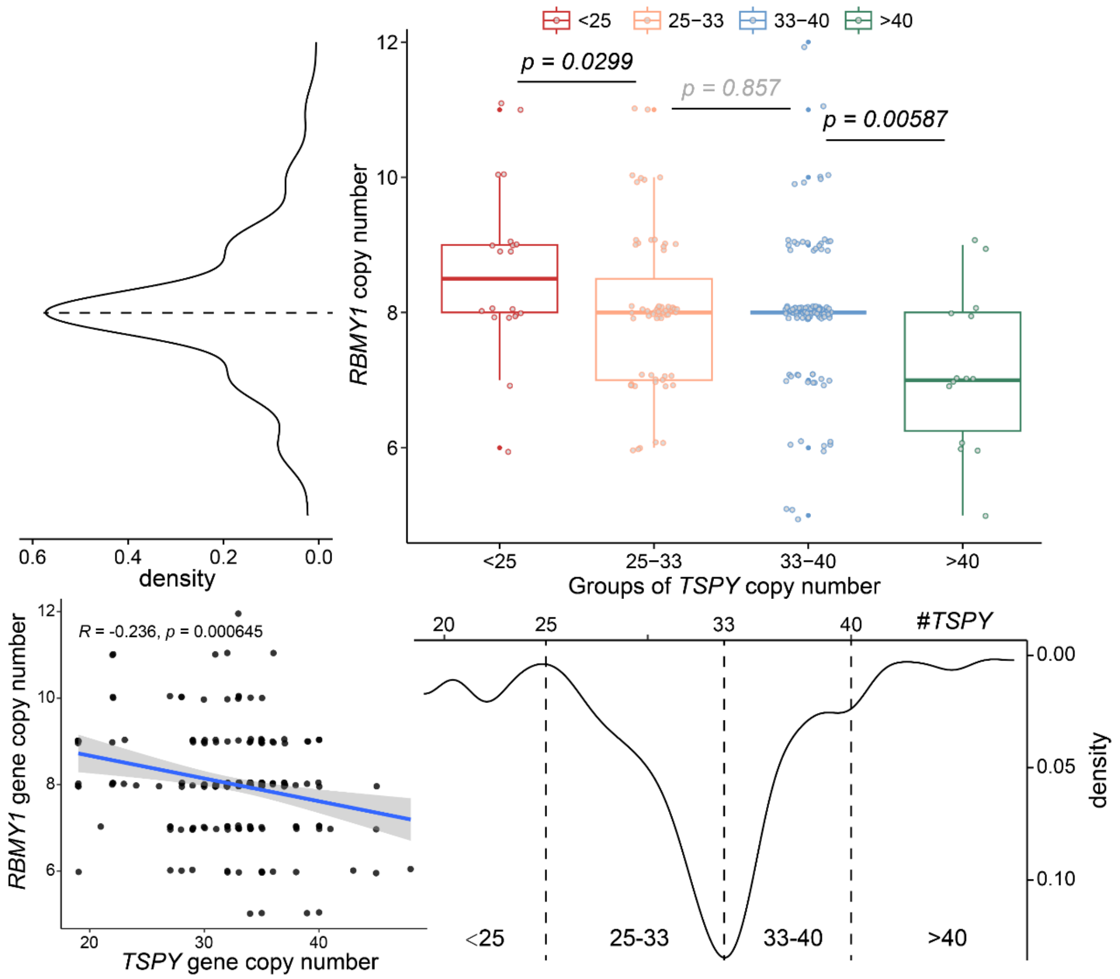


**Figure S5. A negative correlation of gene copy numbers between *TSPY* (Testis-specific protein, Y-encoded) and *RBMY1* (RNA-binding motif gene 1 on Y chromosome).**

The negative association is more obvious when comparing the *RBMY1* copy number between samples with low-copy (< 25) and high-copy *TSPY* (> 40) groups, represented by the boxplot. Pearson correlation test was performed for *TSPY* and *RBMY1* genes.

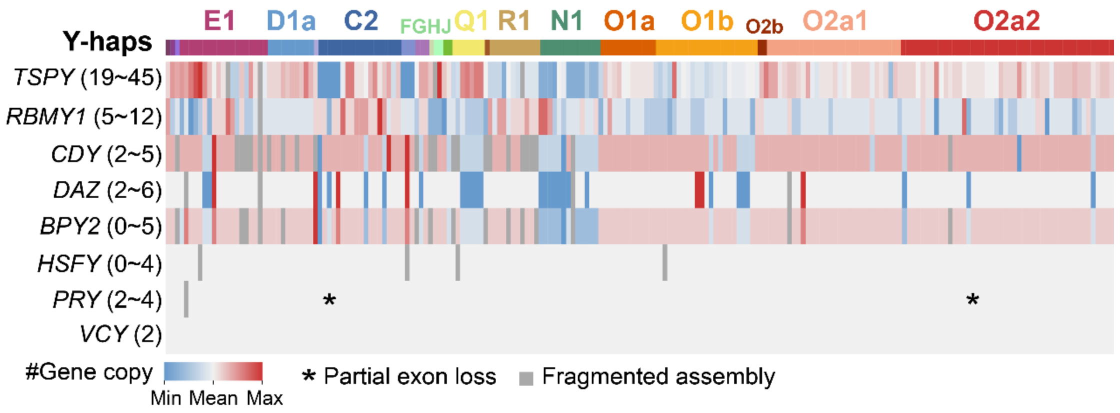


**Figure S6. The copy number variation of eight protein-coding gene families across Y haplogroups.**

The star marks represent the partial exon loss in *PRY* (PTPN13 Like Y-Linked) gene. The gray color represents ambiguous gene copy estimation due to assembly gaps. The numbers in the brackets represent the copy number range for each gene family. *CDY*: Chromodomain Y-Linked 1. *DAZ*: Deleted in Azoospermia. *BPY2*: Basic Charge Y-Linked 2. *HSFY*: Heat Shock Factor Y-Linked. *VCY*: Variable Charge Y-Linked.

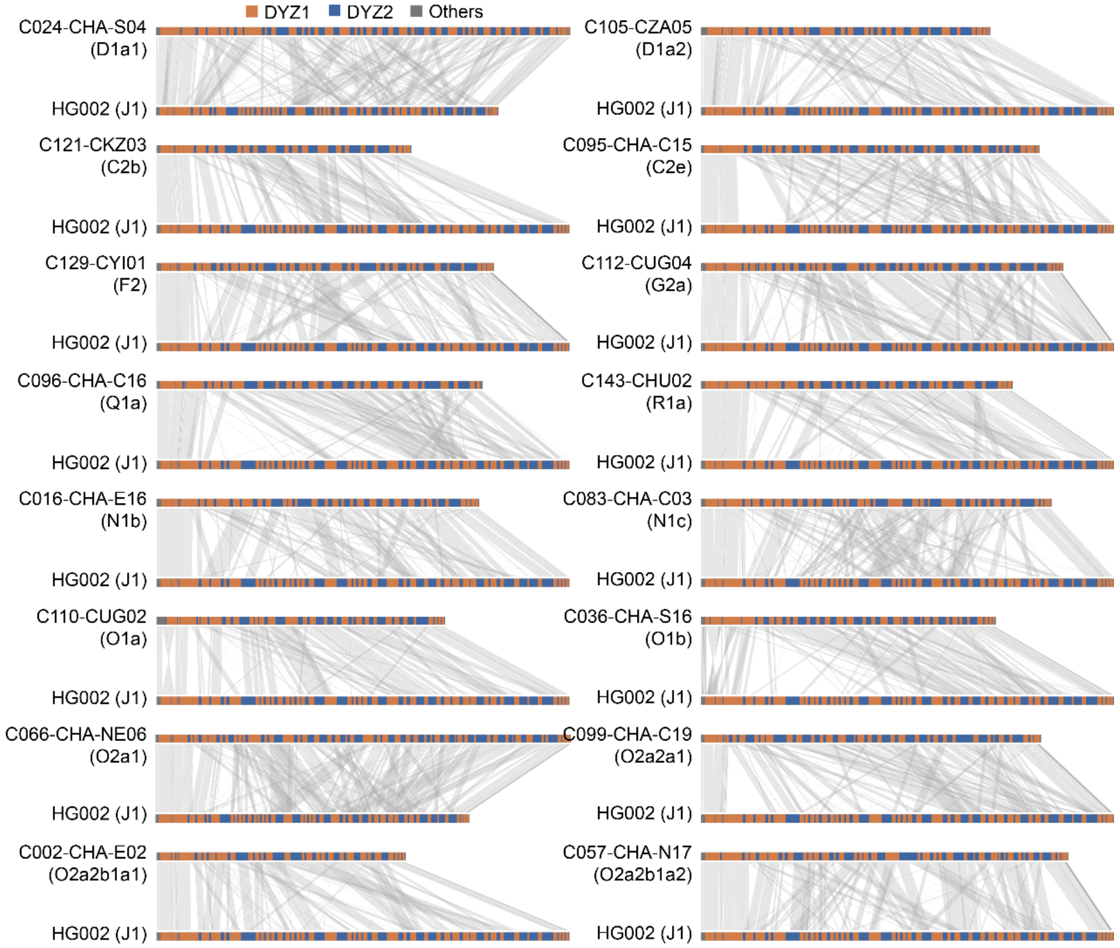


**Figure S7. The DYZ organization and synteny plots of 16 representative Y haplogroups from 160Y samples compared to HG002-Y for the Yq12 regions.** Pairwise alignments were performed using minimap2[6]. For the multiple alignments, only ones with the highest alignment quality and length were retained. 'Others' represents the DYZ18 repeats.

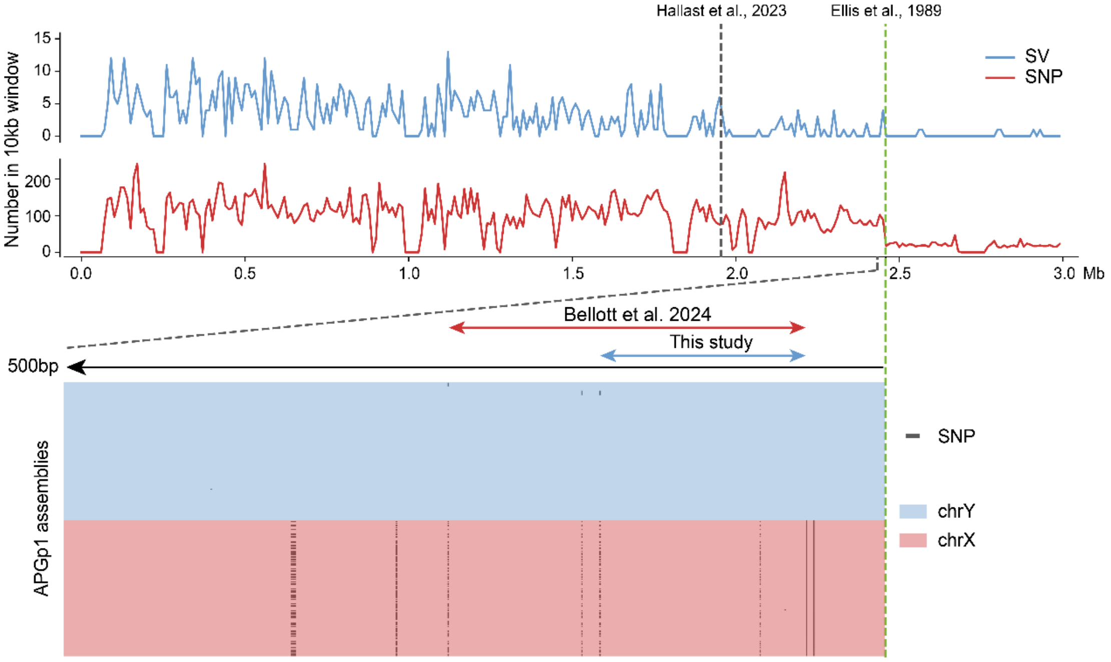


**Figure S8. The PAR1 boundary was re-defined using new variants in this study.** Previous study[1] suggests a fronted PAR1 boundary due to the lack of SV (structural variation) in the region of about 500Kb long before the canonical boundary[7]. Consistent with another study[8], the SNP (single nucleotide polymorphism) linkage between X and Y chromosomes suggests the real boundary might lay about 50bp ahead of the canonical boundary.

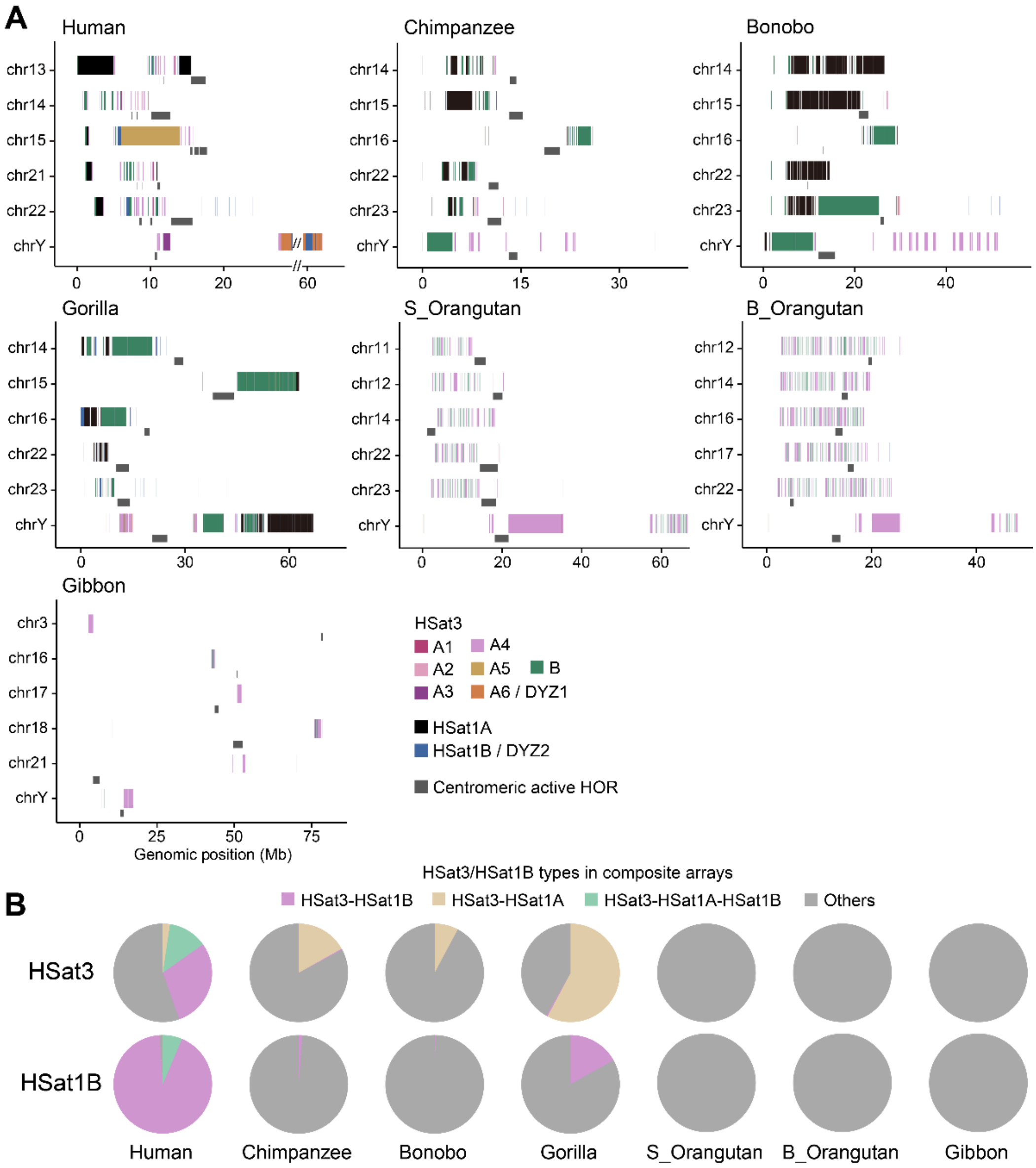


**Figure S9. Satellite sequences identified in great ape telomere-to-telomere (T2T) assemblies.**

**A)** Identification and annotation of human satellite sequences (HSat1A, HSat1B and HSat3) on recently released great ape genomes[9]. HSat3-A6 and HSat1B constitute the major components of the DYZ1 and DYZ2 repeats, respectively. HSat1A and HSat1B units were annotated using RepeatMasker (v4.1.2; Dfam 3.3), whereas HSat3 annotations were further refined using published scripts[10] (https://github.com/altemose/chm13_HSat). For visualization, the six chromosomes exhibiting the highest proportions of HSat3, HSat1A, or HSat1B repeats were selected. Gray narrow bars indicate active higher-order repeats (HORs) in centromeric regions. **B)** Proportions of different HSat repeats within composite satellite arrays across ape species. The alternating HSat3-HSat1B (DYZ1-DYZ2) array shows the highest proportion in humans, consistent with the large expansion of the Yq12 region. Only composite arrays in which all constituent units share the same orientation (e.g., HSat3(+)-HSat1B(+) or HSat3(-)-HSat1A(-)-HSat1B(-)) were included, whereas arrays composed of units with mixed orientations (e.g., HSat3(+)-HSat1B(-)) were classified as ‘Others’.

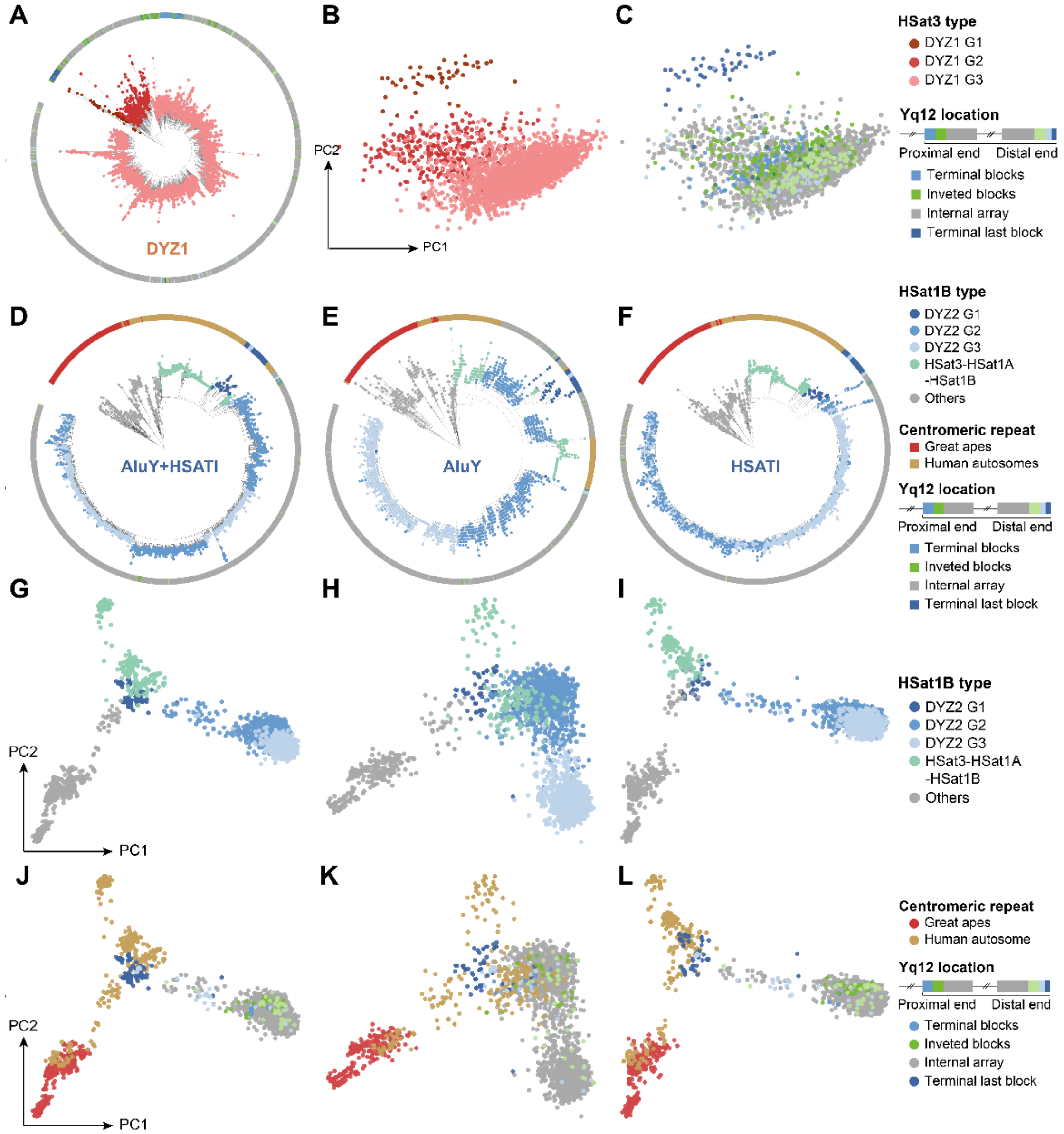


**Figure S10. Sequence classification of DYZ1 and DYZ2 satellite arrays.**
**A)** Maximum-likelihood (ML) phylogenetic tree of core DYZ1 repeat units (*n* = 3,166; alignment length > 1.0Kb) derived from 85 gapless Y chromosomes, together with HG002-Y[2] and CN1-Y[11]. Core units were defined as repeat units exhibiting pairwise sequence similarity < 99% (**METHODS**). The tree resolves three major subgroups (G1, G2 and G3), indicated by different dot colors. Circos bars denote the physical locations of units along Yq12. DYZ18 (HSat3-A4) was used as the outgroup. These results demonstrate that DYZ1 repeats have diversified into distinct lineages. **B-C)** Principal component analysis (PCA) of all core DYZ1 units based on 15-mer frequency profiles, colored by DYZ1 sub-groups (**b**) and by their genomic positions within Yq12 (**C**). **D-F)** ML phylogenetic trees of DYZ2 core units constructed from *AluY+HSATI* (**D**), *AluY* (**E**), and *HSATI* (**F**) sequences (*n* = 4,243; alignment length > 800bp). Core units were defined as those whose *AluY+HSATI* sequences exhibit pairwise similarity < 99.3%. Homologous sequences from composite satellite arrays on other human chromosomes (e.g., chr13, 14, 15, 21 and 22) as well as from ape chromosomes were included for tree construction. The internal node that distinguished gorilla clade was re-rooted for better visualization of DYZ2 structure. Dot colors indicate different HSat1B types, and circos bars represent species identity and Yq12 locations (for DYZ2 sequences only). The

structural discrepancies observed between G2 and G3 subgroups across these three trees suggest that the individual components of the DYZ2 unit (*AluY* and *HSATI*) may be subject to distinct evolutionary pressures or recombination dynamics. **G-L)** PCA of all core DYZ2 units based on 15-mer frequency profiles, annotated by DYZ2 sub-groups (**G-I**) and by ape species and Yq12 location (**J-L**).

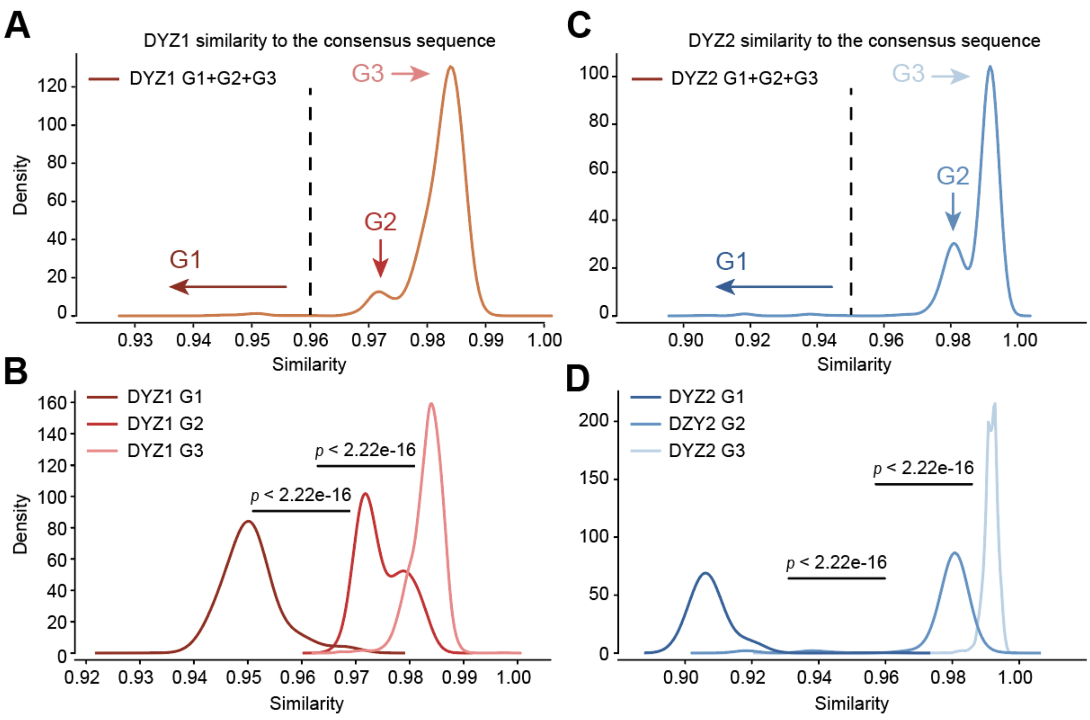


**Figure S11. The unit similarity of DYZ sub-groups with the consensus sequence, respectively.**

**A)** and **C)**, the similarity of all DYZ units compared to consensus sequence. **B)** and **D)**, the separated density plots of DYZ sub-group similarity compared to consensus sequence.

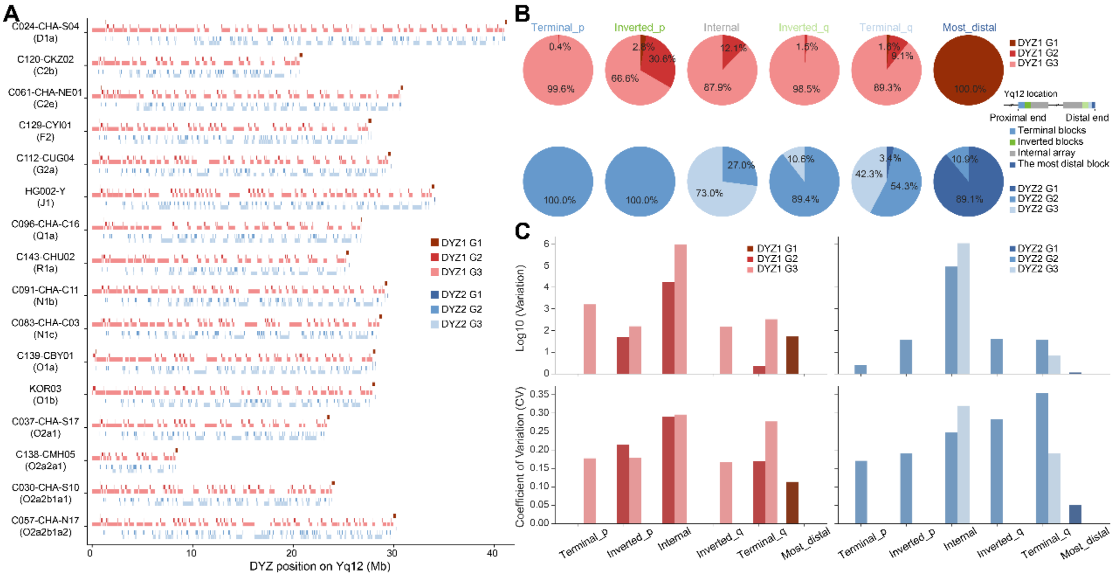


**Figure S12. The distribution, composition and copy number variations (CNVs) of DYZ subgroups within the Yq12 region.**

**A)** Representative samples with gapless Yq12 assemblies, randomly selected from each major haplogroup. Note that G1 DYZ units consistently localize to the last terminal block across all samples. **B)** Mean proportions of DYZ subgroups across Yq12 subregions. To minimize confounding effects from structural rearrangements, analysis was restricted to individuals (*n* = 45) possessing gapless assemblies and lacking non-canonical inversions (i.e., retaining only the two fixed peripheral Yq12 inversions; **METHODS**). The legend indicates the relative positioning of DYZ blocks: 'Terminal_q', 'Inverted_p', 'Internal', 'Inverted_q', and 'Most_distal'. **C)** Copy number variation (upper) and corresponding coefficients of variation (CV; lower) for DYZ repeats. For cross-regional comparison, only subgroups with a mean copy number > 5 are shown to prevent CV (coefficient of variation) inflation caused by low-abundance repeats.

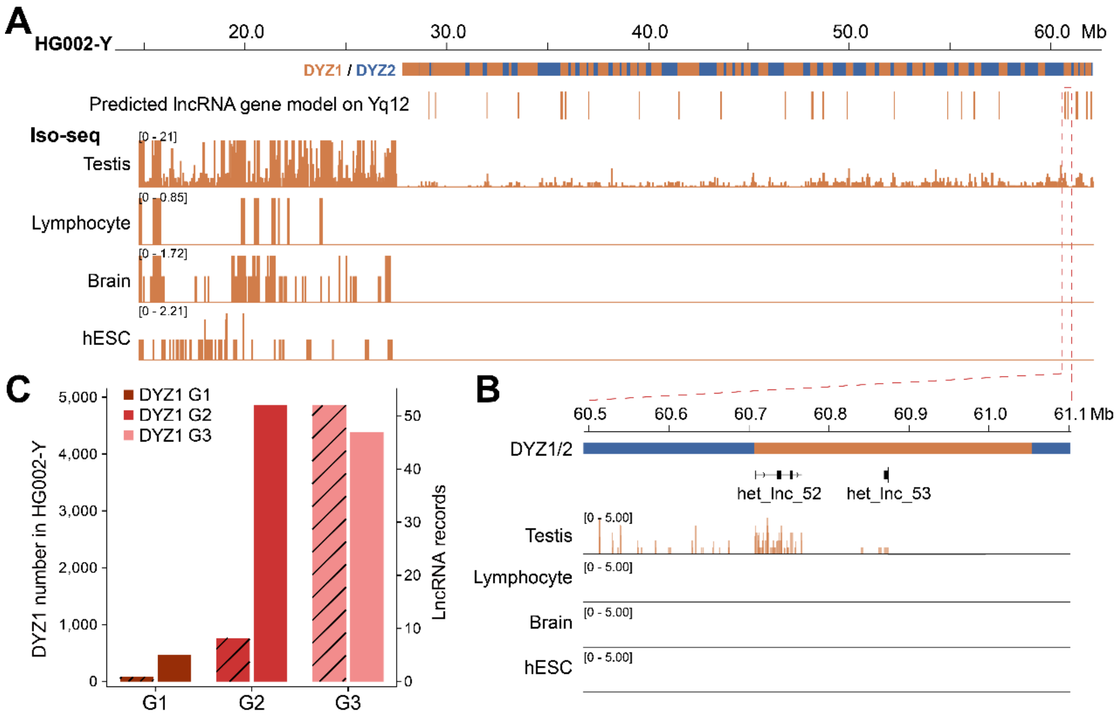


**Figure S13. The lncRNA transcripts in Yq12 DYZ1 units across different tissues.** **A)** The lncRNA signals derived from DYZ1 repeats in testis but no other tissues. The transcripts from testis Iso-seq data were predicted for lncRNA gene models ($n$ = 90, confident models) (**METHODS**). hESC: human embryonic stem cell. **B)** Examples for the predicted lncRNA genes. **C)** The predicted lncRNA records among DYZ1 sub-groups. Although DYZ1 G1/2 have fewer copies, they overlap with the higher number of predicted lncRNA models.

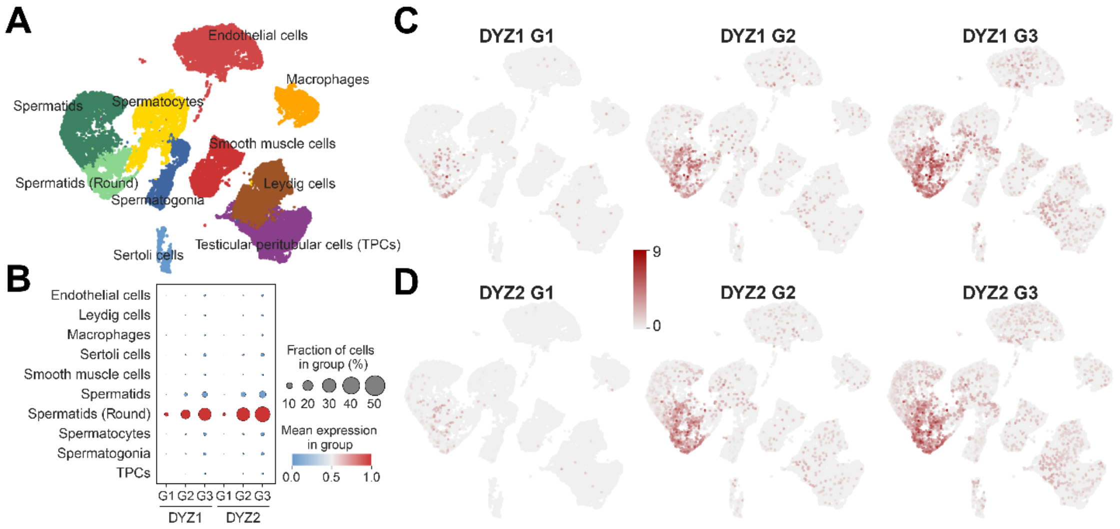


**Figure S14. The expression of DYZ subgroups in testis scRNA data.**

**A)** Cell-type annotation of the testis samples (*n* = 6). Cell-type markers were adopted from a previous study[12]. **B-D)** Both DYZ1 and DYZ2 show significant expression in round spermatids. Expression levels of DYZ subgroups were aggregated and treated as a single locus using the sc-TE pipeline[13]. Thus, the expression is tightly related to the copy number of each subgroup.

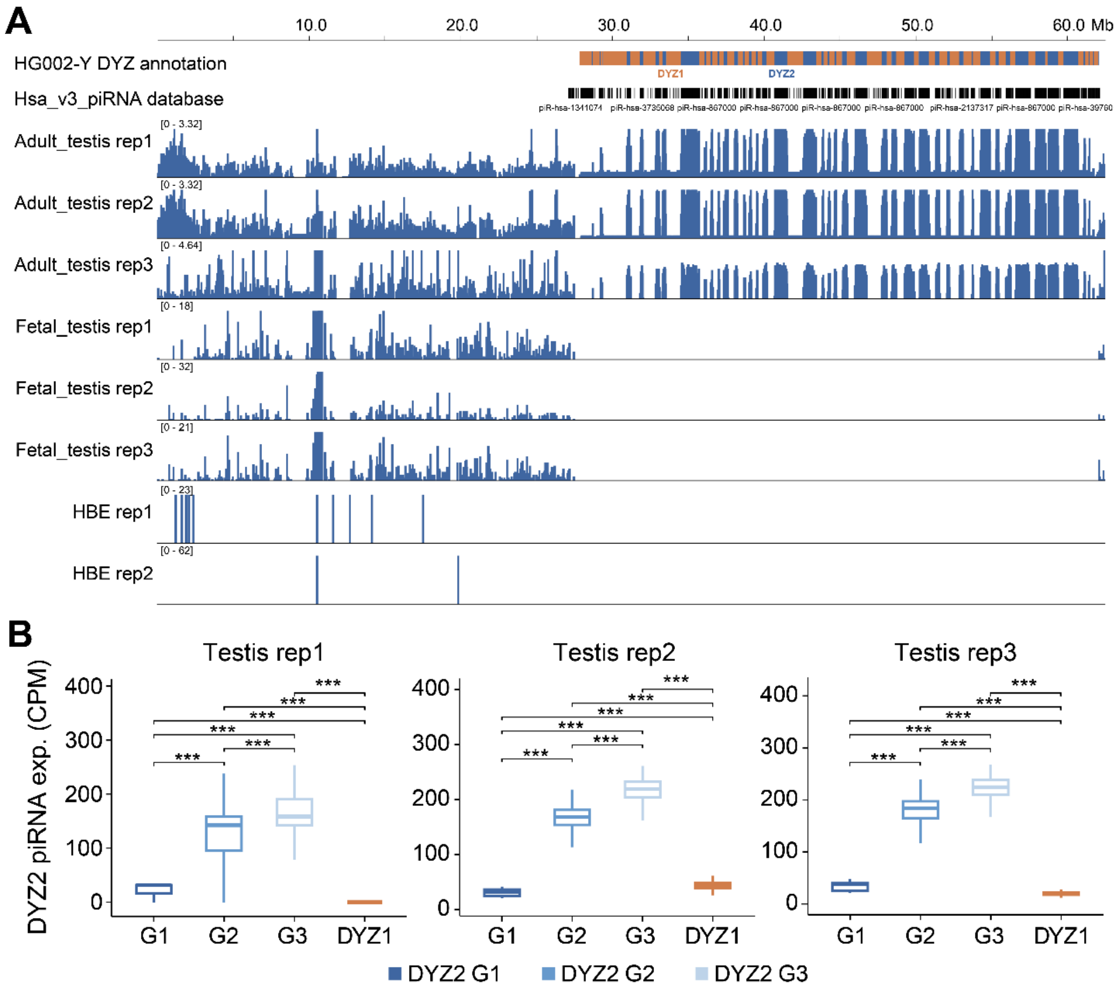


**Figure S15. The piRNA expression among DYZ2 subgroups**.

**A)** The piRNA read depth for different tissues. HBE: human bronchial epithelial. **B)** The piRNA expression (CPM, Counts Per Million) among DYZ2 subgroups for different adult testis replicates. ***: $P < 2.22e-16$.

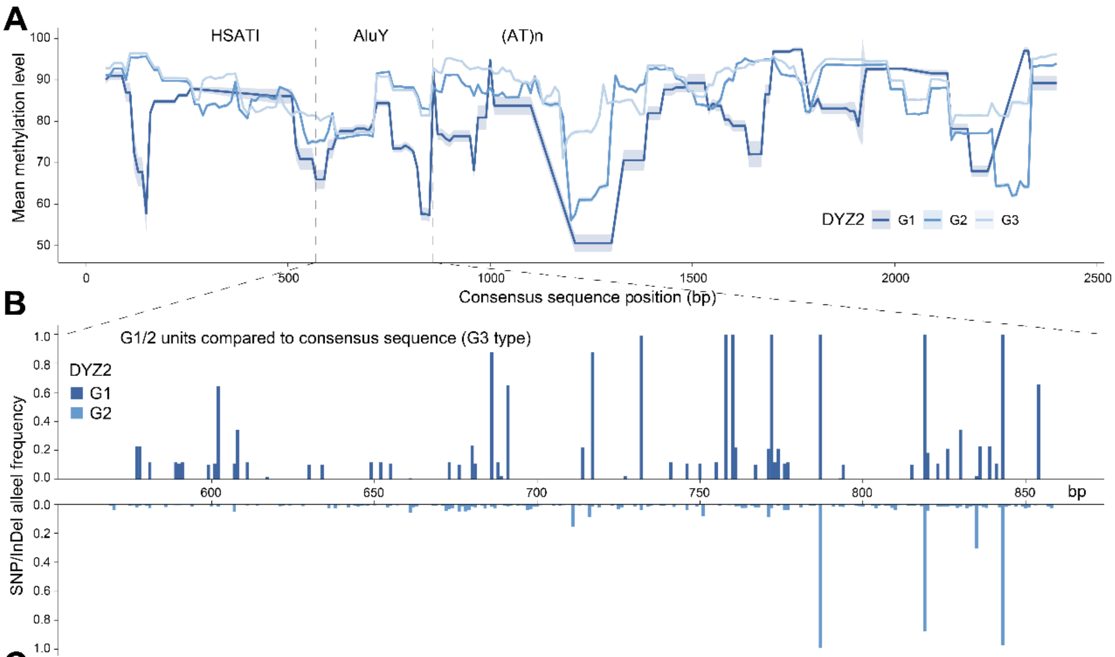


C

| Motif | Type | Start | Stop | Strand | Score | P-value | Q-value | Sequence |
|---|---|---|---|---|---|---|---|---|
| ZN467 | DYZ2_Alu_G1 | 27 | 48 | + | 9.83673 | 3.49E-05 | 0.0168 | TTGGAGGGCAAGGCAAGGGGAT |
| ZN281 | DYZ2_Alu_G1 | 177 | 191 | + | 10.3182 | 3.53E-05 | 0.0173 | GGGAGGCTGAGGGGG |
| COT2 | DYZ2_Alu_G1 | 180 | 196 | + | 11.6636 | 4.05E-05 | 0.0214 | AGGCTGAGGGGGCACAA |
| PLAL1 | DYZ2_Alu_G1 | 185 | 194 | + | 11.4679 | 4.61E-05 | 0.0242 | GAGGGGGCAC |
| BRAC | DYZ2_Alu_G1 | 162 | 181 | - | 11.0714 | 6.03E-05 | 0.0321 | CTCCCAGGTAGCTGGGAGTA |
| HXA7 | DYZ2_Alu_G1 | 192 | 206 | - | 10.8416 | 6.67E-05 | 0.0209 | GTTCAAGCGATTGTG |
| SALL4 | DYZ2_Alu_G3 | 251 | 260 | - | 12.6633 | 2.50E-05 | 0.0132 | ACAGGGTGGG |
| KLF5 | DYZ2_Alu_G3 | 245 | 258 | - | 11.8485 | 3.44E-05 | 0.0171 | AGGGTGGGGTACAA |
| SOX8 | DYZ2_Alu_G3 | 214 | 229 | - | 11.3804 | 4.04E-05 | 0.022 | TCACTGCAACTTCTGC |
| GLIS3 | DYZ2_Alu_G3 | 248 | 257 | - | 11.8427 | 4.68E-05 | 0.0247 | GGGTGGGGTA |

**Figure S16. The DNA methylation (blood), differentiated sequence and banding motifs of DYZ2 sub-groups.**

**A)** Aggregated DNA methylation landscape of DYZ2 sub-group units for 160Ys samples. The base-pair methylation levels were derived from the blood ONT reads. **B)** Differentiated sites of DYZ2 *AluY* of G1 and G2 groups compared to consensus sequence which is derived from the G1 group. G1 exhibits multiple highly differentiated sites while G2 shows three near-fixed single-nucleotide sites in the *AluY* regions compared to G3 consensus sequence. **C)** Sub-group specific motifs in DYZ2 *AluYs* using FIMO program from the MEME Suite[14], underlying potential function differences due to sequence divergence.

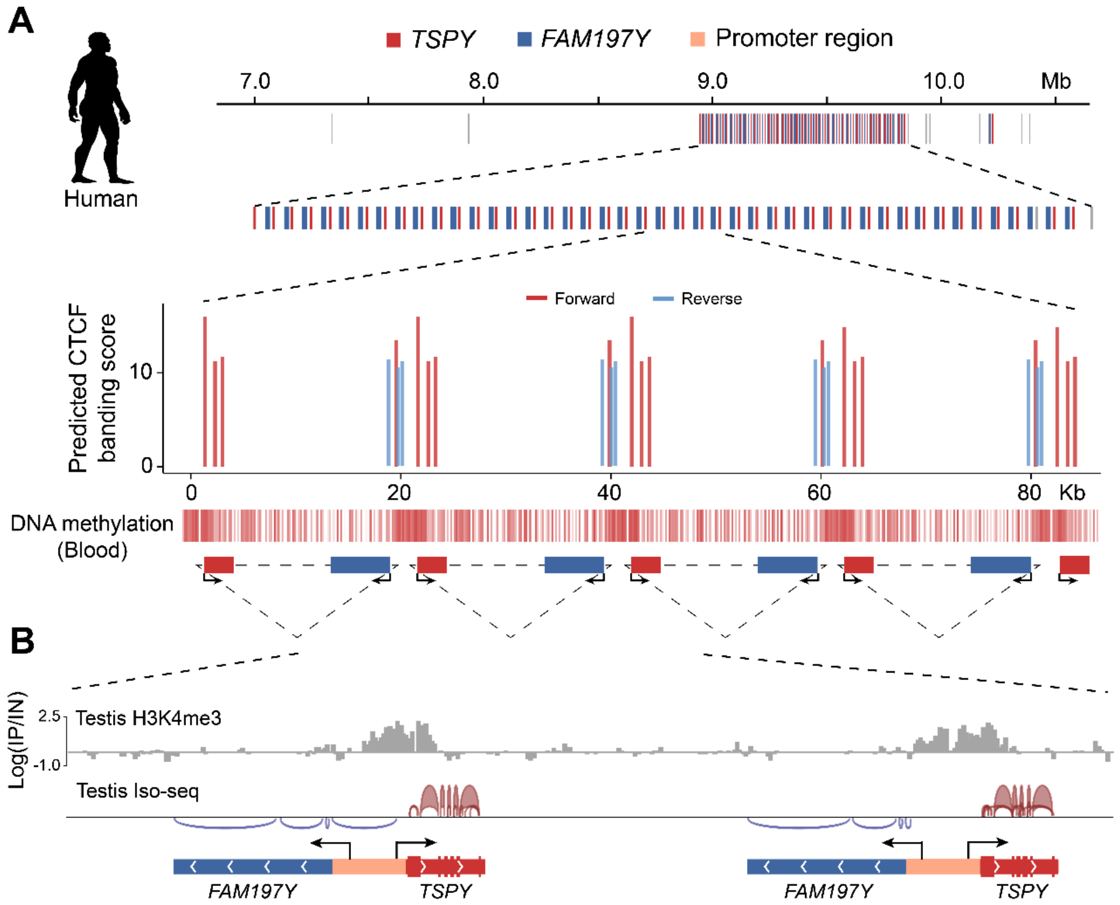


**Figure S17. The organization of *TSPY/FAM197Y* units in human Y.**

**A)** The upper panel suggests the *TSPY/FAM197Y* unit could form a 'TAD' (topologically associating domain) like structure with reverse CTCF (CCCTC-binding factor) binding sequences located between two genes. Their boundaries exhibit high DNA methylation (blood) levels and H3K4me3 modification (testis, **B**). The transcription orientations are also different between the adjacent genes.

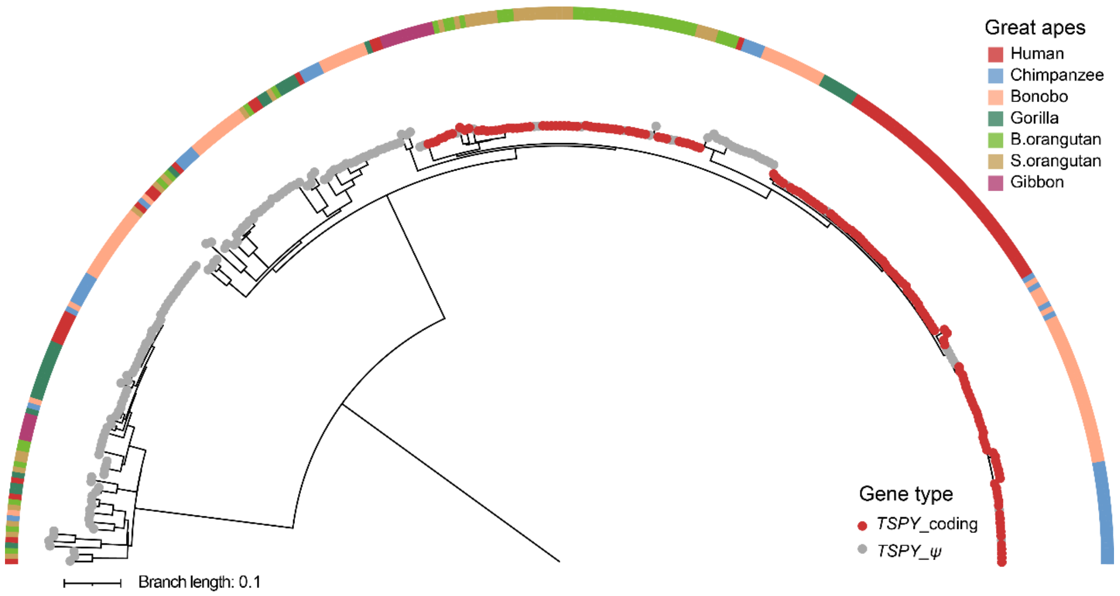


**Figure S18. The phylogenetic tree of nucleic acid of all *TSPY* genes across great ape species.**

Functional protein-coding genes are clearly separated from pseudogenes (*TSPY_ψ*). For the protein-coding *TSPYs*, genes from each ape species form distinct clusters.

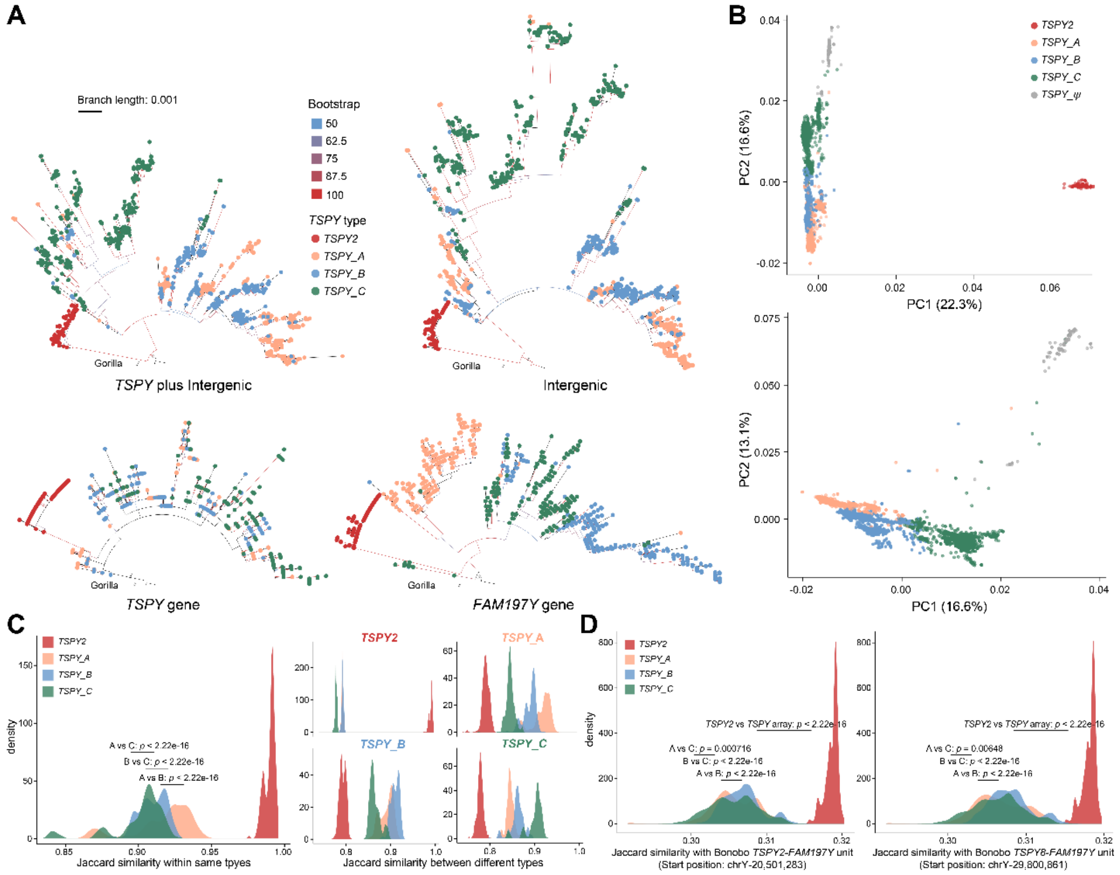


**Figure S19. The classification of *TSPY* unit subgroups.**
**A)** The four *TSPY* sub-clusters are clearly resolved in most phylogenies, except for the tree based solely on *TSPY* gene sequences. This discrepancy underscores the necessity of including flanking regions for robust phylogenetic reconstruction. While the distinction between *TSPY_B* and *TSPY_C* remains ambiguous in the '*TSPY* plus Intergenic' and 'Intergenic' trees, *TSPY2* and *TSPY_A* are consistently recovered as distinct clades. **B)** PCA was performed on all *TSPY/FAM197Y* units across 206 samples, using variants (SNPs and INDELs) identified relative to the *TSPY2/FAM197Y* reference unit from HG1890 (A0b). The right panel displays the PCA results for arrayed *TSPY* units following the exclusion of *TSPY2* units. **C)** Comparison of k-mer similarity (k = 21) both within and between different types of *TSPY/FAM197Y* units. **D)** Analysis of k-mer similarity between all human *TSPY/FAM197Y* units and two randomly selected bonobo orthologs.

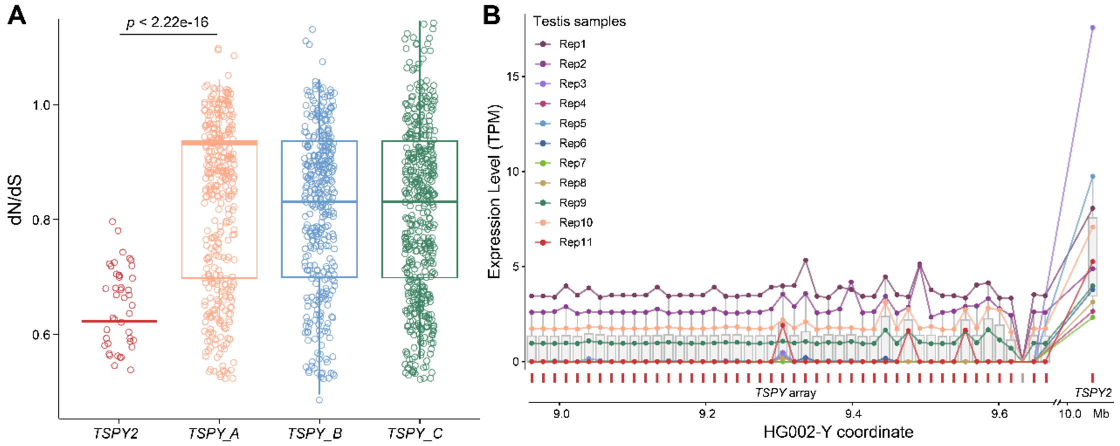


**Figure S20. The sequence and expression difference between *TSPY2* and *TSPY1* genes.**

**A)** The *dN/dS* values of human *TSPY* genes estimated using the free-ratio model implemented in PAML[15], with gorilla *TSPY* genes used as the outgroup. **B)** *TSPY* gene expression levels in human testis RNA-seq replicates. Each dot represents one protein-coding *TSPY* gene, ordered according to its genomic position on HG002-Y.

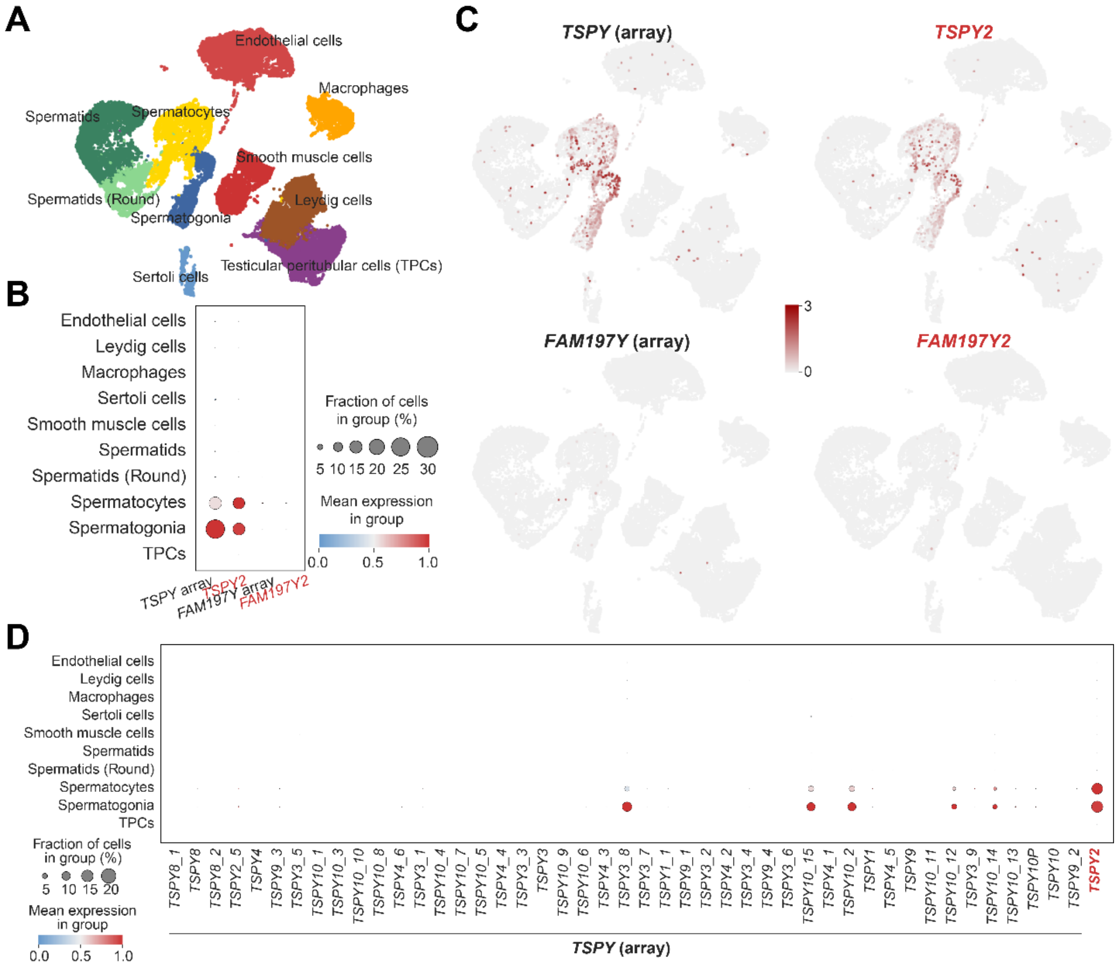


**Figure S21. The *TSPY* and *FAM197Y* gene expression in the testis scRNA data (*n* = 6).**

To avoid the default filtering of multi-mapped reads in canonical scRNA-seq analyses (e.g., the Cell Ranger pipeline), the scTE pipeline was adopted for the expression count of *TSPY* genes (**METHODS**). **A-C)** *TSPY* genes are predominantly expressed in spermatogonia and spermatocyte cells. Note that the expression of the *TSPY* array represents the aggregated expression of all the arrayed genes, and the same applies to the corresponding *FAM197Y* genes. By contrast, *TSPY2* and *FAM197Y2* represent standalone gene units. **D)** Expression levels of individual *TSPY* genes. It should be noted that read cross-mapping may occur among genes within the *TSPY* array.

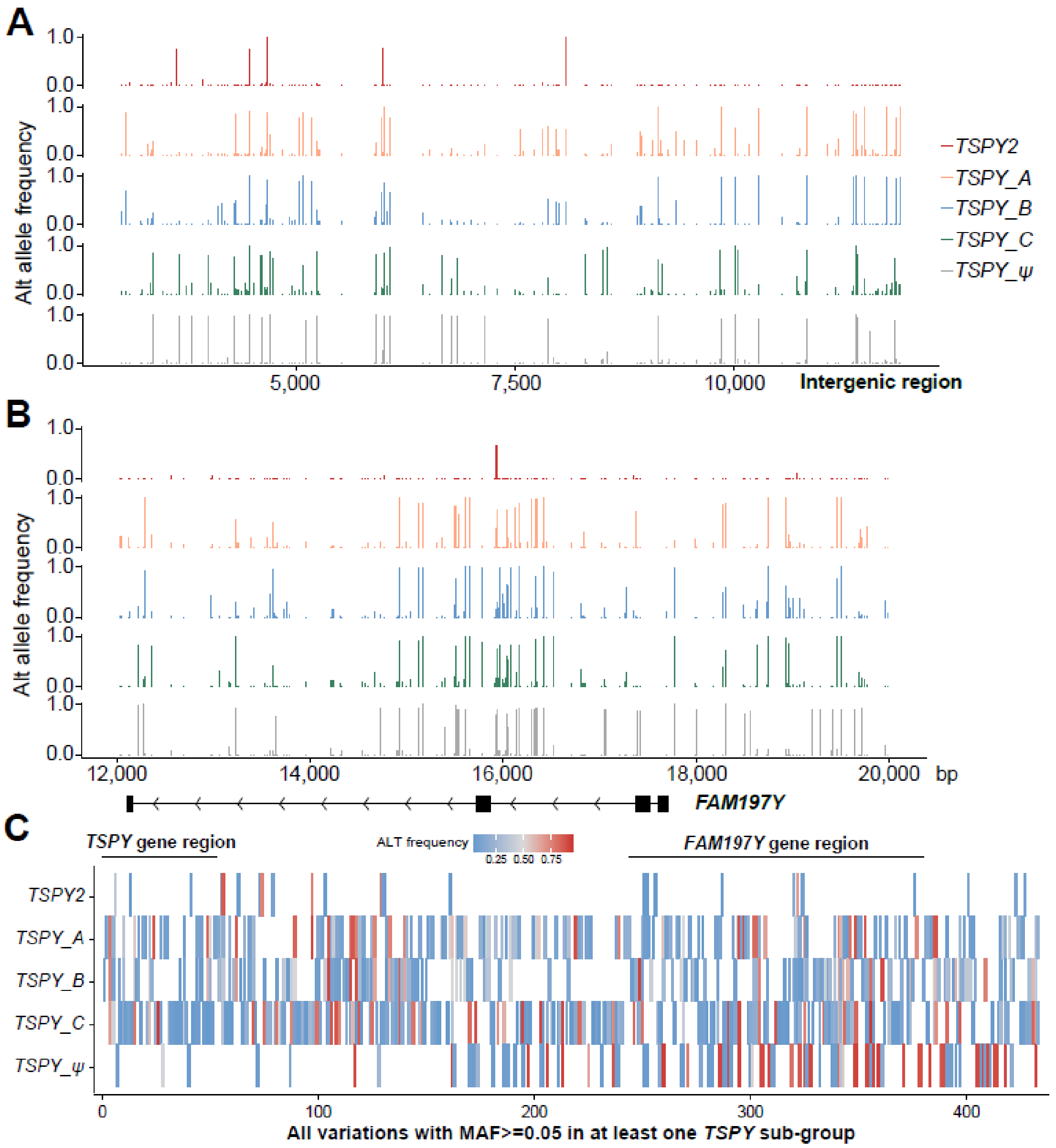


**Figure S22. The alternative (ALT) allele frequency of *TSPY-FAM197Y* sub-regions.**

**A)** Alternative allele (ALT) frequencies in the intergenic regions of *TSPY-FAM197Y* units, using the HG01890 *TSPY2* unit as the reference. **B)** ALT allele frequencies across *FAM197Y* sub-regions. **C)** Numbers of ALT alleles across *TSPY* sub-groups. Only alleles with a minor allele frequency (MAF) ≥ 0.05 in at least one *TSPY* sub-group were included.

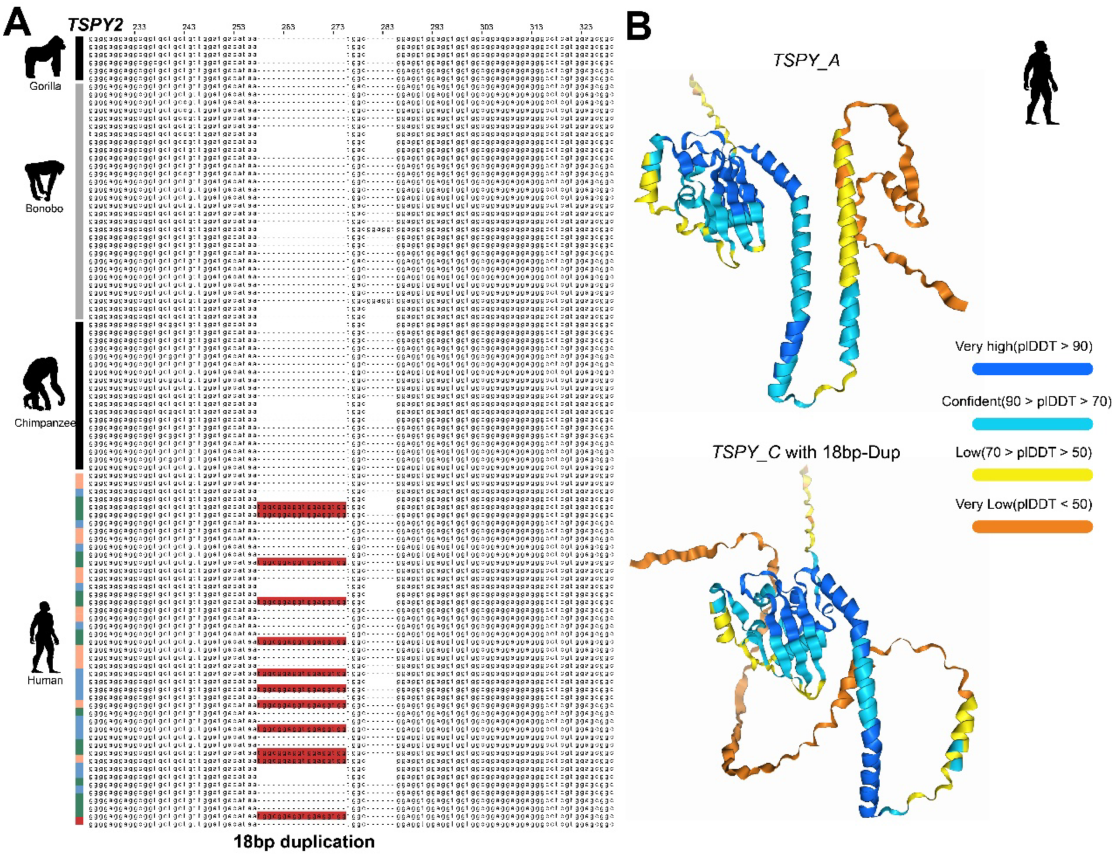


**Figure S23. The sequence differentiation between *TSPY* array genes.**
**A)** An 18-bp duplication occurring at high frequency in *TSPY_C* genes. HG002-Y was used as the representative human Y chromosome. The frequency of this 18bp-duplication is higher in *TSPY_C* (allele frequency: 0.452 vs. 0.157) than in other groups. **B)** This duplication is predicted to cause a significant alteration in the protein three-dimensional structure, as inferred using AlphaFold3[16].

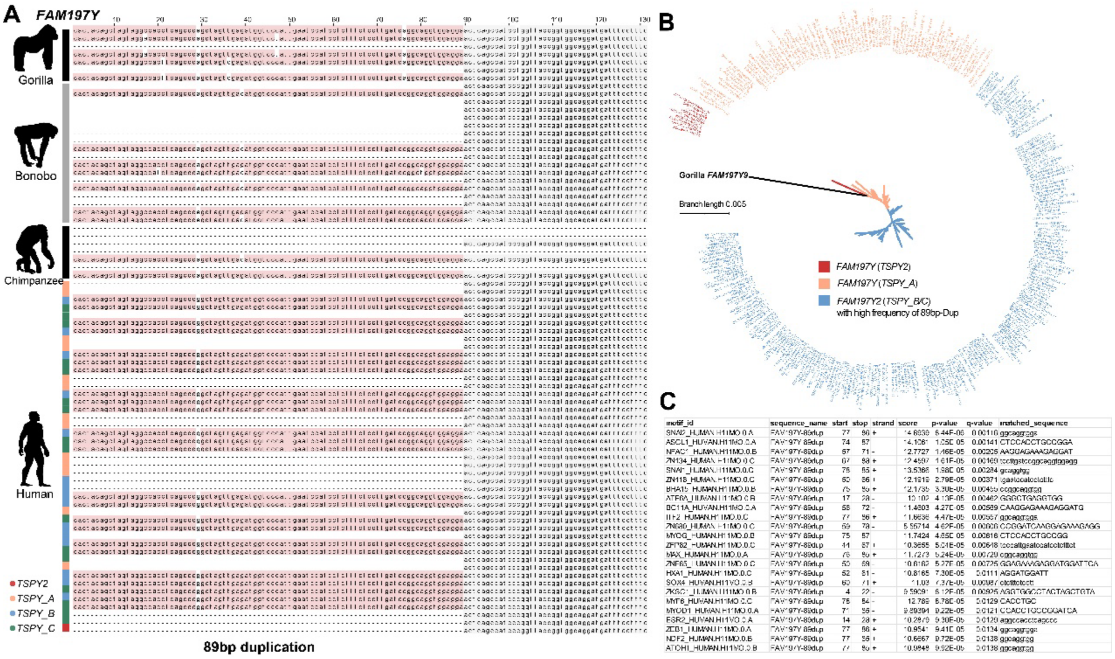


| motif_id | sequence_name | start | stop | strand | score | p-value | q-value | matched_sequence |
|---|---|---|---|---|---|---|---|---|
| SNAI2_HUMAN.H11MO.0.A | FAM197Y-89dup | 77 | 86 | + | 14.6939 | 6.44E-06 | 0.00116 | ggcaggtgga |
| ASCL1_HUMAN.H11MO.0.A | FAM197Y-89dup | 74 | 57 | | 14.1061 | 1.05E-05 | 0.00141 | CTCCACCTGCCGGA |
| NFAC1_HUMAN.H11MO.0.B | FAM197Y-89dup | 57 | 71 | - | 12.7727 | 1.46E-05 | 0.00205 | AAGGAGAAAGAGGAT |
| ZN134_HUMAN.H11MO.0.C | FAM197Y-89dup | 67 | 88 | + | 12.4597 | 1.61E-05 | 0.00160 | tccttgatccggcaggtggagg |
| SNAI1_HUMAN.H11MO.0.C | FAM197Y-89dup | 78 | 85 | + | 13.5366 | 1.98E-05 | 0.00284 | gcaggtgg |
| ZN118_HUMAN.H11MO.0.C | FAM197Y-89dup | 50 | 66 | + | 12.1919 | 2.79E-05 | 0.00371 | tgaatccatcctcttc |
| BHA15_HUMAN.H11MO.0.B | FAM197Y-89dup | 75 | 86 | + | 12.1735 | 3.30E-05 | 0.00455 | ccggcaggtgg |
| ATF6A_HUMAN.H11MO.0.B | FAM197Y-89dup | 17 | 28 | - | 10.102 | 4.13E-05 | 0.00462 | GGGCTGAGGTGG |
| BC11A_HUMAN.H11MO.0.A | FAM197Y-89dup | 58 | 72 | - | 11.4603 | 4.27E-05 | 0.00589 | CAAGGAGAAAGAGGATG |
| ITF2_HUMAN.H11MO.0.C | FAM197Y-89dup | 77 | 86 | + | 11.6636 | 4.47E-05 | 0.00557 | ggcaggtgga |
| ZN680_HUMAN.H11MO.0.C | FAM197Y-89dup | 59 | 78 | - | 5.55714 | 4.62E-05 | 0.00606 | CCGGATCAAGGAGAAAGAGG |
| MYOG_HUMAN.H11MO.0.B | FAM197Y-89dup | 75 | 87 | | 11.7424 | 4.65E-05 | 0.00616 | CTCCACCTGCCGG |
| ZFP82_HUMAN.H11MO.0.C | FAM197Y-89dup | 44 | 67 | + | 10.3655 | 5.01E-05 | 0.00648 | tcccattggatccatcctctttct |
| MAX_HUMAN.H11MO.0.A | FAM197Y-89dup | 76 | 86 | + | 11.7273 | 5.24E-05 | 0.00729 | cggcaggtgg |
| ZNF65_HUMAN.H11MO.0.C | FAM197Y-89dup | 50 | 69 | - | 10.6162 | 5.27E-05 | 0.00725 | GGAGAAAGAGGATGGATTCA |
| HXA1_HUMAN.H11MO.0.C | FAM197Y-89dup | 52 | 61 | - | 10.8165 | 7.30E-05 | 0.0111 | AGGATGGATT |
| SOX4_HUMAN.H11MO.0.B | FAM197Y-89dup | 60 | 71 | + | 11.03 | 7.37E-05 | 0.00987 | ctctttctcctt |
| ZKSC1_HUMAN.H11MO.0.B | FAM197Y-89dup | 4 | 22 | - | 9.59091 | 8.12E-05 | 0.00925 | AGGTGGCCTACTAGCTGTA |
| MYF6_HUMAN.H11MO.0.C | FAM197Y-89dup | 78 | 84 | - | 12.789 | 8.78E-05 | 0.0129 | CACCTGC |
| MYOD1_HUMAN.H11MO.0.A | FAM197Y-89dup | 71 | 88 | - | 9.89394 | 9.22E-05 | 0.0121 | CCACCTGCCCGGATCA |
| ESR2_HUMAN.H11MO.0.A | FAM197Y-89dup | 14 | 28 | + | 10.2879 | 9.30E-05 | 0.0120 | aggccacctcagccc |
| ZEB1_HUMAN.H11MO.0.A | FAM197Y-89dup | 77 | 86 | + | 10.9541 | 9.41E-05 | 0.0134 | ggcaggtgga |
| NDF2_HUMAN.H11MO.0.B | FAM197Y-89dup | 77 | 85 | + | 10.6667 | 9.72E-05 | 0.0138 | ggcaggtgg |
| ATOH1_HUMAN.H11MO.0.B | FAM197Y-89dup | 77 | 86 | + | 10.9848 | 9.92E-05 | 0.0138 | ggcaggtgg |

**Figure S24. The sequence differentiation between *FAM197Y* genes.**

**A)** An 89-bp duplication in the promoter region of *FAM197Y2* genes (highlighted in red), which are linked to *TSPY_B/C* genes. **B)** The maximum-likelihood (ML) phylogenetic tree of *FAM197Y* genes in HG002-Y reveals three distinct clusters corresponding to the *TSPY* sub-clusters. **C)** The duplicated 89-bp sequence is enriched in binding motifs, suggesting potential functional relevance.

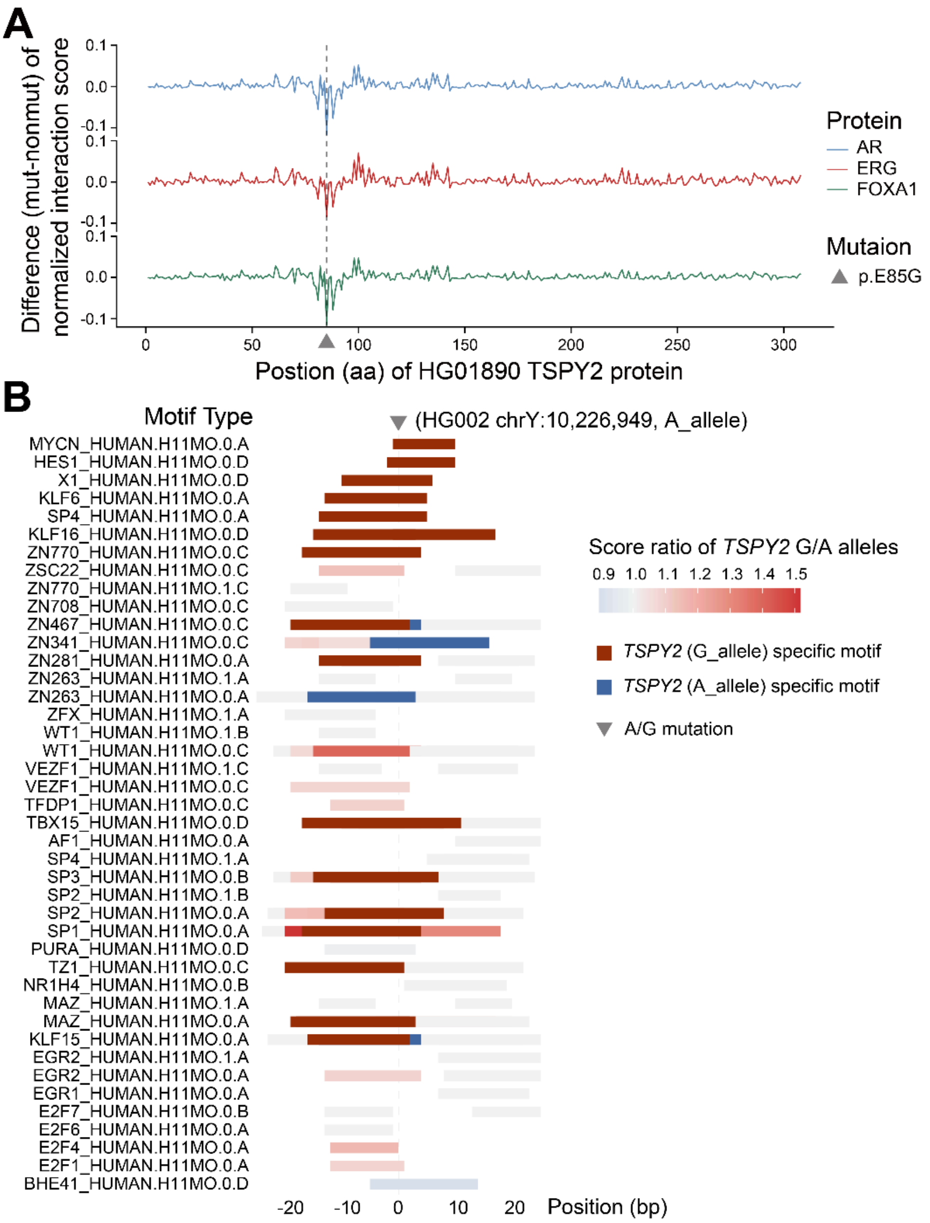


**Figure S25. The mutations which could affect the affinity of TSPY with other proteins.**

**A)** The amino acid mutation (p.E85G) of TSPY2 protein could decrease the interaction with other proteins, e.g., the TFs related to prostate cancer (*AR*, *ERG* and *FOXA1*). The Y-axis represents the delta of interaction scores comparing the mutated (G) to wild (E) amino acids. **B)** The DNA mutation (A>G) could alter the binding affinity for transcription factors and induce new binding motifs. The enrichment score ratios were compared between A/G alleles for the shared motifs.

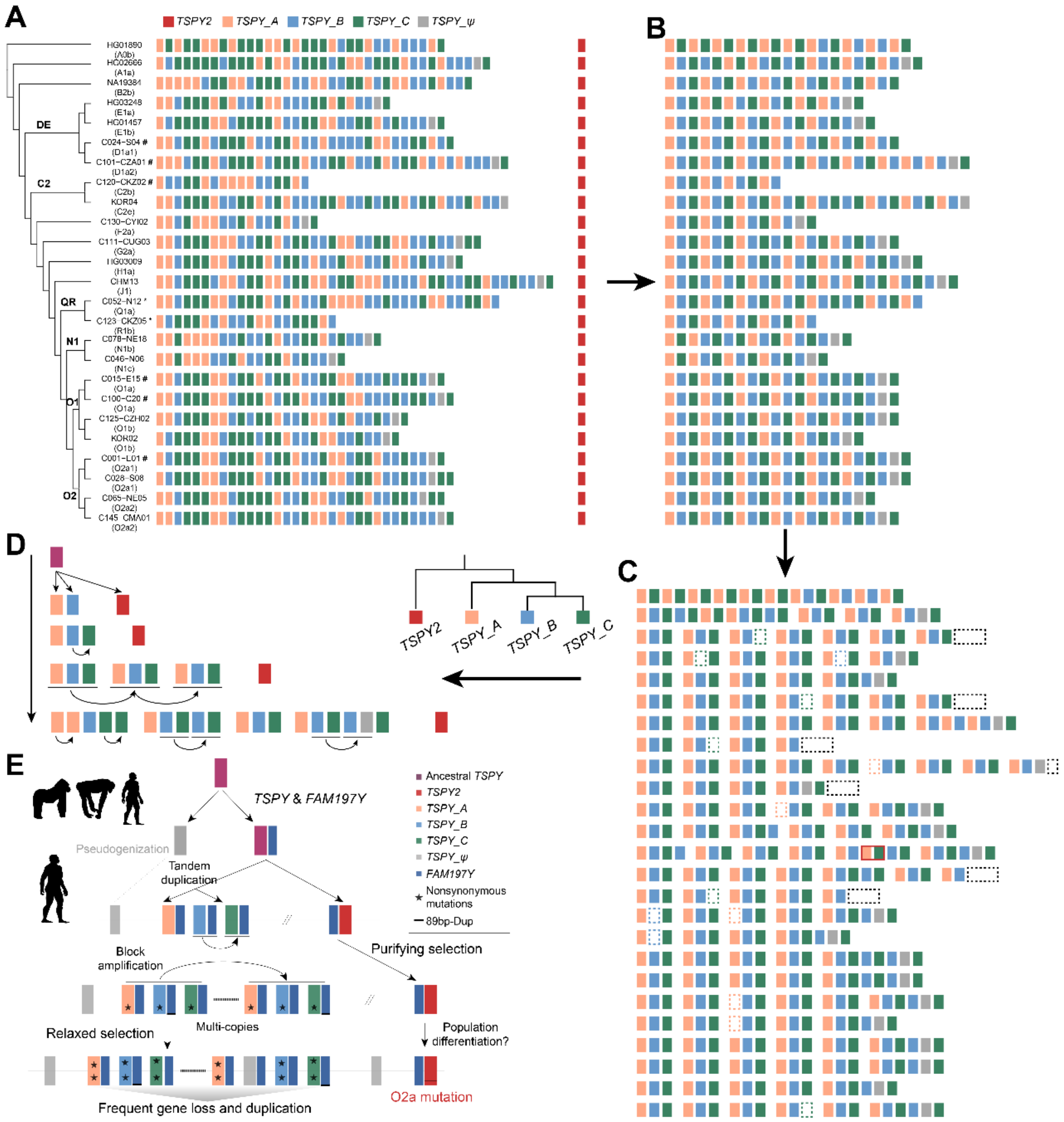


**Figure S26. The model of *TSPY* gene evolution.**
**A)** Gene order of all *TSPY* copies for each representative haplogroup. The symbol "#" indicates samples in which gene order was adjusted due to nested inversions (*INV_1* and *INV_2*) spanning AMPL1 and AMPL2 (see Figure 4A). **B)** Genes were compacted by collapsing neighboring duplicated genes of the same type. **C)** The duplication unit is most likely composed of *TSPY_A+B+C*. Dashed boxes indicate possible gene conversion or deletion events. **D)** A simplified expansion model of *TSPY* genes according to the *TSPY* phylogeny. **E)** Proposed evolutionary model for the *TSPY* gene family in human Ys. Following the acquisition of *FAM197Y*, the *TSPY-FAM197Y* unit diverged into two oppositely oriented clusters in humans: *TSPY*, forming a tandem array, and *TSPY2*, remaining as a singleton. *TSPY* expanded through tandem duplication and/or block amplification accompanied by relaxed selection, whereas the *TSPY2* sequence largely remained conserved under purifying selection. Frequent gene conversion events may have contributed to sequence homogenization and the functional maintenance of the *TSPY* array.

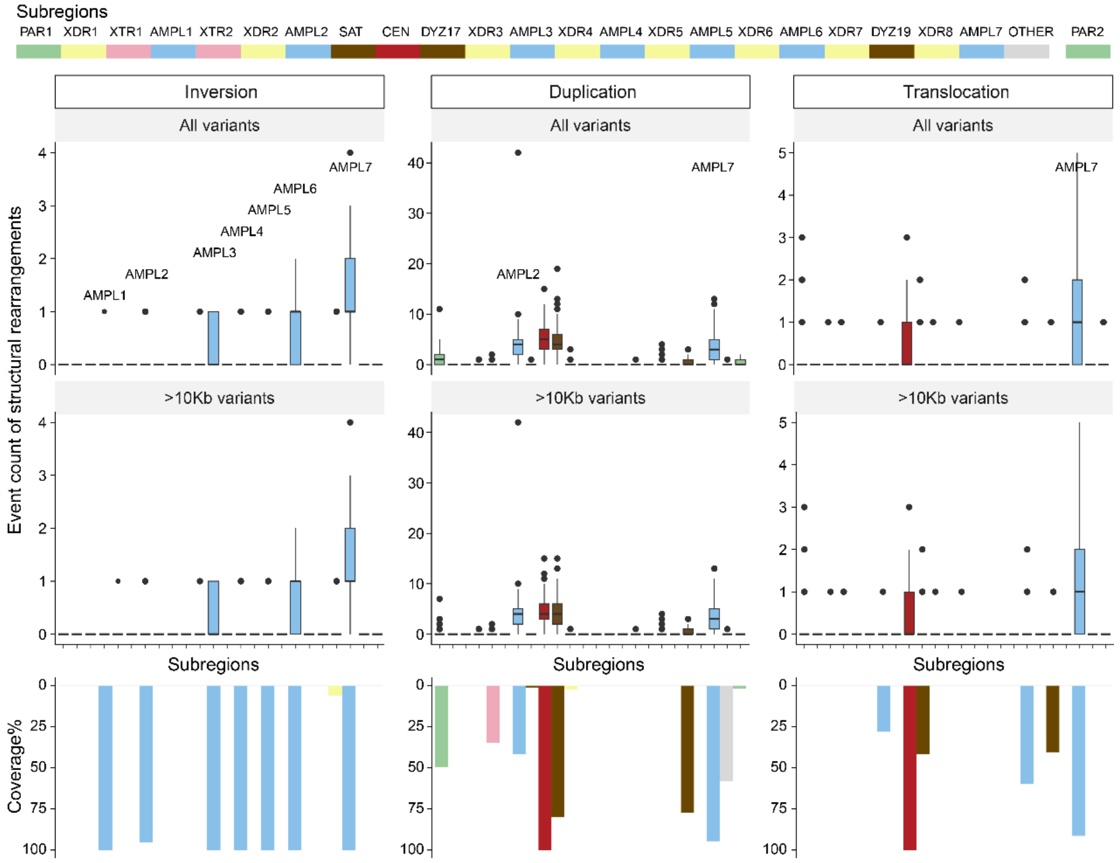


**Figure S27. The structural rearrangement events identified in euchromatic subregions.**

Each dot in the boxplot represents one individual. All inversions and translocations are larger than 10kb. The bar plots indicate the coverage (%) of rearrangements occurring in each subregion. Only non-singleton events were considered.

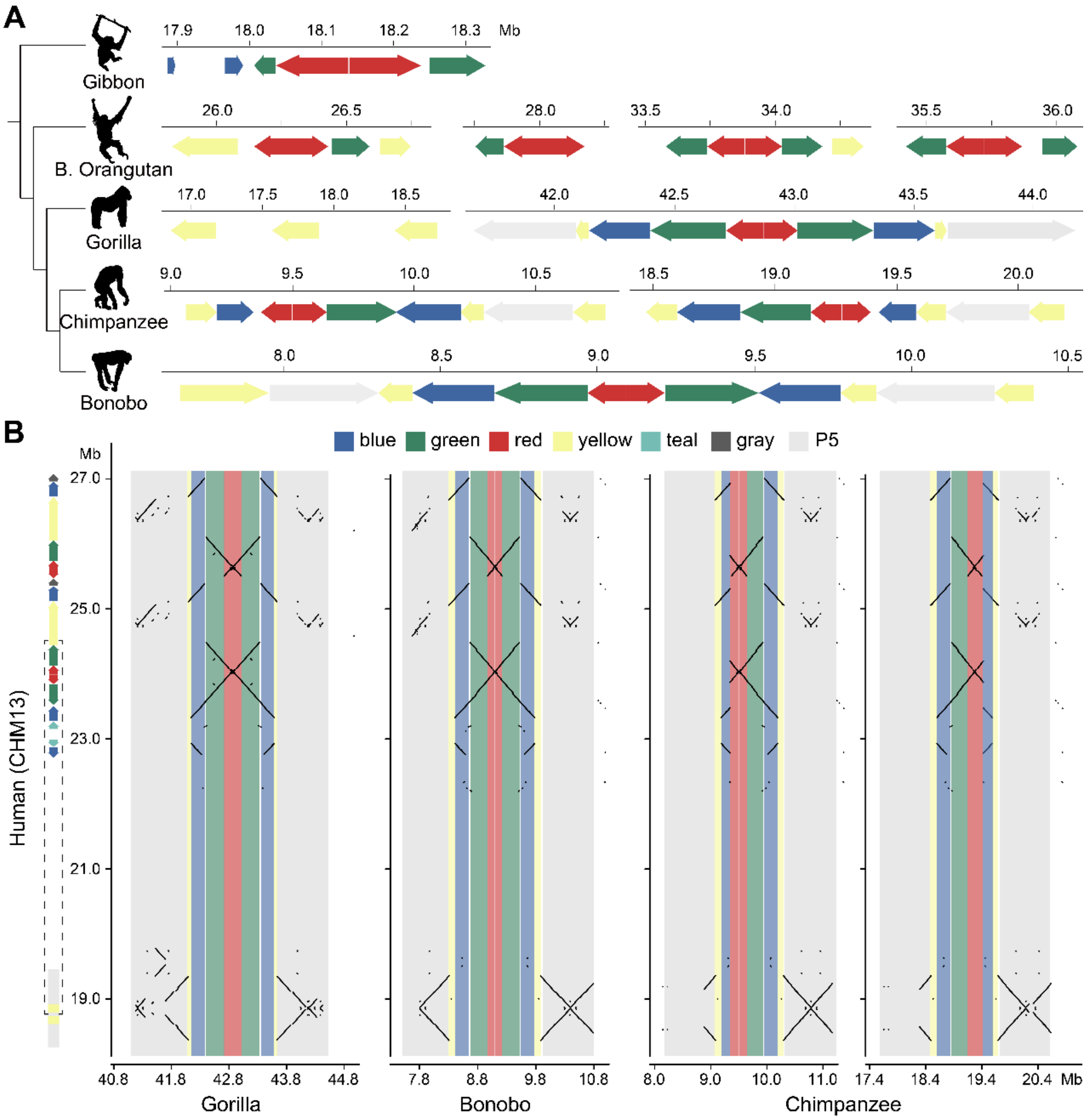


**Figure S28. The origin of amplicons across great apes.**

**A)** The homologous sequences of the human-Y AMPL7 amplicons and palindrome 5 (P5) among the Y chromosomes of great apes. The sequences of red and green amplicons are conserved across apes, whereas blue amplicons originated in the common ancestor of Hominidae. Only partial sequences of human yellow amplicons can be detected in other apes. The flanking sequences of green-bule-red amplicons in apes are homologous to the P5 sequences, which are far away from amplicons in current human Ys. **B**) The synteny of amplicons between humans (Y-axis) and other apes (X-axis). There could be an inversion (black dashed box on the X-axis) occurring between similar sequences of palindrome 5 (P5) and yellow amplicons specifically in the human ancestor. The subsequent duplication and other rearrangements have made large palindromes (P1-P3) in modern humans.

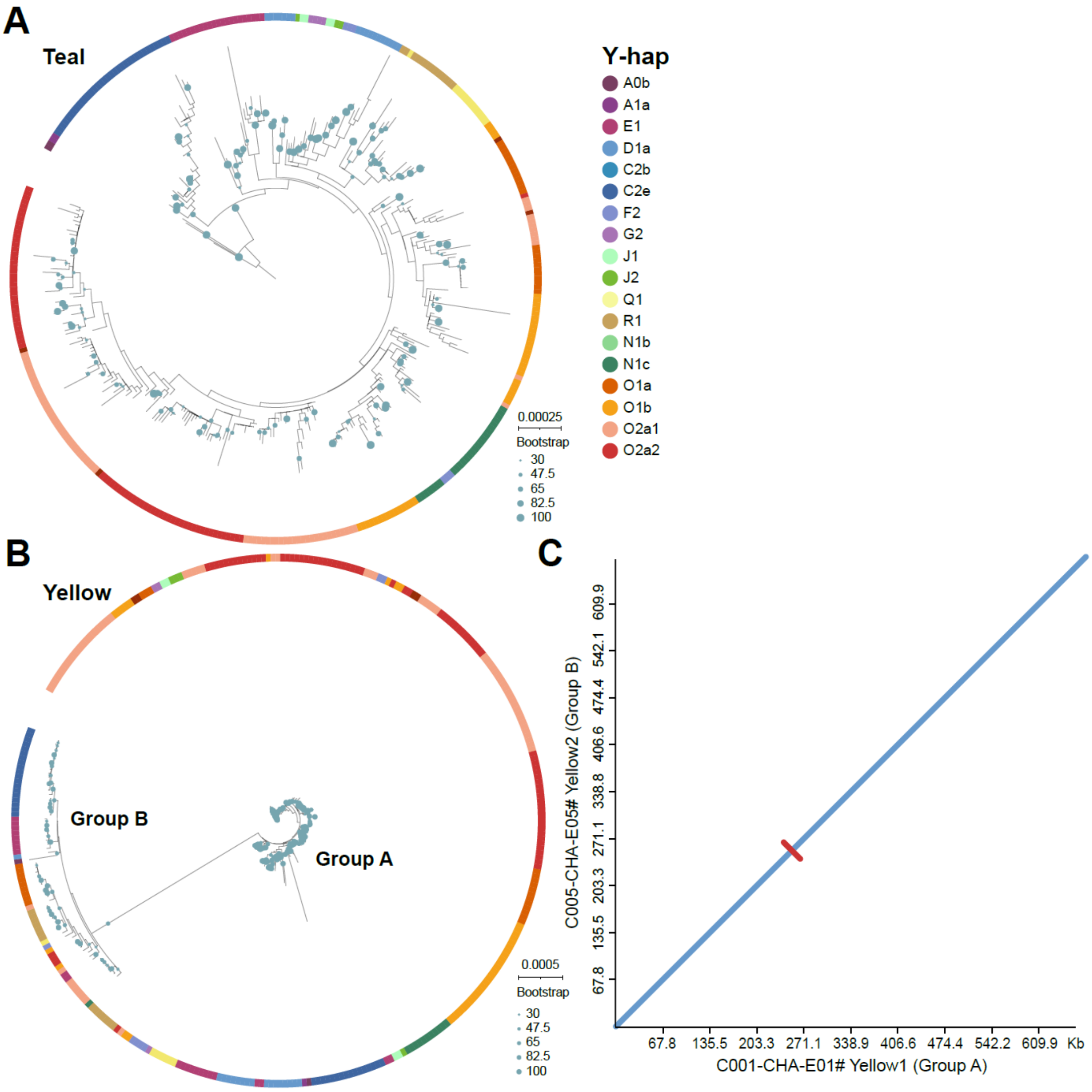


**Figure S29. The phylogenetic trees of teal and yellow amplicons**.
**A-B)** The phylogenetic subclusters for teal and yellow amplicons are not clear as in other amplicons. Both trees were using the ones from the A0b haplogroup as outgroups. **B)** A small group (Group B; *n* = 42) of yellow amplicons are clustered away from the main group (Group A), due to a ~30Kb inversion (red color, **C**) within yellow amplicons.

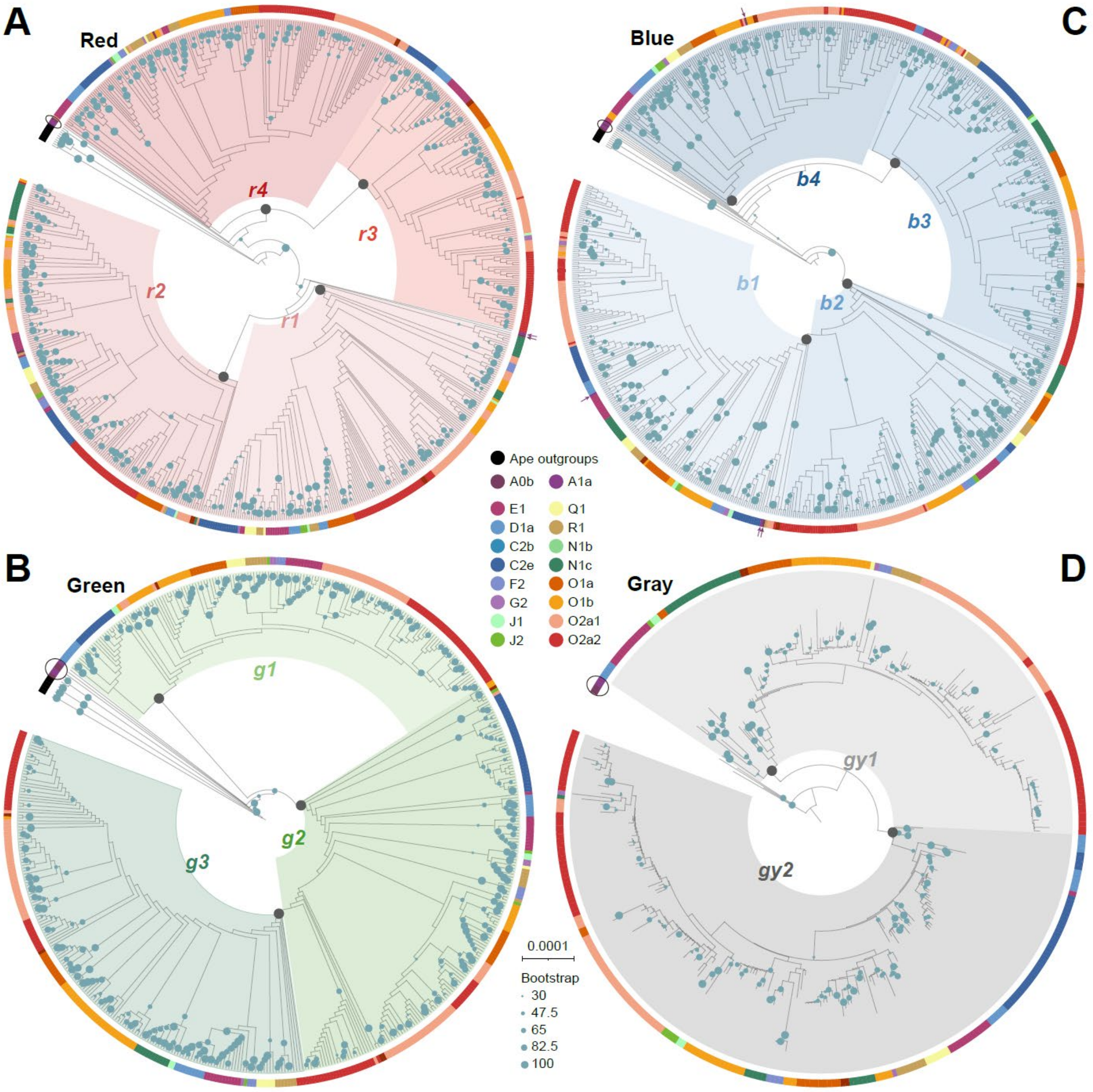


**Figure S30. The phylogenetic trees for red (A), green (B), blue (C) and gray (D) amplicons.**

The first three ML trees were using the gorilla homologous sequences as root outgroups, and the tree of gray amplicons were using the ones from the A0b haplogroup as outgroups. The external bars represent the Y haplogroups. The division of amplicon subtypes, e.g., *r1* and *r2*, seems to occur after the divergence from the two basal Y-haplogroups: A0b (HG01890) and A1a (HG02666). The green dots represent the bootstrap supports, while the gray dots represent the CT ancestral nodes within each subgroup. The circus represent the amplicons from A0/A1 individuals.

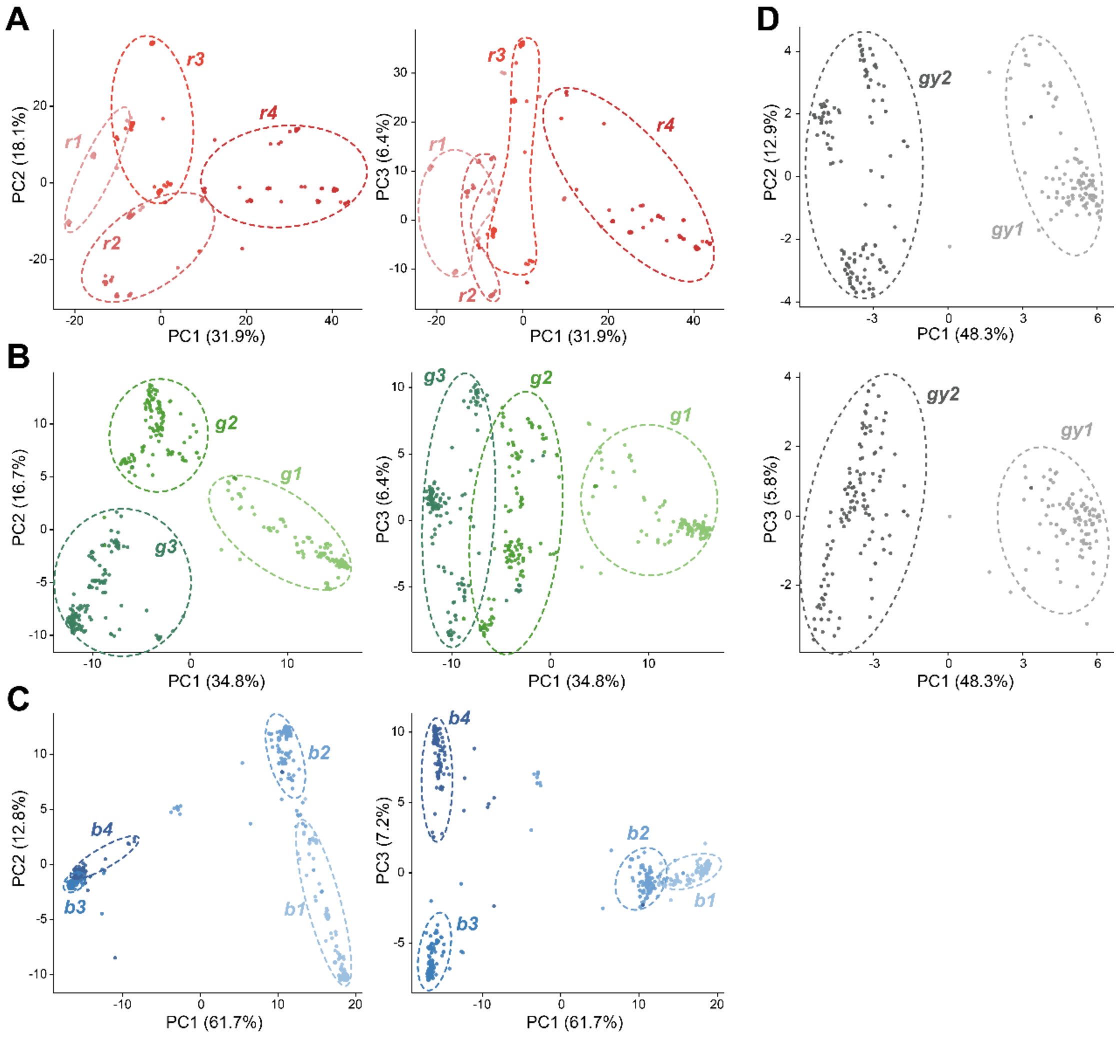


**Figure S31. The PCA of four amplicon sub-groups.**

The PCA results suggest clear sub-clustering for green, blue and gray amplicons, while the red amplicons show more dispersive clustering.

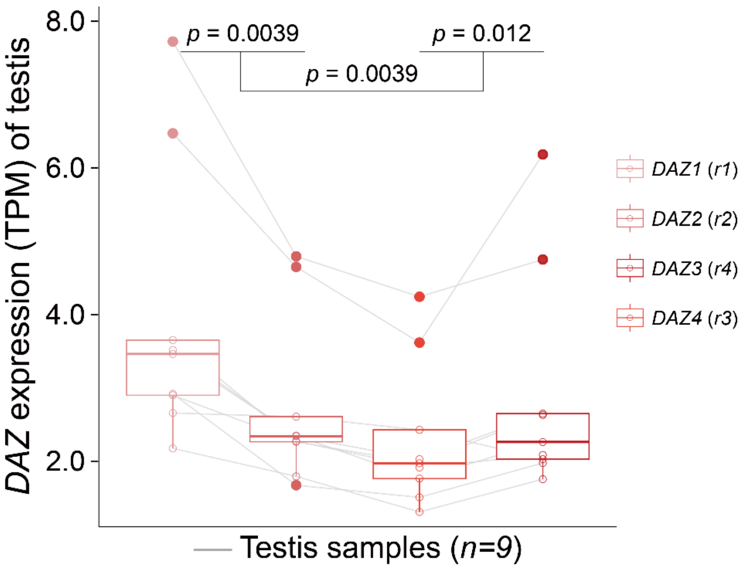


**Figure S32. Differentiated gene repression of *DAZ* gene paralogs.**
The four *DAZ* genes show different expressions in testis, implying functional differentiation could have driven the homologous dominance of amplicons. In HG002-Y reference, *DAZ1* and *DAZ2* genes are located on *r1* and *r2* amplicons, respectively, and *DAZ3/4* are located on *r4/r3*.

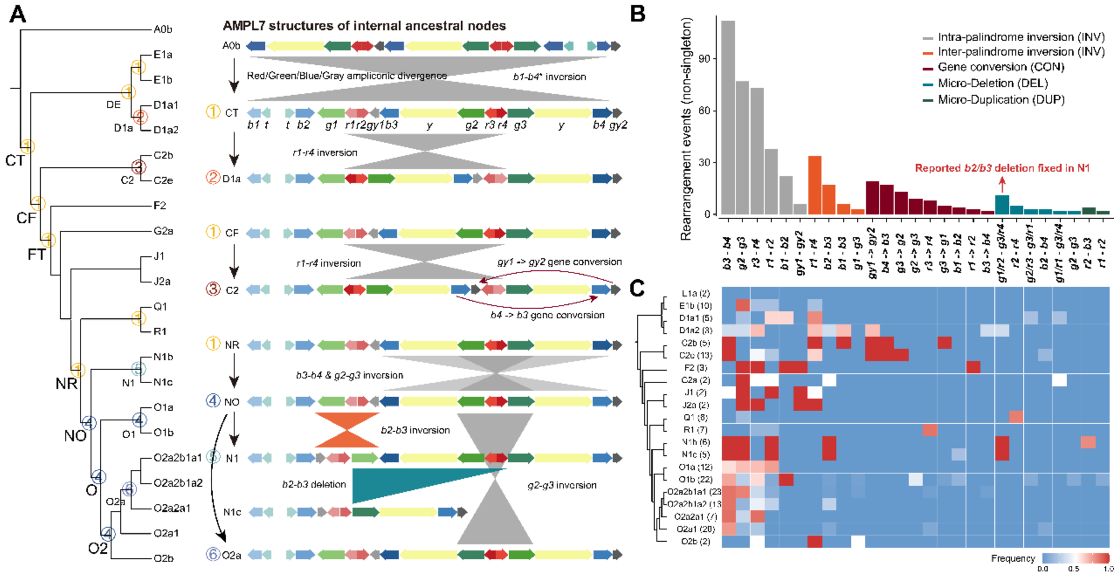


**Figure S33. Rearrangements across Y-haplogroups in AMPL7.**

**A)** Structural rearrangements of representative haplogroups and internal nodes relative to the inferred ancestral haplotype (CT node). The numerical label (1) designates the ancestral configuration for all CT-descendant haplogroups. A large-scale inversion spanning nearly the entire AMPL7 region distinguishes the CT ancestral haplotype from the HG01890 (A0b) lineage. Asterisks indicate unclassified amplicon sub-types within the A0b haplogroup. **B)** Distribution of non-singleton rearrangement events of individuals relative to CT ancestral haplotype. These events include both structural alterations and gene conversions between amplicons. **C)** Frequency of various rearrangement types across major Y-haplogroups. Values in parentheses adjacent to haplogroup labels indicate the respective individual counts.

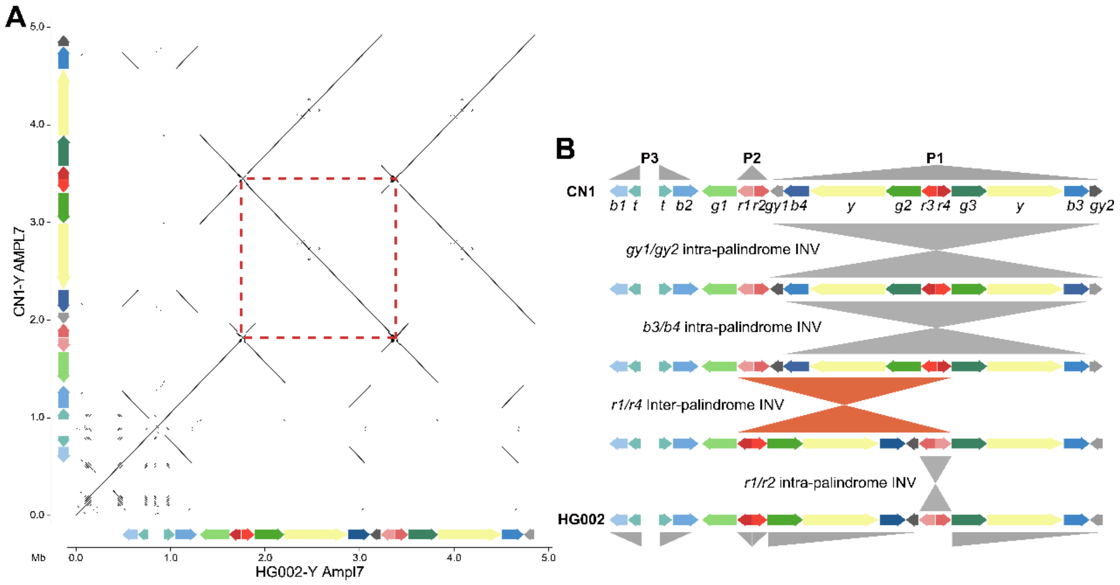


**Figure S34. The rearrangements of AMPL7 between HG002- and CN1-Y.**
**A)** The synteny plot suggests an inversion (red dashed box) between HG002-Y and CN1-Y[11] in the AMPL7 region. **B)** The comparison of refined haplotypes via amplicon classification reveals complex rearrangement steps rather than one simple inversion between two assemblies.

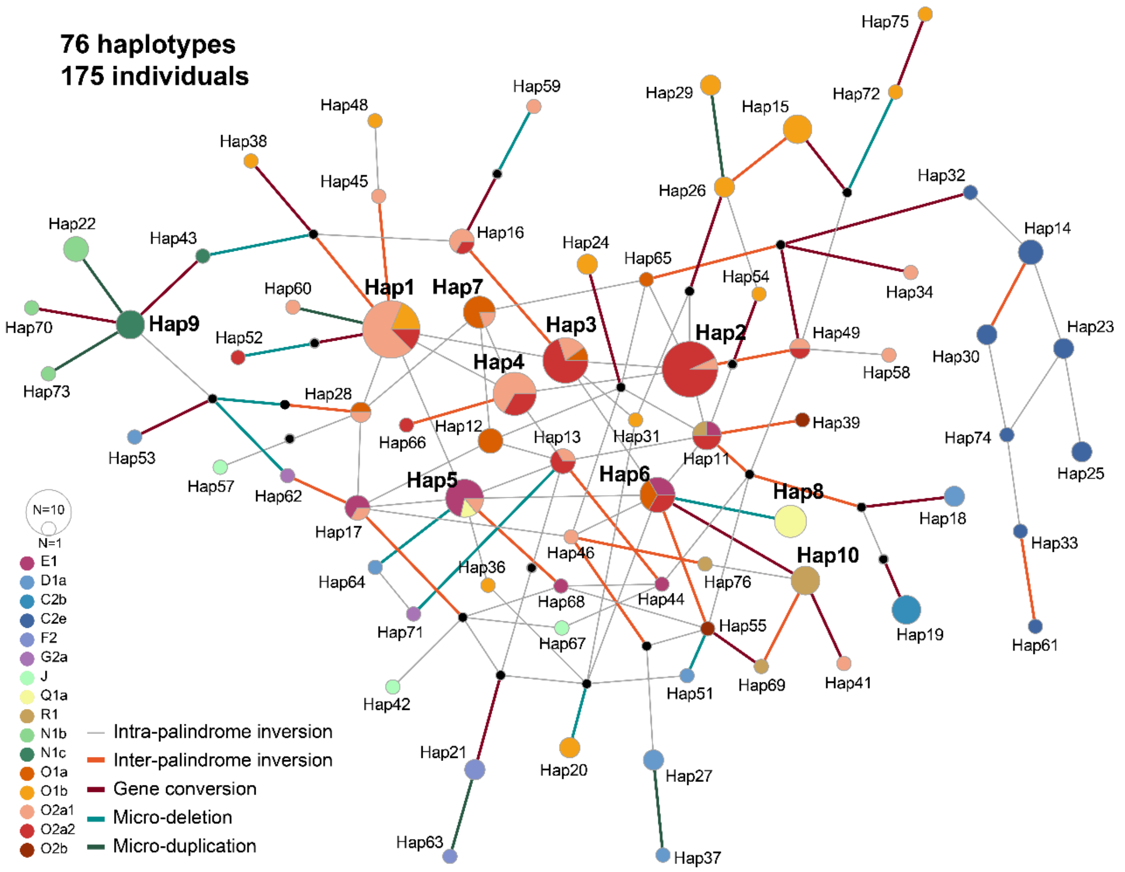


**Figure S35. Haplotype network architecture of the AMPL7 region.**

A total of 76 distinct haplotypes were inferred from 175 individuals with high-quality AMPL7 assemblies. Among these, 30 are non-singleton haplotypes comprising 129 individuals. The complex structural transitions between haplotypes, including intra- and inter-palindromic inversions, gene conversions, micro-deletions, and duplications, are represented by the color-coded connecting lines. Node color denotes the associated Y haplogroups, while node size is proportional to the individual count.

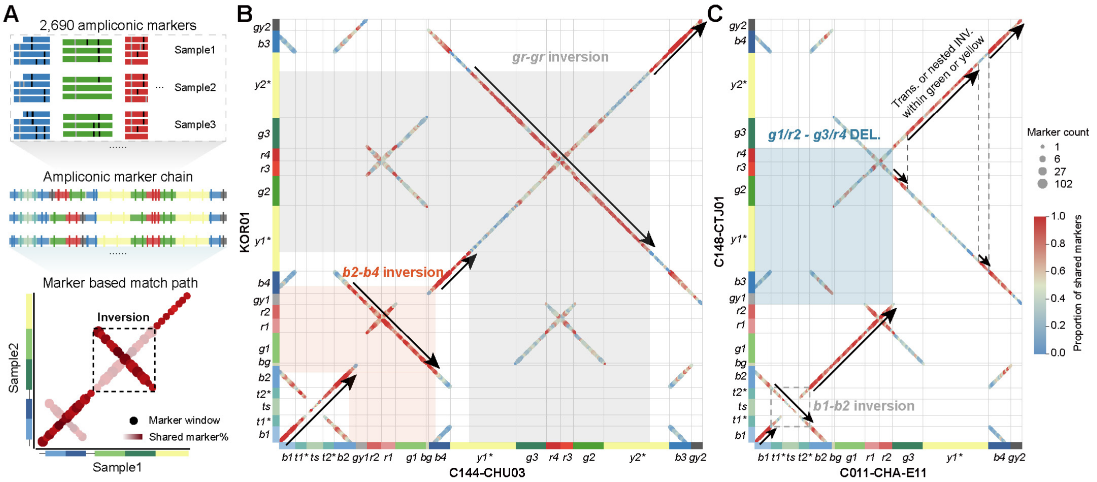


**Figure S36. Validation of AMPL7 structural rearrangements using an allele-marker based matching strategy.**

**A**) Schematic illustration of the marker-based matching approach. Amplicons belonging to the same color class from all samples were projected onto a common reference amplicon to identify allele-specific SNP/INDEL markers. Local shared-marker proportions between samples were calculated to construct orientation-aware matching paths. Two examples (B-C) are presented. **B**) Representative marker-based similarity dotplot validating the inversions between two samples, C144-CHU03 and KOR01. Each dot represents one homologous window, and dot colors represent the proportion of shared allele markers between local windows, with red indicating the highest similarity. Continuous anti-diagonal trajectories indicate inversion events, whereas diagonal trajectories indicate collinear regions. The X/Y axis bars represent the annotated sub-amplicon order in two samples. The marker matching suggests two inversions, *b2-b4* and *gr-gr*, which are consistent with the sub-amplicon synteny in previous analysis, with more precise breakpoint intervals. The black arrows represent the optimal matching path between two samples. The “*” represents the teal and yellow amplicons which cannot be phylogenetically divided into sub-types. The "*ts* " and "*bg*" represent spacer sequences between teals and blue/green amplicons, respectively. **C**) Representative marker-based similarity dotplot validating a *g1/r2-g3/r4* microdeletion between C011-CHA-E11 and C148-CTJ01. The marker trajectories also reveal the complex rearrangements within yellow and green amplicons. The rearrangements can be consistently inferred from both sub-amplcon order and marker matching method, while the latter provides more precise breakpoint intervals.

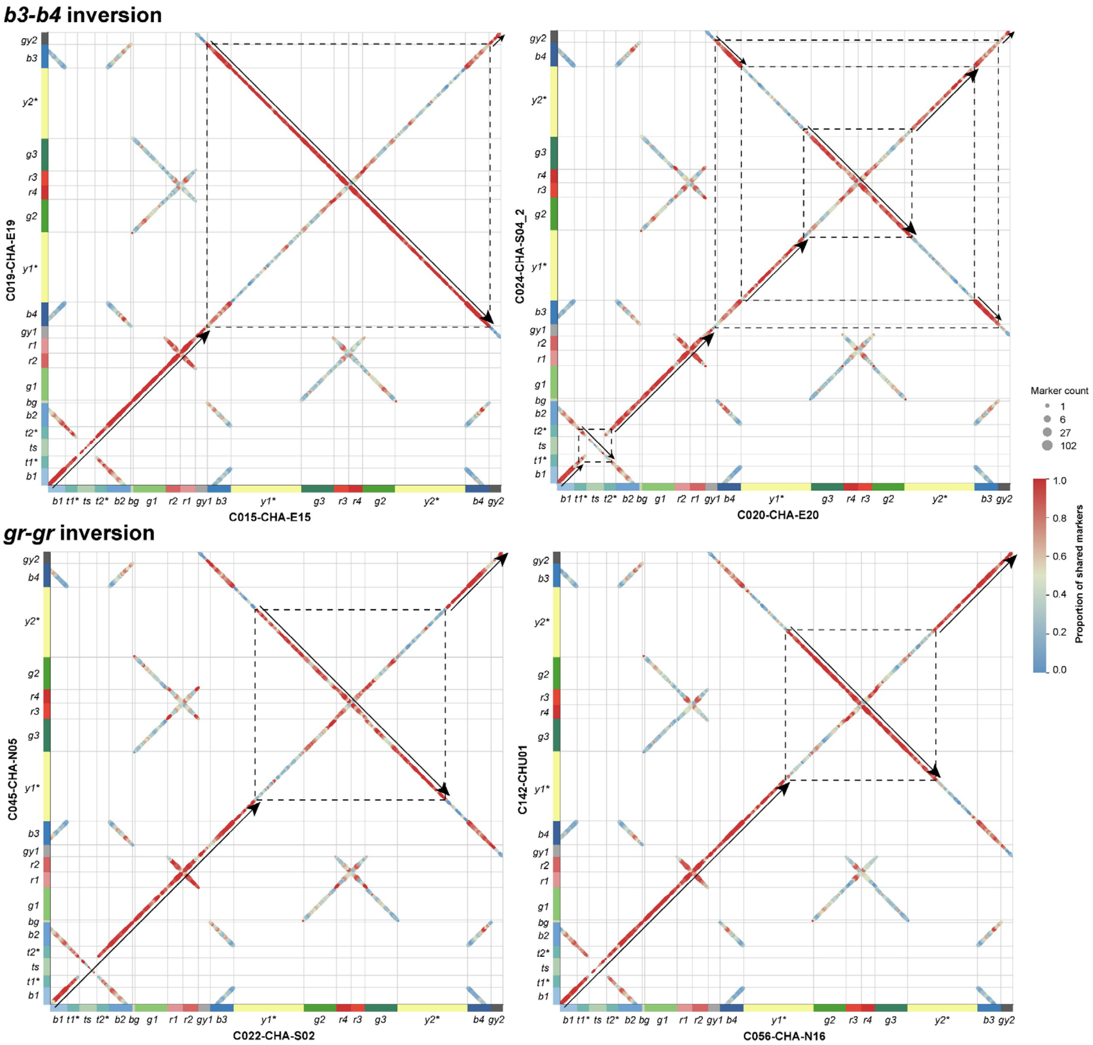


**Figure S37. Marker based alidation of the *b3-b4* and *gr-gr* inversions in AMPL7.** Two examples respecitvely for the *b3-b4* and *gr-gr* inversions. The upper panels represent the two different categories of *b3-b4* inversions. Compared to the left one, the right one exhibits additional translocation or two nested inversions between two yellow amplicons. This breakpoint variations are extensive (check below Figure S38 and SIII39, and related Supplementary materials), indicating more complex structural rearrangements than previously estimated. The below panels represent different breakpoint intervals of *gr-gr* inversions. The bars on the X and Y-axis represent the classified AMPL7 haplotype which was constructed by sub-amplcon order.

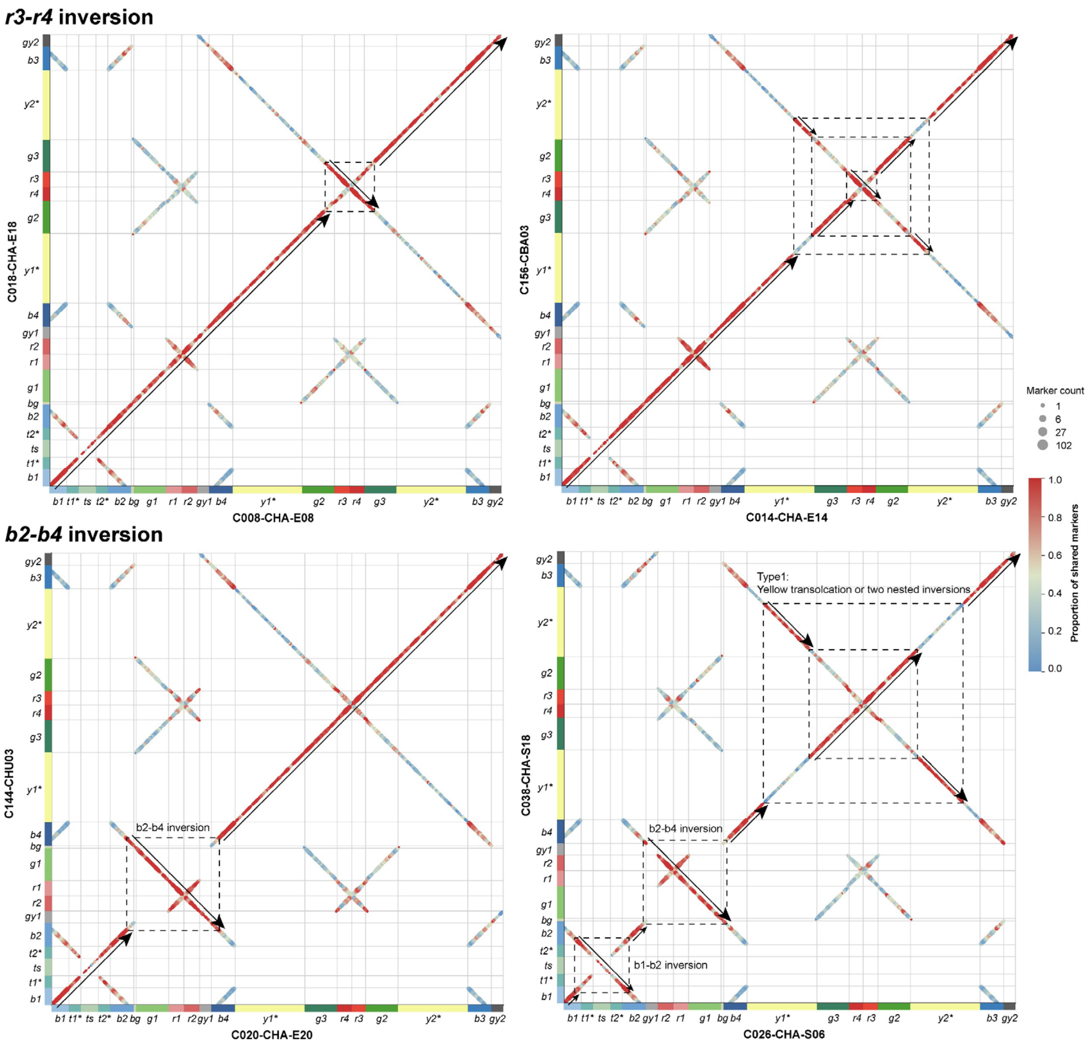


**Figure S38. Marker based validation of the *r3-r4* and *b2-b4* inversions in AMPL7.**

Two examples respecitvely for validating the *r3-r4* and *b2-b4* inversions. Similarly, these examples indicate different breakpoint intervals for the same type of AMPL7 rearrangements.

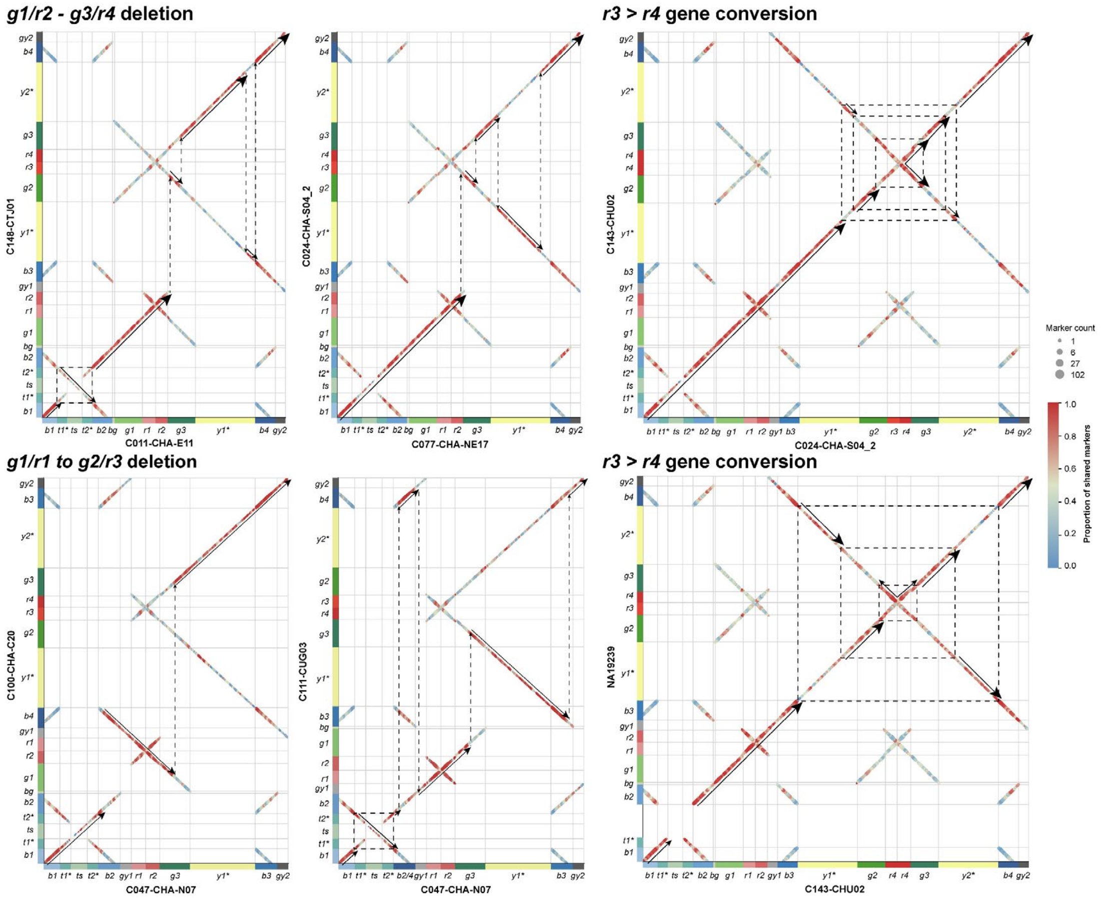


**Figure S39. Marker based validation of two micro-deletions and *r3>r4* gene conversions in AMPL7.**

Examples for validating the major micro-deletions and gene conversions. Legends are the same to the above figures.

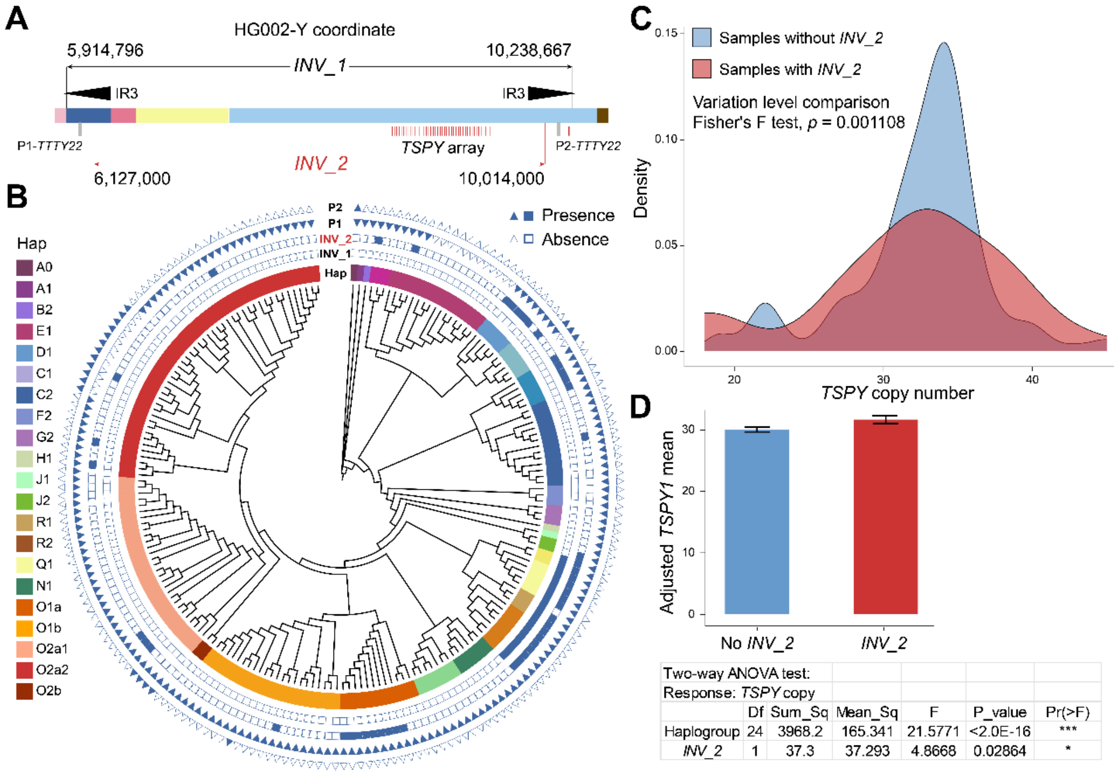


**Figure S40. The two inversions within the IR3 repeat.**

**A)** The genomic position of inversions whose breakpoints are located within IR3 repeats, with HG002-Y as reference. Besides that, the deletion of two *TTTY22* genes (gray bars) are coupled in *INV_1*. **B)** The presence and absence of two inversions and gene deletions across 206 haplogroups. ‘P1’ and ‘P2’ represent two *TTTY22* genes, respectively. **C)** The samples with *INV_2* show higher variation levels of *TSPY* copy number. **D)** *INV_2* could increase the *TSPY* copy number by 1.6, independent of haplogroup effects.

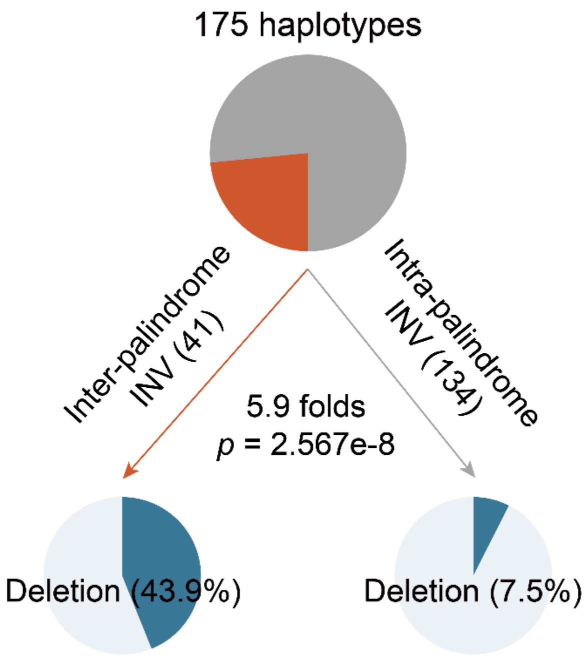


**Figure S41. Association of amplicon micro-deletions and inter-palindrome inversions.**
Pi-charts illustrate that inter-palindrome inversions are more likely to facilitate subsequent deletions than intra-palindrome inversions.

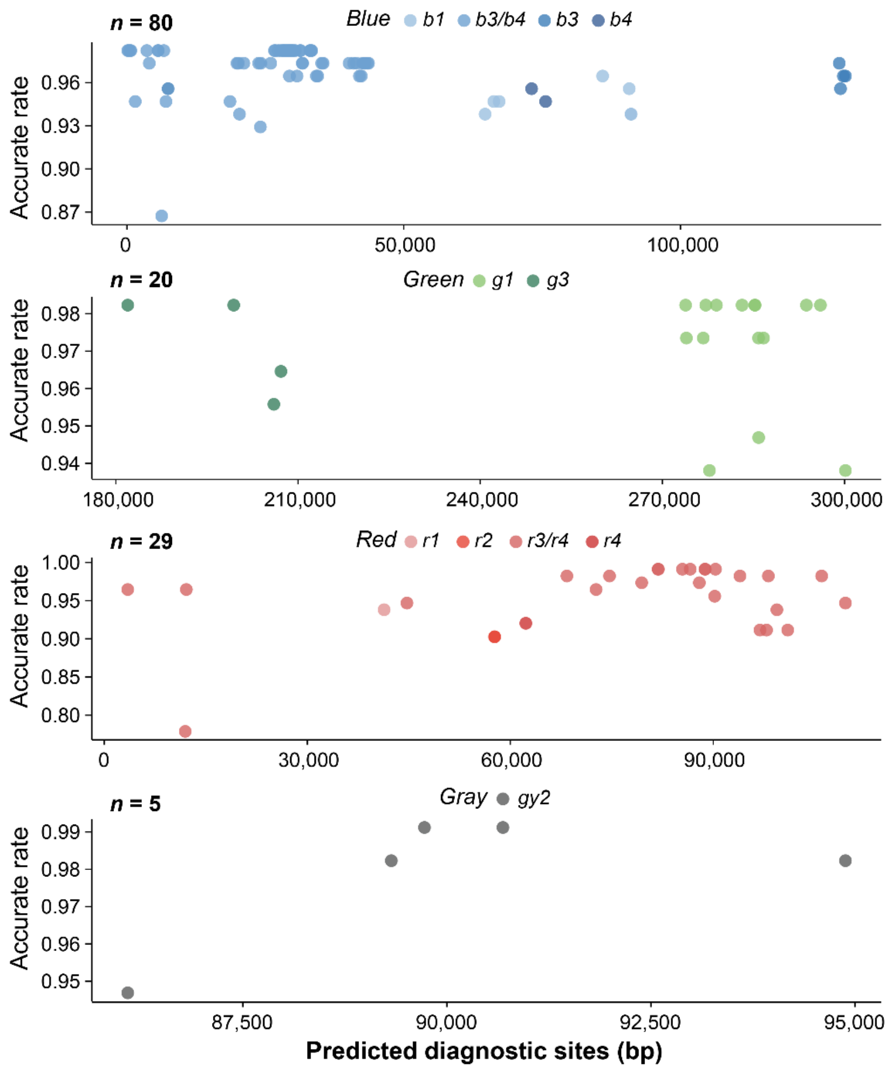


**Figure S42. The divergent sites identified between amplicon subgroups.**

The four amplicon families harbor different numbers of divergent sites. Dot colors indicate sites at which either the alternative or reference allele shows a high frequency in one or two specific amplicon subgroups, while remaining at low frequency in the other subgroups; only sites with frequency differences greater than 0.95 were retained. For example, dots shown in the lightest blue represent sites with high frequencies exclusively in the *b1* sub-amplicons but low frequencies in the other three subgroups. The x-axis denotes the genomic coordinates of these divergent sites on the reference sequence. Reference sequences for *r1*, *b2*, *g1*, and *gy1* amplicons were derived from the HG03248 sample, which carries the ancestral haplotype (CT). Using each divergent site as the central anchor, 31-mers corresponding to the reference (Ref.) and alternative (Alt.) alleles were extracted. These k-mers were subsequently used to assess the accuracy of each divergent site (Y-axis) by comparing their copy numbers across 175 AMPL7 haplotypes (**METHODS**). Specifically, the ones which have lost one or more sub-amplicons (e.g., N1 and Q1) will show different copies of Ref./Alt. k-mers on these sites.

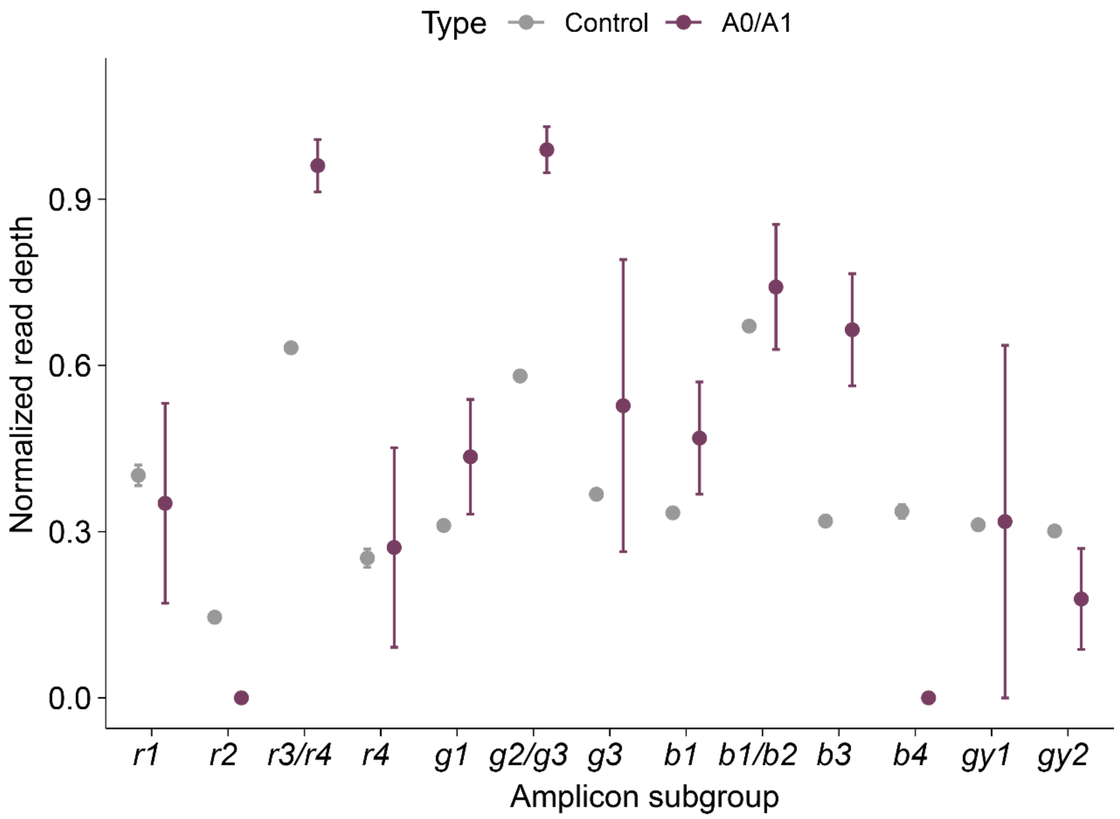


**Figure S43. Normalized short-read depth of k-mers (31-mer) specific to each subgroup of colored amplicons in samples of A0/A1 (*n* = 5) and CT descendent haplogroups.**

The X-axis indicates the amplicon subgroups defined from Figure 4E, whose depths of diagnostic k-mers were counted and normalized by the average read coverage of the XDR region. ‘Control’ represents individuals from CT haplogroup without any detected gene conversions or micro-deletions in the APML7 region.

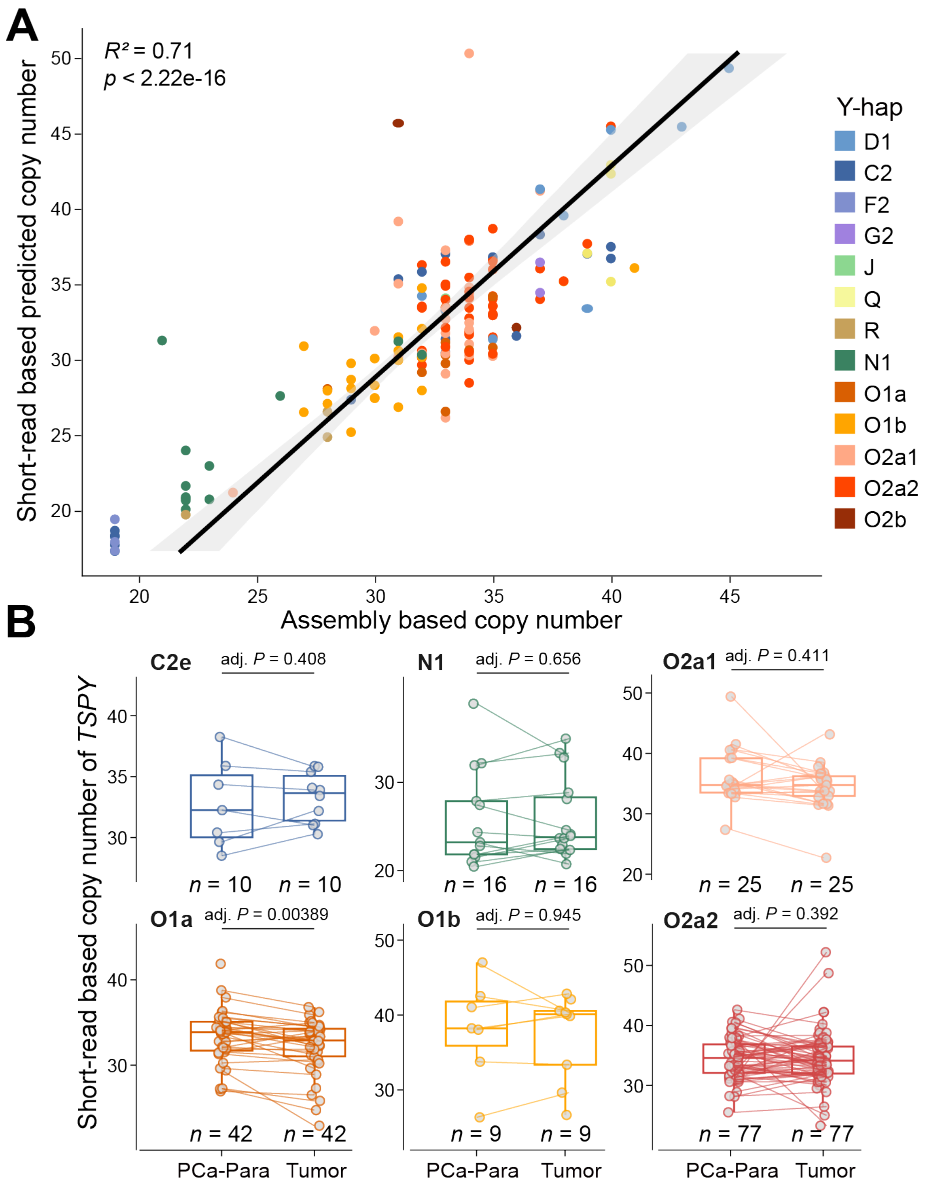


**Figure S44. Copy number difference of *TSPY* protein-coding genes between healthy samples from blood and prostate cancer (PCa) para-carcinoma tissue across Y-haplogroups.**

**A)** The correlation between short-read based predicted and assembly based *TSPY* gene copy for all the 160 Ys. **B)** The copy number was estimated based on short-read depth across the haplogroups with the individual number larger than 5. PCa-Para: Adjacent normal (clinical) tissue of prostate.

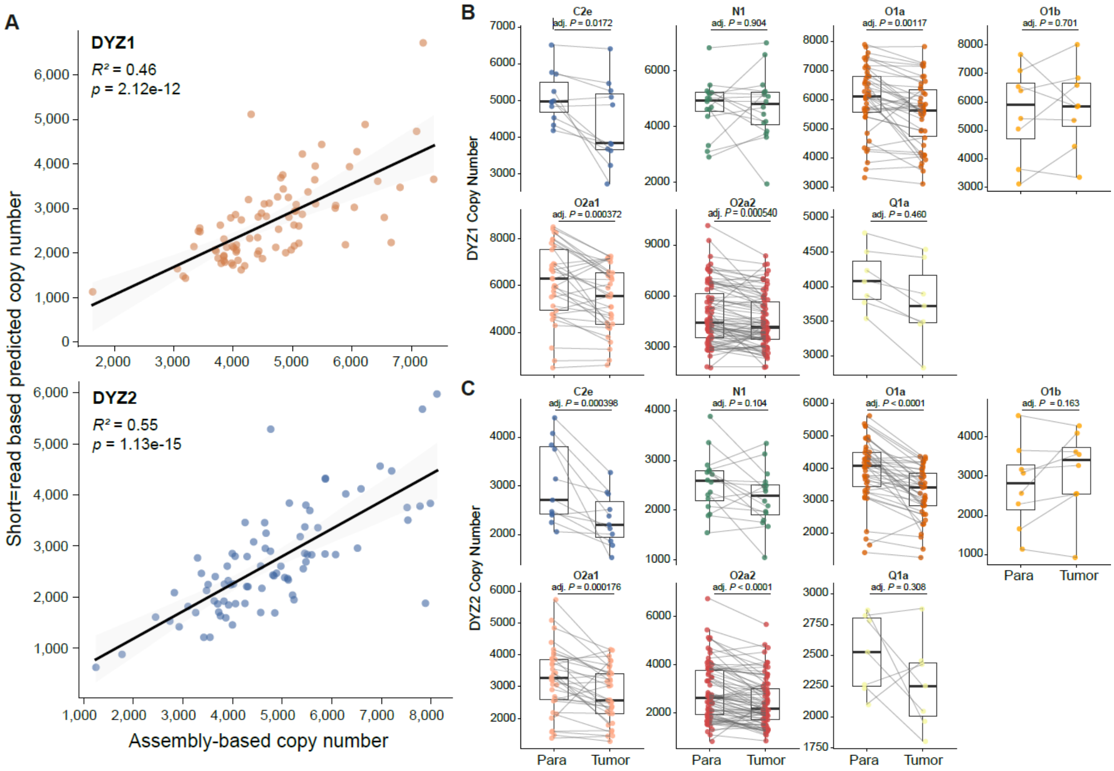


**Figure S45. NGS-based copy number estimation of DYZ1 and DYZ2.**

A) Correlation between short-read (MGISEQ) predicted and assembly-derived copy numbers for DYZ1 and DYZ2 in 85 gapless samples from the APGp1 dataset. Copy number was estimated using short-read depth. Significantly positive correlations were observed for both DYZ1 ($R^2$ = 0.46, $P$ = 2.12e-12) and DYZ2 ($R^2$ = 0.55, $P$ = 1.13e-15). The comparison of short-read estimated copy number for DYZ1 (B) and DYZ2 (C) satellite repeats between prostate cancer (PCa-Tumor) and adjacent normal (clinical) tissue (PCa-Para).

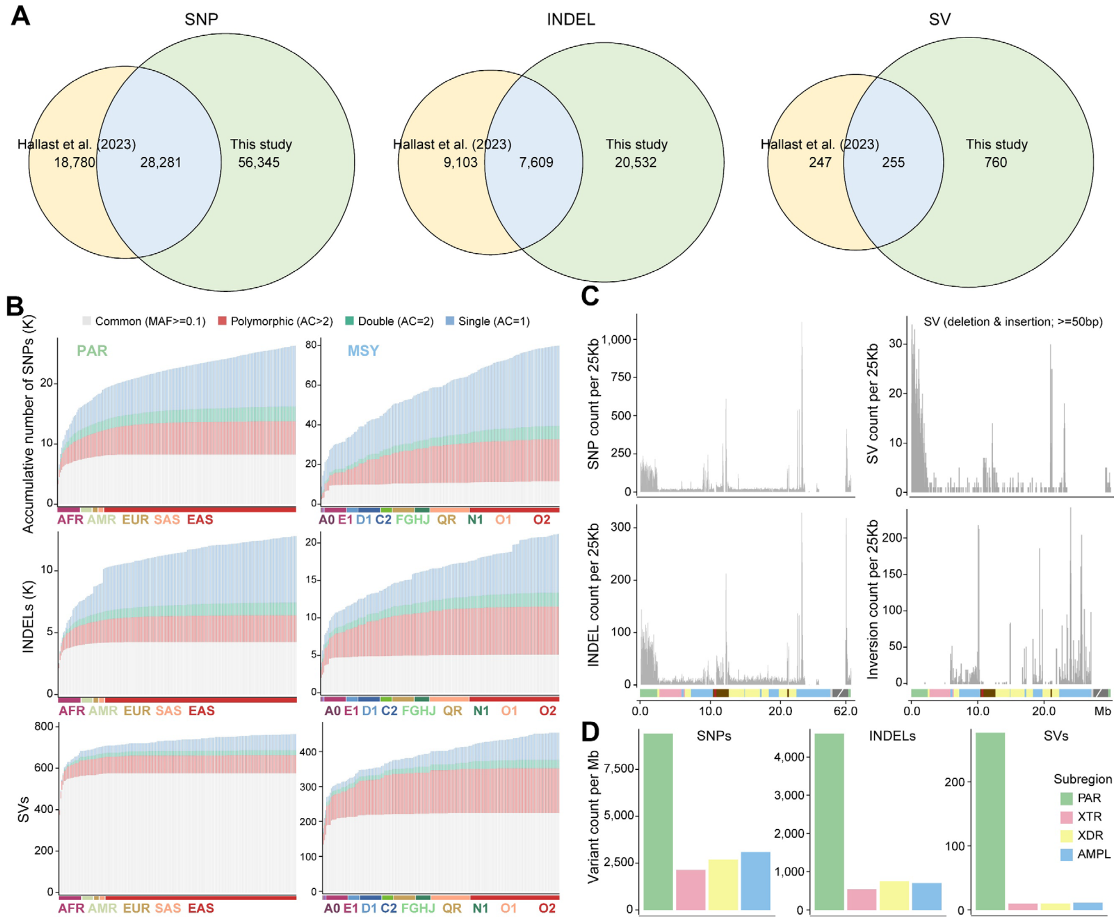


**Figure S46. The mutation profiles across subregions and populations or major haplogroups.**

**A)** The comparison of two variant sets for the euchromatic regions of human Y chromosomes. **B)** All types of mutation show similar patterns that populations share more mutations in PAR regions than haplogroups in MSY regions, due to the recombination suppression in the latter. Yet the mutation saturation is not achieved for MSY mutations, indicating hidden genetic diversity in MSY regions remaining to be explored. **C-D)** The PAR regions exhibit the highest SNP, INDEL and SV (deletions and duplications) density, except for several complex regions whose accurate alignment is difficult, e.g., centromeric and AMPL7 regions. The inversion density is the highest in AMPL regions.

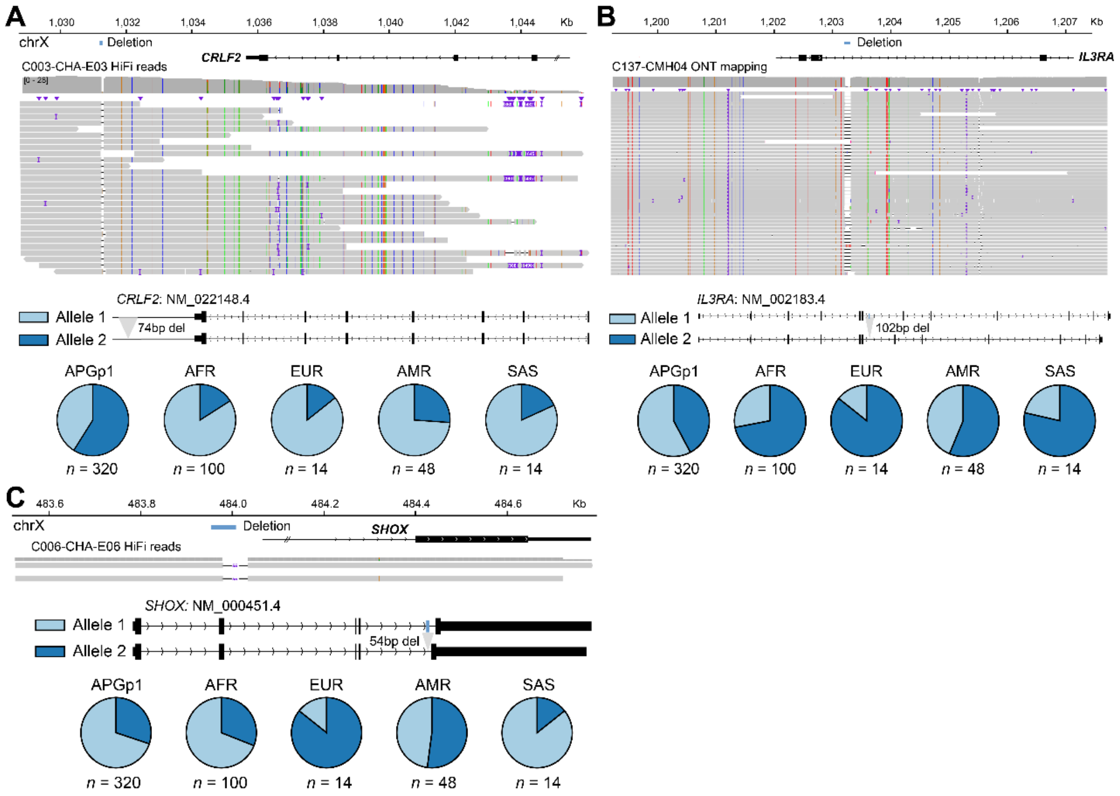


**Figure S47. The mutation profiles for three SV-related genes in PAR1.**
**A)** *CRLF2* (Cytokine receptor-like factor 2), **B)** *IL3RA* (Interleukin-3 receptor subunit alpha) and **C)** *SHOX* (Short stature homeobox). The IGV panels show the SV deletions. The pie-chart panels indicate the allele frequency across the continental populations. Allele 2 represents the derived allele using HG002-Y as reference, and the numbers below the pi-chats represent the population numbers.

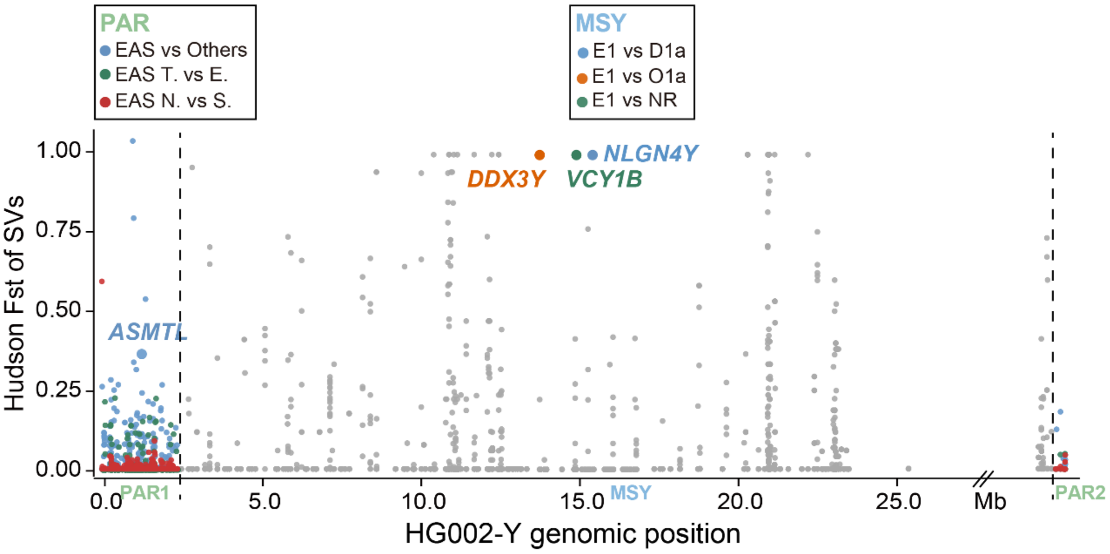


**Figure S48. The population or haplogroup differentiation of SVs.**
The dot-plot represents the Hudson's *Fst* for SVs, calculated between populations for PAR regions and between haplogroup E1 and other haplogroups for MSY regions. Each dot represents one SV. Four SVs are located near or within protein-coding genes (colored) and exhibit notably high lineage-specific differentiation.

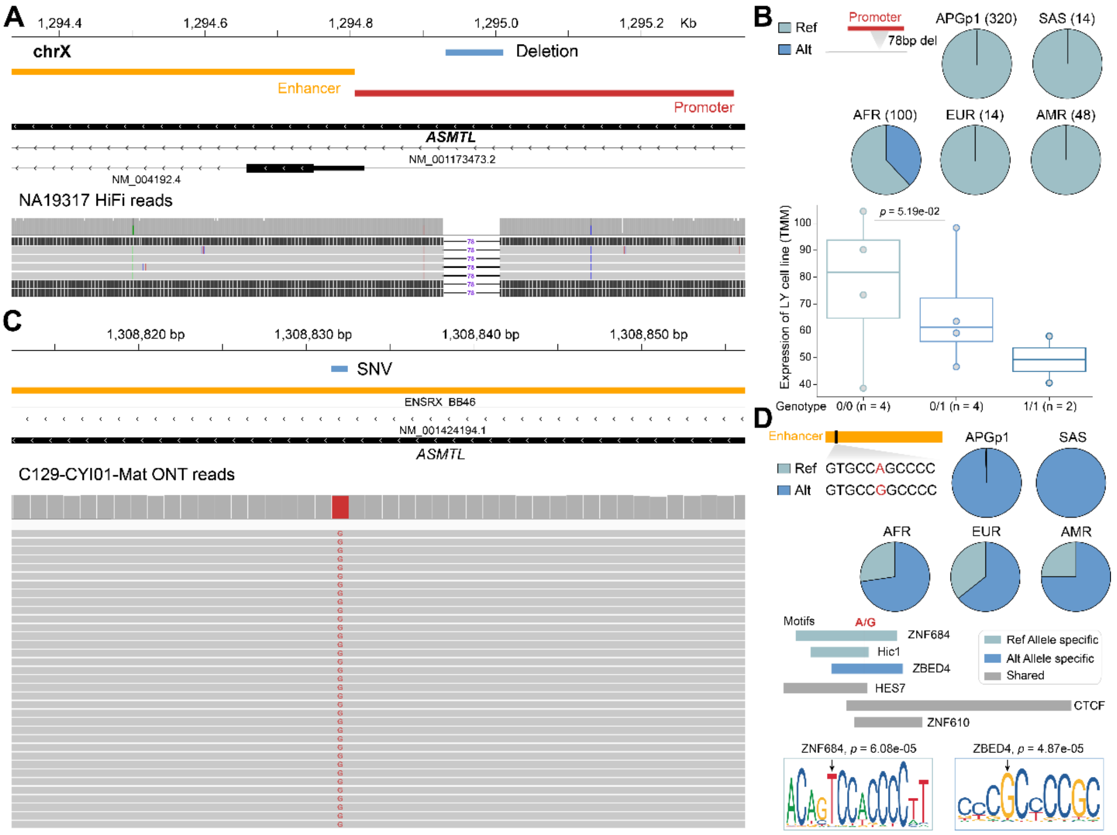


**Figure S49. Lineage differentiated mutations in the *ASMTL* gene.**

**A, C)** IGV snapshots for two mutations in *ASMTL* (Acetylserotonin O-methyltransferase-like) gene. Promoter and enhancer elements were colored red and yellow, respectively. **B, D)** A 78bp deletion in the promoter region of *ASMTL* gene shows high frequency in African populations, while a single nucleotide variant (SNV, **D**) in the adjacent enhancer is nearly fixed in East Asian and South Asian populations. The boxplot represents the expression level and genotypes for the deletion. The expression data was extracted from Multi-Ancestry Analysis of Gene Expression (MAGE)[17]. The SNV located on the nearby enhancer could alter the banding motifs compared to reference allele, implying potential function. Ly: lymphocyte.

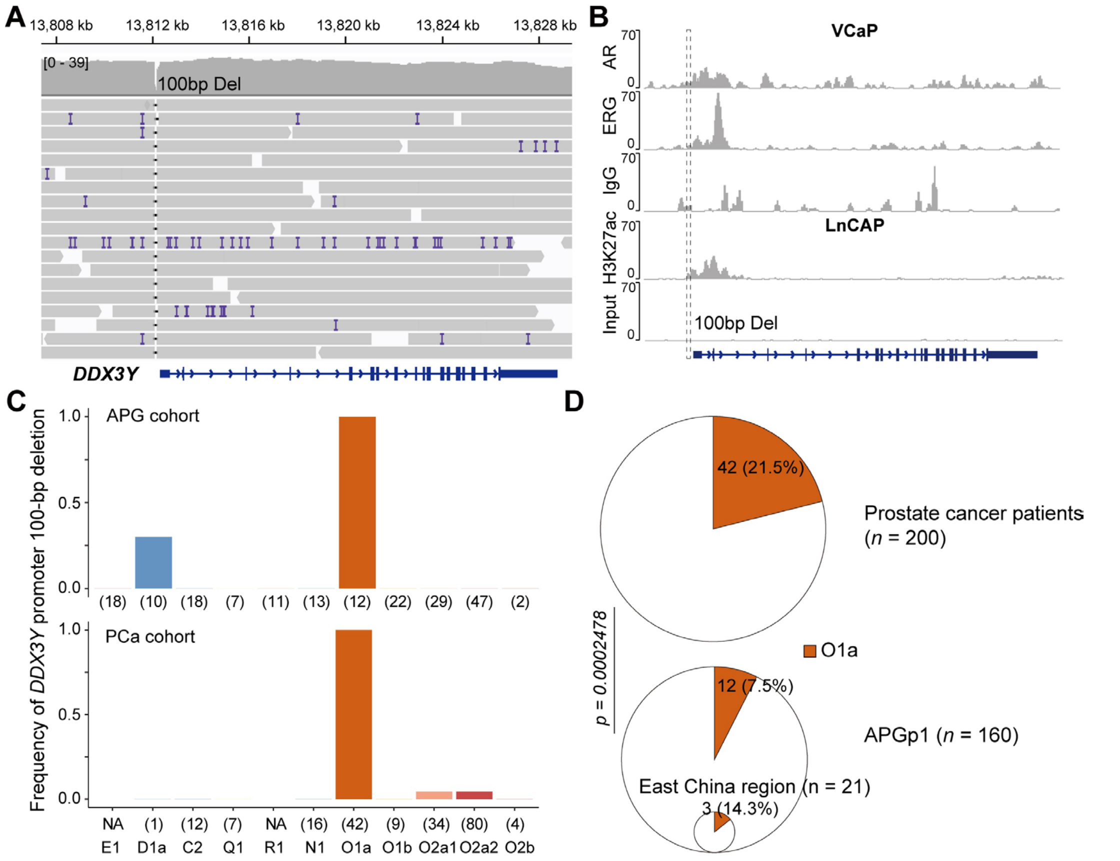


**Figure S50. The proportion of 100-bp deletion on the DDX3Y promoter and O1a in the APG and PCa cohort.**

**A)** The IGV screen shot of HiFi read alignments for validation of this deletion. **B)** The promoter 100bp-deletion is located in the banding region of *AR* (androgen receptor) and *ERG* (ETS transcription factor) in the prostate cancer cell lines. VCaP: Vertebral-Cancer of the Prostate; LNCaP: Lymph Node Carcinoma of the Prostate. **C)** The frequency of 100-bp deletion on the *DDX3Y* promoter in the APG and PCa cohort. The number in the bracket represents the individual number for each major Y haplogroup. **D)** The O1a haplogroup exhibits a significantly higher proportion in patients with PCa cohort compared to the APG cohort. This difference of O1a proportion between the two cohorts remains when comparing among the samples from the East-China region.

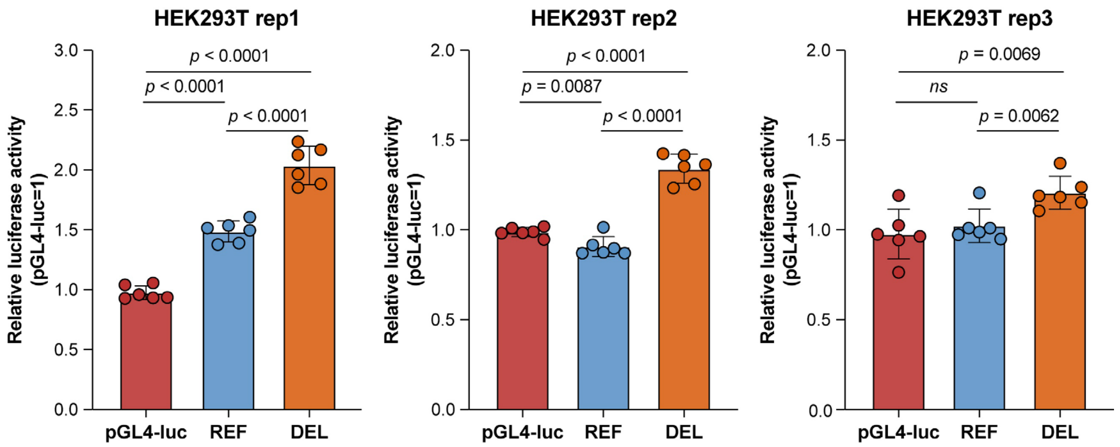


**Figure S51. The reporter assay for the 100-bp deletion in the *DDX3Y* promoter region.**

The x-axis shows the control construct (pGL4-luc), the 100-bp deletion allele (200-bp sequence lacking the deleted fragment), and the reference allele (300-bp sequence containing the deleted fragment). The y-axis indicates normalized luciferase activity. Reporter assays were performed in cultured HEK293T (human embryonic kidney 293T) cells.

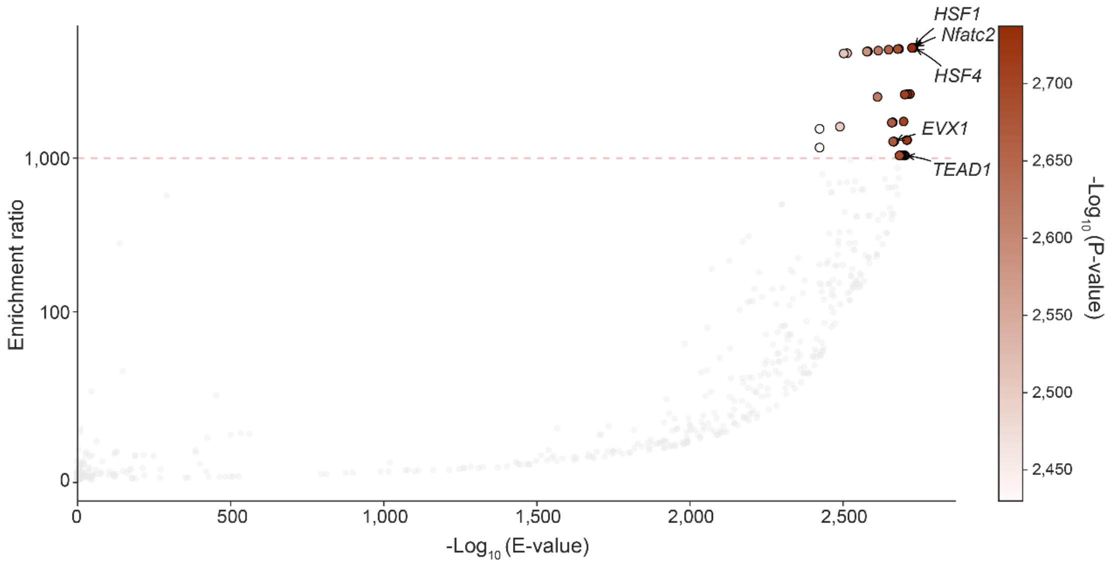


**Figure S52. Motif enrichment analysis of Yq12 DYZ1 using MEME-SEA.**
Each point represents a transcription factor binding motif tested for enrichment in DYZ1 regions relative to the DYZ2 regions. The X-axis shows the -$\log_{10}$ (E-value), which accounts for multiple testing across all evaluated motifs, and the Y-axis indicates the enrichment ratio (observed motif frequency in target regions divided by that in background regions). Motifs with both high enrichment ratios and low E-values represent strongly and specifically enriched binding signatures. Motifs related to prostate cancer are labeled.

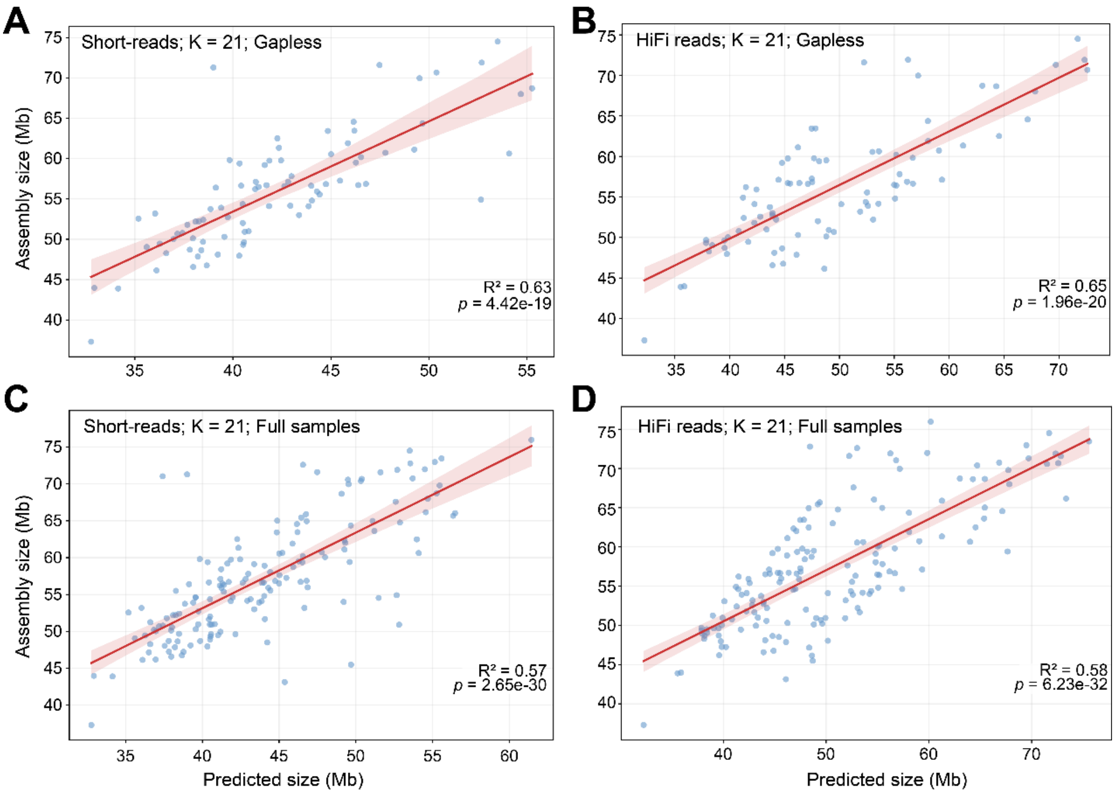


**Figure S53. The comparison of predicted and assembled size of Y chromosomes.**

Each scatter represents one sample. These plots show strong positive correlations between Y-assembly size and predicted size based on the k-mer frequency spectrum of both NGS reads (left panels) and HiFi reads (right panels), with k-mer size of 21. **A-B)** Only gapless samples were used for comparison. **C-D)** All samples were used for comparison.

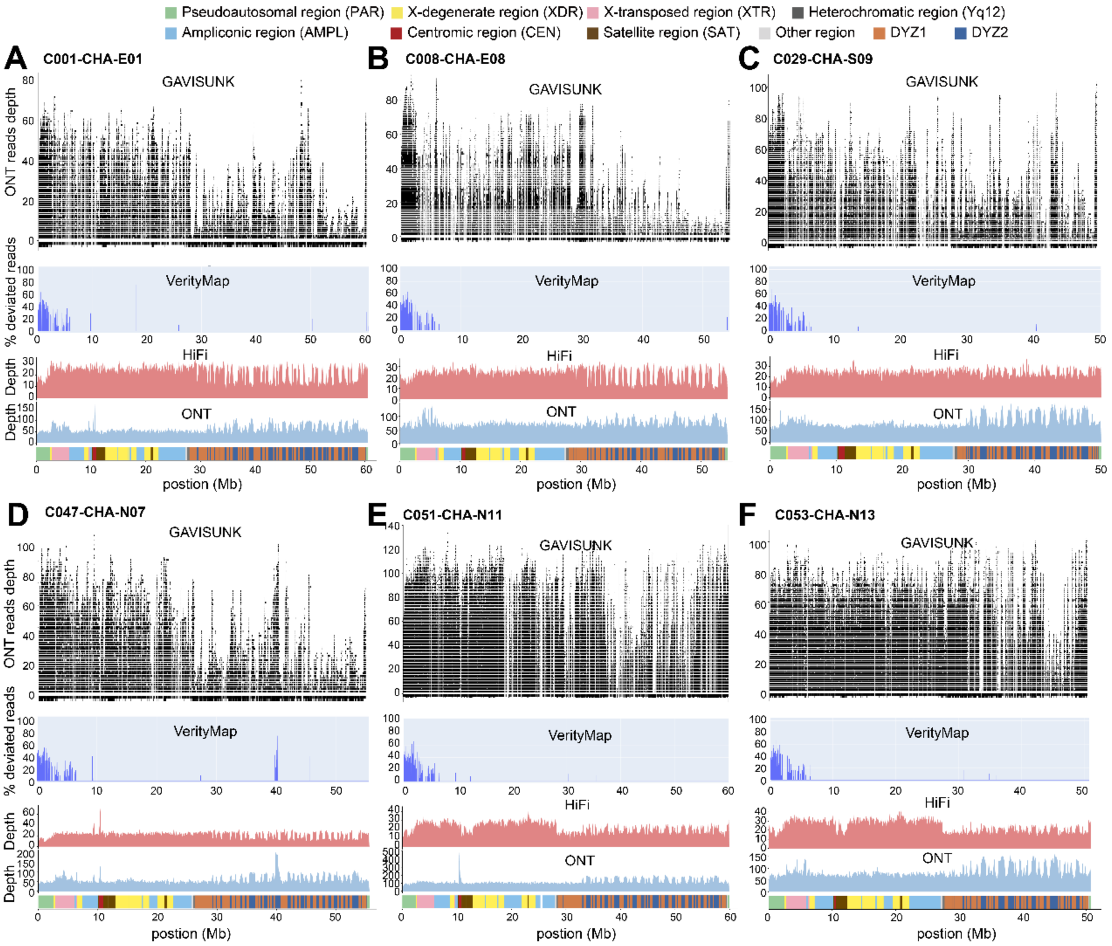


**Figure S54. Y chromosome quality assessment using k-mer validation and sequencing depth metrics.**

**A-F** panels show six representative Y chromosome assemblies with five stacked annotation and evaluation tracks. From upper to lower panels, they are: **1)** GAVISUNK[18] validation. Concordant SUNK k-mer matches are shown as full-height black tick marks, indicating validated regions with high confidence. **2)** VerityMap[19] plot (200bp window) highlights potential assembly discrepancies. Blue bars indicate sites with deviated read proportions: potential heterozygous variants (20-80% deviation) and likely assembly errors (>80% discordant reads). **3)** HiFi sequencing depth. Depth coverage from high-fidelity reads shown in red, reflecting base-level consistency across the chromosome. **4)** ONT sequencing depth. Long-read Oxford Nanopore coverage in blue, useful for identifying dropouts and structural inconsistencies. **5)** Subregion annotation. Color-coded genomic subregions include PAR, XDR, XTR, AMPL, CEN, Satellite (SAT), HET (Yq12), DYZ1, DYZ2, and other regions.

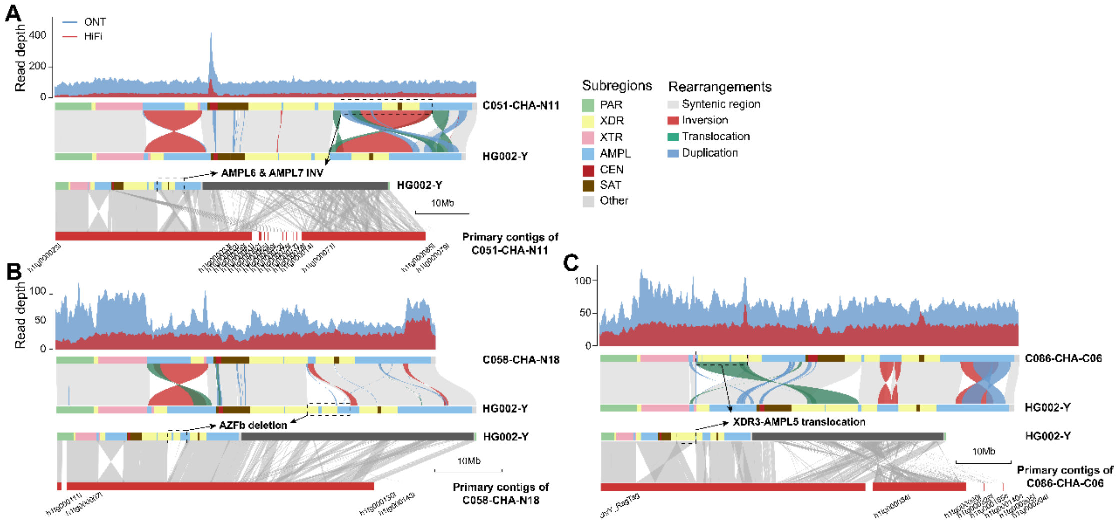


**Figure S55. The validation for large and rare rearrangements through read depth and primary contig synteny.**

**A)** The inversion (*INV_9*) spanning AMPL6-7 (with black dashed box) in C051-CHA-N11 are supported by both HiFi/ONT read depth and primary contig alignment with HG002-Y. The same applies for the deletion that overlaps the AZFb region in C058-CHA-N18 **(B)** and XDR3-AMPL5 translocation in C086-CHA-C06 **(C)**.

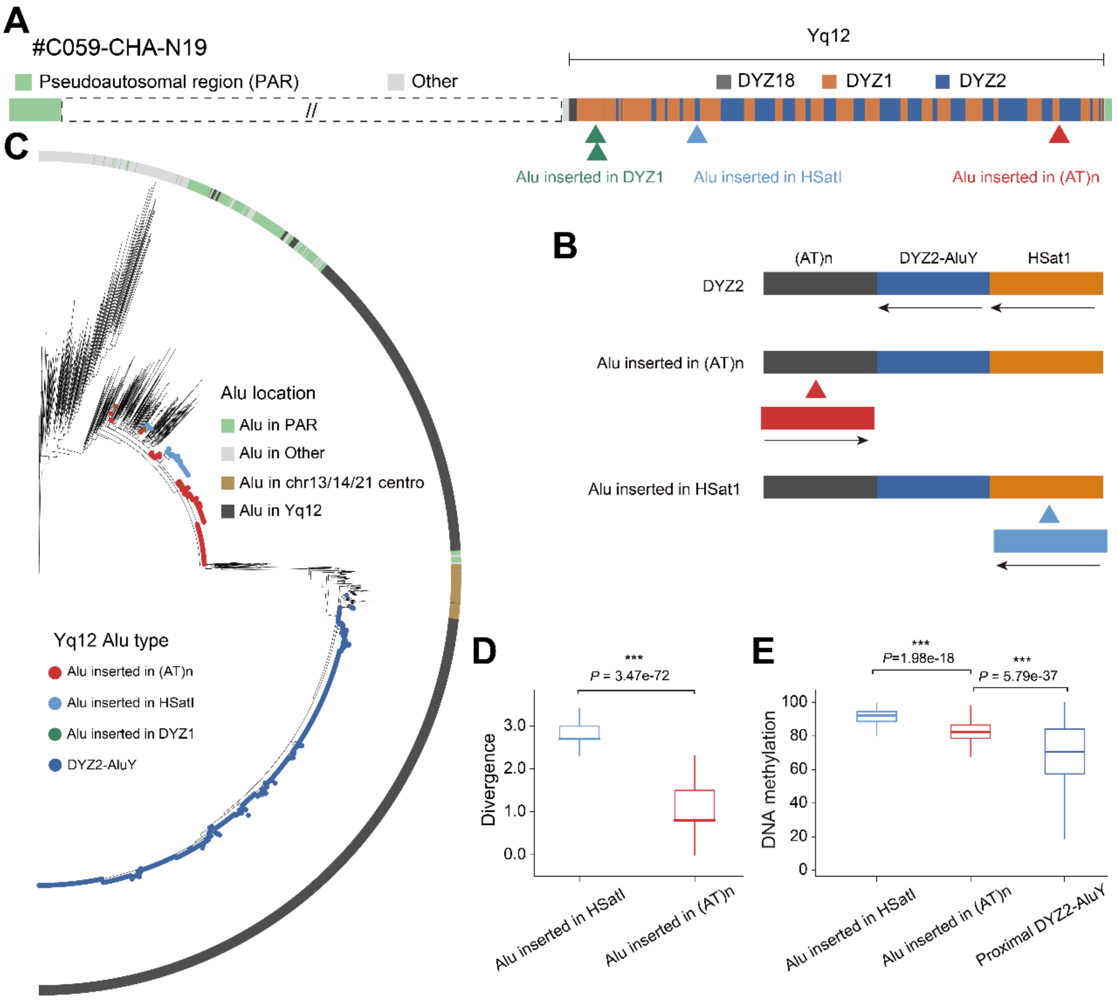


**Figure S56. *Alu* Insertions in the Yq12 region.**
**A)** Annotation of Y chromosome heterochromatin regions. **B)** *Alu* insertion patterns in DYZ2: Inserted in AT-repeat region: *Alu* elements are inserted in reverse orientation relative to DYZ2-*AluY*. Inserted in *HSatI* region: *Alu* elements are inserted in forward orientation.
**C)** Phylogenetic analysis of *Alu* elements (rooted with *AluS*), including DYZ2-*AluY*, Inserted *Alu* in (AT)n or *HSatI* regions (this study), Inserted *Alu* in DYZ1, *Alu* from other regions of Y chromosomes; *Alu* from other chromosomes. **D)** Divergence of inserted *Alu*: Pairwise divergence (Kimura 2-parameter distance) is significantly higher for *Alu* in (AT)n repeats compared to *HSatI* ($P < 0.001$, Mann-Whitney test). **E)** Methylation levels of inserted Alu: Methylation in inserted Alu compared to adjacent DYZ2-*AluY* ($p < 0.001$, Mann-Whitney test).

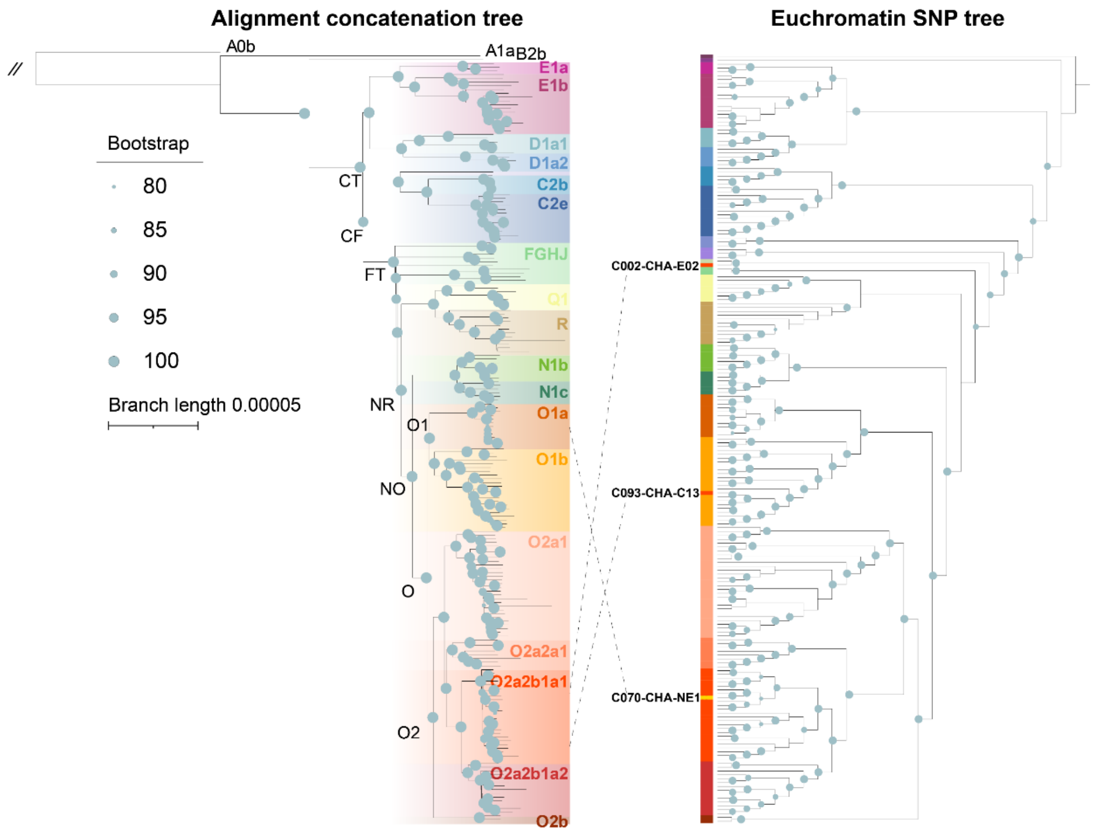


**Figure S57. The phylogeny comparison of alignment-based concatenation tree and SNP-based tree.**

The haplogroup information of three samples (C002-CHA-E02, C070-CHA-NE1 and C093-CHA-C13) was corrected based on the concatenation tree. The blue dots represent phylogeny bootstrap supports.

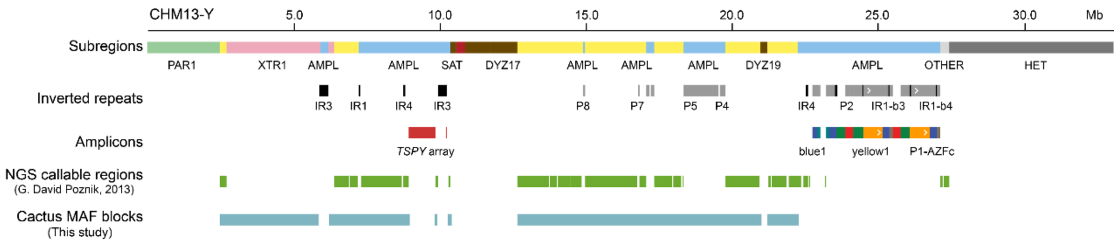


**Figure S58. The euchromatic regions used for tree construction in this study.** Compared to previously NGS (Next-generation sequencing) callable regions, the assembly alignment-based way has greatly expanded analyzable sequences, especially the XTR regions. The NGS callable regions were extracted from previous work[20] and further transformed into HG002-Y based coordinates.

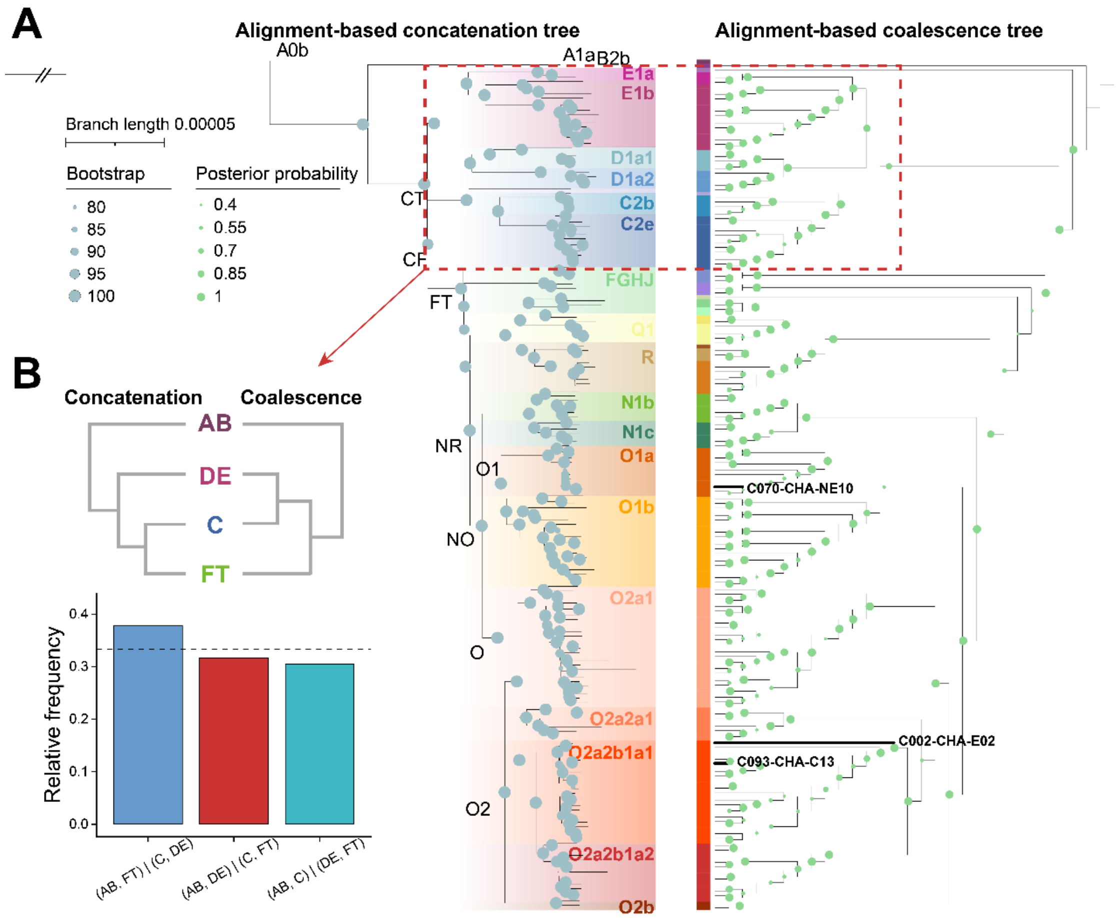


**Figure S59. The phylogenetic discordance of concatenation and coalescence trees.**

**A)** The coalescence was produced by Astral-III software[21] using 1,657 gene trees, which were generated from local alignment blocks using IQ-TREE software[22]. The major difference between the two trees is on the DE and C/FT nodes, illustrated in **B)**. The alternative topology (AB, FT) | (C, DE) receives a relatively higher supports of 50Kb gene trees, although the bootstrap values were high for these nodes in the concatenation tree which supports the topology of (AB, DE) | (C, FT). The three samples with different haplogroup definition in concatenation and SNP trees are highlighted on the coalescence tree. Their phylogenetic positions are consistent in two alignment-based trees.

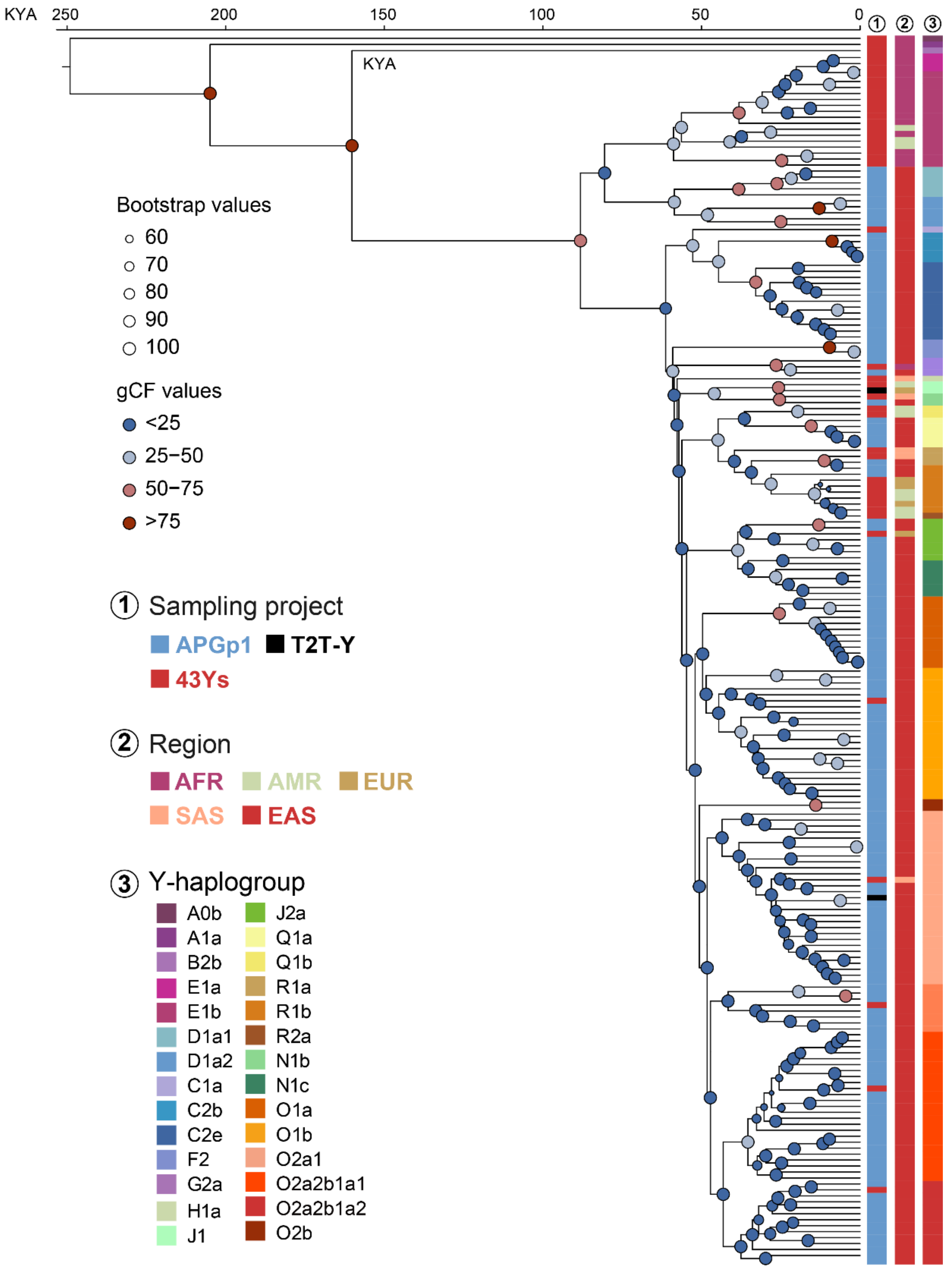


**Figure S60. The divergence time of all 206 worldwide Ys.**
The time was inferred from Cactus based whole-Y alignments using MCMCtree[23]. Two calibration points were used: **A)** Calibration point of A0b: 254 (95% CI: 192 - 307) KYA; **B)** Calibration point of CT: 71.0 (95% CI: 53.8 - 85.8 KYA)[24]. 'T2T-Y' includes both HG002-Y and CN1-Y. gCF: gene concordance factor.

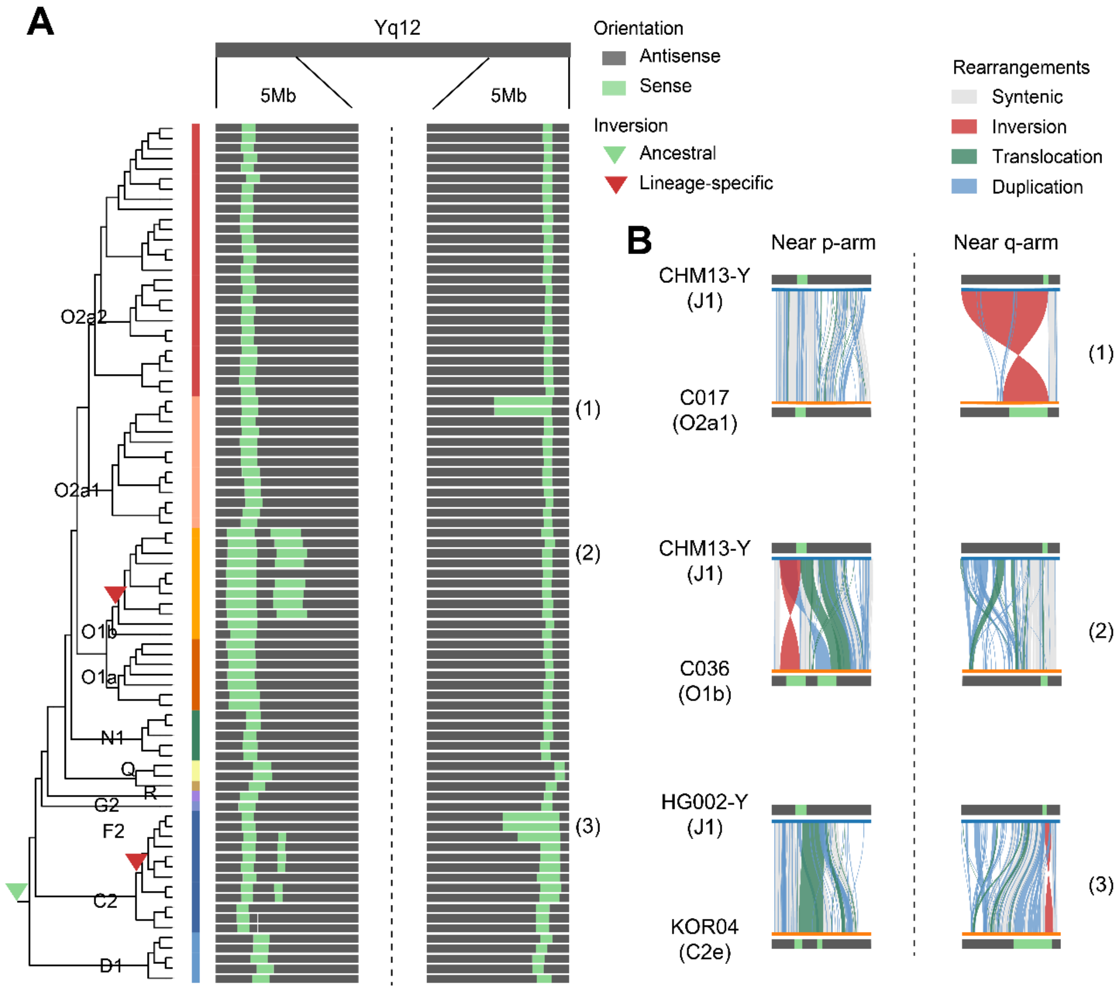


**Figure S61. Structural landscape of the heterochromatic region (Yq12) reveals conserved and lineage-specific inversions.**

**A)** Inversion patterns across the Yq12 region in 85 gapless Y chromosome assemblies from this study, mapped against the HG002-Y reference. Orientation of each region is represented in green (sense) and gray (antisense). Two large inversions are consistently observed at the distal boundaries of Yq12 across all haplogroups, indicating a shared structural transition that separates the array into conserved and expanded domains. In addition to the conserved inversions, lineage-specific inversions (marked with red triangles) are identified in haplogroups O1a, O1b and F2. **B)** Synteny plots between representative assemblies and the HG002-Y reference illustrate three structural configurations, with colored links between assemblies representing synteny (gray), inversions (red), translocations (green), and duplications (blue). For the selected samples: 1) samples with expanded near-q-arm inverted regions, 2) samples with an additional near-p-arm inversion, 3) samples with an additional near-p-arm inversion and expanded near-q-arm inverted region.

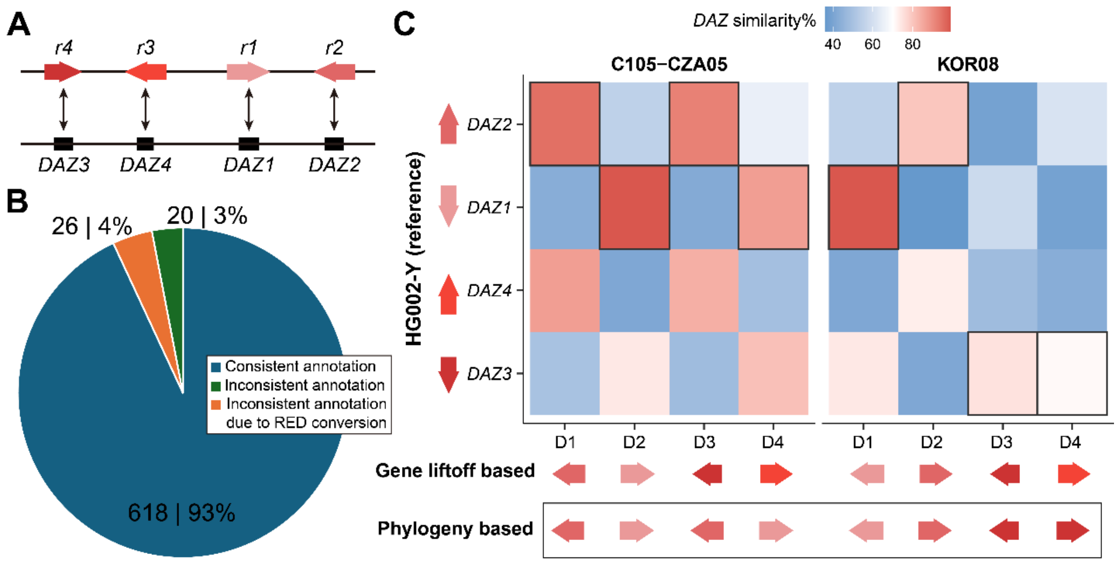


**Figure S62. The consistency of phylogenetic classification and gene-liftover annotation.**

**A)** For HG002-Y, the red amplicons and *DAZ* (Deleted in Azoospermia) genes have one-to-one correspondence. **B)** For all samples, the consistency rate of RED classification and *DAZ* gene-liftoff annotation is 93%. **C)** Two samples with annotation inconsistency were selected for validation. The heatmap is the sequence similarity between *DAZ* genes of HG002-Y (Reference, y-axis) and individuals (x-axis). The 'D1-D4' represents the *DAZ* genes according to the coordinate sequences. Specifically, though the third and fourth *DAZ* genes of the C105-CZA05 sample are annotated as *DAZ3* and *DAZ4* (darker red colors) by gene-liftoff method, their similarity with HG002-Y *DAZ2* and *DAZ1* is instead higher (black boxes), corresponding to the phylogeny-based classification (two *DAZ1/2*, lighter red colors). The same goes for the KOR08 sample, in which the fourth *DAZ* gene should be annotated as *DAZ3* (the darkest red color) according to the highest similarity with *DAZ3* of HG002-Y, rather than *DAZ4* by the gene-liftoff method. These comparisons suggest the limitation of liftoff-based gene annotation for duplicated genes.

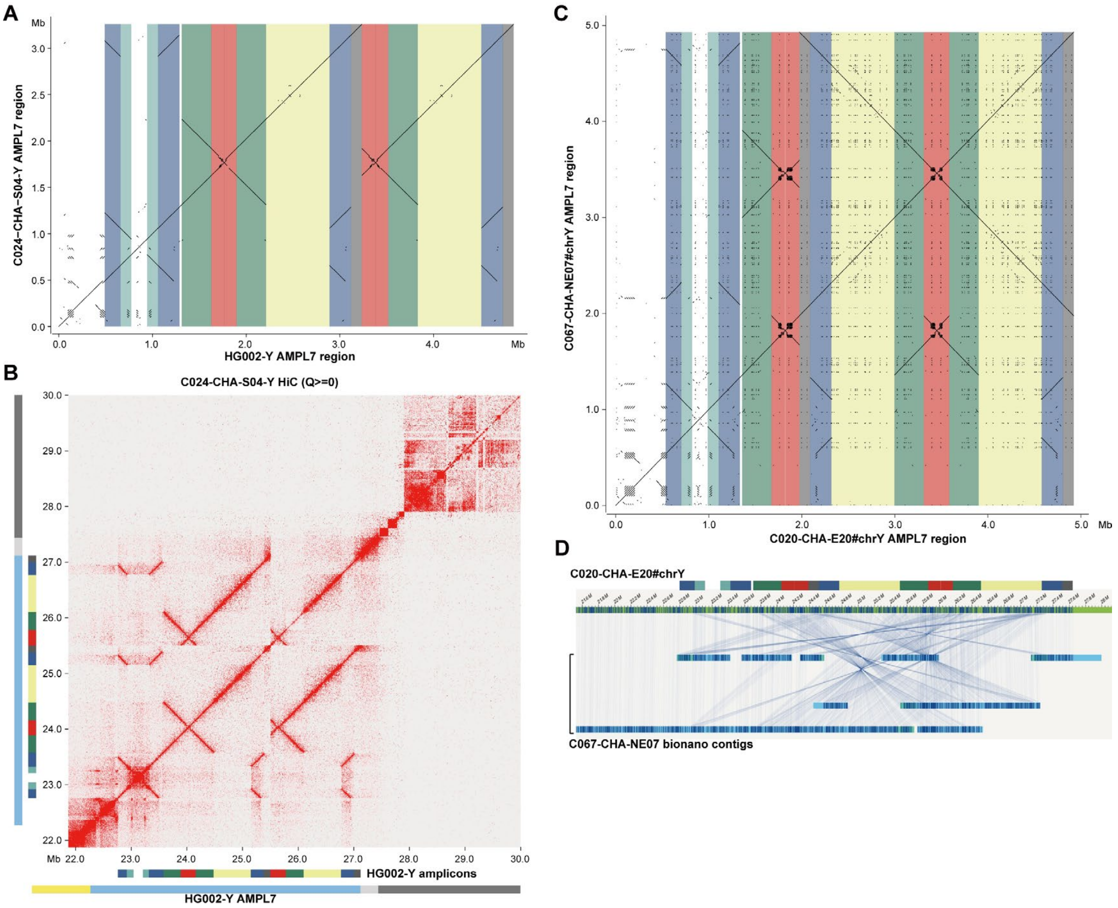


**Figure S63. The Hi-C and bionano reads are not effective to detect or validate rearrangements in AMPL7 region.**

**A)** The C024-CHA-S04 exhibits micro-deletion compared to the HG002-Y reference (repeat masked). **B)** The heatmap of Hi-C interaction of the C024 individual with HG002-Y as reference. No filtering regarding read mapping quality was applied to retain Hi-C reads in AMPL7 region. **C)** The individual C067 exhibits the same AMPL7 structure compared to C020-Y reference (repeat unmasked), yet the alignment of bionano read based contigs of C067 to the C020 genome **D)** was too chaotic to provide valid structural information.
